# The risks of autoimmune- and inflammatory post-acute COVID-19 conditions: a network cohort study in six European countries, the US, and Korea

**DOI:** 10.1101/2024.05.15.24307344

**Authors:** Theresa Burkard, Kim López-Güell, Martí Català, Edward Burn, Antonella Delmestri, Sara Khalid, Annika M Joedicke, Daniel Dedman, Jessie O Oyinlola, Alicia Abellan, Laura Pérez-Crespo, Núria Mercadé-Besora, Talita Duarte-Salles, Daniel Prieto-Alhambra, Johnmary T Arinze, Mees Mosseveld, Raivo Kolde, Jaime Meléndez-Cardiel, Raúl López-Blasco, Álvaro Martínez, Bernardo Valdivieso, Dominique Delseny, Gregoire Mercier, Chungsoo Kim, Ji-woo Kim, Kristin Kostka, Juan Manuel Ramírez-Anguita, Miguel A Mayer, Nhung TH Trinh, Hedvig ME Nordeng, Roger Paredes, Anneli Uusküla, Akihiko Nishimura, Cora Loste, Lourdes Mateu, Junqing Xie

**Affiliations:** Nuffield Department of Orthopaedics, Rheumatology and Musculoskeletal Sciences, University of Oxford, Oxford, UK; CPRD, Medicines and Healthcare products Regulatory Agency, London, UK; Fundació Institut Universitari per a la recerca a l’Atenció Primària de Salut Jordi Gol i Gurina (IDIAPJGol), Barcelona, Spain; Department of Medical Informatics, Erasmus University Medical Center, Rotterdam, the Netherlands; Institute of Computer Science, University of Tartu, Tartu, Estonia; Biocomputing Unit, Aragon Health Sciences Institute (IACS), Zaragoza, Spain; The Health Research Institute Hospital La Fe (Instituto de Investigación Sanitaria La Fe), Avenida Fernando Abril Martorell, 106 Torre A 7a planta, 46026, Valencia, Spain; The University and Polytechnic La Fe Hospital of Valencia, Avenida Fernando Abril Martorell, 106 Torre H 1a planta, 46026, Valencia, Spain; Public Health Department, University Hospital of Montpellier, 34295 Montpellier, France; IDESP, Université de Montpellier, INSERM, 34000 Montpellier, France; Department of Biomedical Sciences, Ajou University Graduate School of Medicine, Suwon, Republic of Korea; Big Data Department, Health Insurance Review and Assessment Service, Wonju, Republic of Korea; The OHDSI Center at the Roux Institute, Northeastern University, Portland, ME, USA; Parc de Salut Mar, Hospital del Mar Medical Research Institute, Barcelona, Spain; Pharmacoepidemiology and Drug Safety Research Group, Department of Pharmacy, Faculty of Mathematics and Natural Sciences, University of Oslo, Oslo, Norway; Department of Child Health and Development, Norwegian Institute of Public Health, Oslo, Norway; Center for Global Health and Diseases, Department of Pathology, Case Western Reserve University School of Medicine, Cleveland, OH; Department of Infectious Diseases & irsiCaixa AIDS Research Institute, Hospital Universitari Germans Trias i Pujol, Badalona, Catalonia, Spain; Chair in Infectious Diseases and Immunity, Center for Health and Social Care Research (CEESS), Faculty of Medicine. University of Vic-Central University of Catalonia (UVic-UCC).; Institute of Family Medicine and Public Health, University of Tartu, Tartu, Estonia; Department of Biostatistics, Bloomberg School of Public Health, Johns Hopkins University, Baltimore, MD, USA; REICOP (Red de Investigación Covid Persistente), Madrid, Spain; Universitat Autònoma de Barcelona, Catalonia, Spain

**Keywords:** COVID-19, postural orthostatic tachycardia syndrome, myalgic encephalomyelitis / chronic fatigues syndrome, ME/CFS, rheumatoid arthritis, juvenile idiopathic arthritis, systemic lupus erythematosus, inflammatory bowel disease, multiple inflammatory syndrome, PACS, PASC, post-COVID-19 conditions, PCC, post-acute sequelae of COVID-19, epidemiology, disease burden, network study, cohort study, SARS-Cov-2

## Abstract

**Objectives:** We aimed to assess the risk of autoimmune- and inflammatory post-acute COVID-19 conditions.

**Design:** Descriptive network cohort study.

**Setting:** Electronic health records from UK and Dutch primary care, Norwegian linked health registry, hospital records of specialist centres in Spain, France, and Korea, and healthcare claims from Estonia and the US.

**Participants:** We followed individuals between September 2020 and the latest available data from the day they fulfilled at least 365 days of prior observation (general population), additionally from day 91 after a SARS-Cov-2 negative test (comparator) or a COVID-19 record (exposed patients).

**Main outcome measures:** We assessed postural orthostatic tachycardia syndrome (POTS) diagnoses/symptoms, myalgic encephalomyelitis / chronic fatigues syndrome (ME/CFS) diagnoses/symptoms, multi-inflammatory syndrome (MIS), and several autoimmune diseases. For contextualisation, we assessed any diabetes mellitus (DM).

Meta-analysed crude incidence rate ratios (IRR) of outcomes measures after COVID-19 versus negative testing yield the ratios of absolute risks. Furthermore, incidence rates (IR) of the outcomes in the general population describe the total disease burden.

**Results:** We included 34’549’575 individuals of whom 2’521’812 had COVID-19, and 4’233’145 a first negative test. After COVID-19 compared to test negative patients, we observed IRRs of 1.24 (1.23-1.25), 1.22 (1.21-1.23), and 1.12 (1.04-1.21) for POTS symptoms, ME/CFS symptoms and diagnoses, respectively. In contrast, autoimmune diseases and DM did not yield higher rates after COVID-19. In individual general database populations, IRs of POTS and ME/CFS diagnoses were 17-1’477/100’000 person-years (pys) and 2-473/100’000 pys, respectively. IRs of MIS were lowest with IRs 0.4-16/100’000 pys, those of DM as a benchmark 8-86/100’000 pys. IRs largely depended on the care setting.

**Conclusion:** In our unmatched comparison, we observed that, following COVID-19, POTS and ME/CFS yielded higher rates than after negative testing. In absolute terms, we observed POTS and ME/CFS diagnoses to have a similar disease burden as DM.

**WHAT IS ALREADY KNOWN ON THIS TOPIC:**

- Observational research suggested positive associations between COVID-19 and so called post-acute COVID-19 conditions, whose spectrum is yet to be established
- Basic research suggested pathways that link COVID-19 with autoimmune- and inflammatory diseases such as postural orthostatic tachycardia syndrome (POTS), myalgic encephalomyelitis / chronic fatigues syndrome (ME/CFS), multiple inflammatory syndrome (MIS), and autoimmune diseases

**WHAT THIS STUDY ADDS:**

- After COVID-19, the rates of POTS symptoms and ME/CFS symptoms/diagnoses was higher than those after negative testing
- After COVID-19 versus negative testing, rates of ME/CFS diagnoses were increased in the working age group and rates of symptoms of POTS and ME/CFS were increased in children and elderly
- Disease burdens of POTS and ME/CFS diagnoses in the general population were higher among women than among men and overall similar to that of diabetes mellitus

## INTRODUCTION

The aftermath of COVID-19 continues to reveal a complex landscape of health challenges, with a significant subset of individuals experiencing prolonged symptoms beyond the acute phase of infection.[1,2] Characterized by a diverse array of persistent symptoms encompassing fatigue, cognitive dysfunction, and dysautonomia, it poses a challenge to both patients and healthcare providers.[3] Observational studies have shed light on its prevalence, yet with a large spread depending on the population, settings, and most importantly the disease definition which is yet to be established. Worldwide estimates from meta-analysed data indicate that around one third of individuals may experience prolonged symptoms following acute COVID-19, and up to half of hospitalised COVID-19 patients.[4,5] On one hand, the close interplay between the immune system and the autonomic nervous system underscores the link between COVID-19 and conditions like postural orthostatic tachycardia syndrome (POTS).[6–12] On the other hand, the precise link between COVID-19 and myalgic encephalomyelitis / chronic fatigue syndrome (ME/CFS) is still being elucidated. Emerging evidence suggests that COVID-19 infection may predispose some individuals to develop ME/CFS-like symptoms or exacerbate pre-existing ME/CFS.[12–15] Additionally, the emergence of multisystem inflammatory syndrome (MIS) in children (MIS-C) and adults (MIS-A) underscores the diverse inflammatory responses elicited by the virus.[16] Finally, COVID-19 has been shown to trigger dysregulated immune responses, including the production of autoantibodies and the activation of inflammatory pathways, which may predispose individuals to the development or exacerbation of autoimmune conditions.[17] Observational research suggests positive associations of COVID-19 with inflammatory disorders such as rheumatoid arthritis (RA), inflammatory bowel disease (IBD), or systemic lupus erythematosus.[18–20]

Elucidating the risk of POTS, ME/CFS, MIS, and autoimmune diseases is paramount for understanding their impact on individuals, society, and healthcare systems. Estimations of disease burdens are essential for resource allocation, healthcare planning, and developing effective strategies for diagnosis, treatment, and support services. Therefore, we aimed to assess relative and absolute rates of these conditions in several countries and populations.

## METHODS

### Study design

We performed a descriptive cohort study in an international network using routinely collected healthcare data mapped to the observational medical outcomes partnership (OMOP) common data model (CDM). Each site ran the study using a common analytical code and shared back results without sharing patient level data.[21–23]

### Data sources

Nine databases from six European countries, the US, and Korea contributed data to this study. Table 1 lists an overview of the characteristics of the individual databases and decodes acronyms. CPRD GOLD and CPRD Aurum contain primary care electronic health records (EHR) registered with general practices (GP) from a representative sample of 5% and 20% of UK inhabitants, respectively.[24–27] IPCI contains EHRs collected from patients registered with GPs in the Netherlands.[28] Norwegian linked health registry (NLHR@UiO) data includes secondary care and prescription information from both primary and secondary care.[29] IMASIS is an EHR hospital database containing records from patients being treated at the Hospital del Mar, Barcelona, Spain.[30] eDOL CHUM (further referred to as CHUM) is an EHR hospital database containing records from patients being treated at the University Hospital Montpellier, France.[31] CORIVA contains national health insurance claims from Estonia.[32] AUSOM is an EHR hospital database containing records from patients being treated at Ajou University Medical Centre, Suwon, Republic of Korea.[33] P+ contains medical and pharmacy claims from more than 107 million unique enrolees in largely commercial health plans.[34]

**Table 1.** Overview of participating databases.

| Database name | Acronym | Country | Data type | Healthcare setting | # people | Record end | COVID-19 negative test | Contribution |
| --- | --- | --- | --- | --- | --- | --- | --- | --- |
| Clinical Practice Research Datalink GOLD | CPRD GOLD | United Kingdom | EHR | Primary care | 21M | 06/2022 | Yes | IR, IRR |
| Clinical Practice Research Datalink Aurum | CPRD Aurum | United Kingdom | EHR | Primary care | 40M | 03/2021 | Yes | IR, IRR |
| The Integrated Primary Care Information | IPCI | The Netherlands | EHR | Primary care | 3M | 12/2022 | Yes | IR, IRR |
| Norwegian linked health registry data | NLHR@UIO | Norway | Administrative health related data | Primary care and secondary care | 5M | 12/2021 | No | IR |
| Hospital records from Parc Salut Mar Barcelona | IMASIS | Spain | EHR | Secondary care | 2M | 20/2022 | Yes | IR |
| Hospital records from the Montpellier University Hospital | CHUM | France | EHR | Secondary care | 2M | 12/2022 | Yes | IR |
| Hospital records from Ajou University Medical Centre | AUSOM | Korea | EHR | Secondary care | 2.7M | 02/2023 | Yes | IR |
| Healthcare claims from Estonia | CORIVA | Estonia | claims | Primary care and secondary care | 300K | 02/2022 | Yes | IR, IRR |
| PharMetrics® Plus for Academics | P+ | USA | claims | Primary care and secondary care | 107M | 06/2022 | No | IR |

EHR: electronic health records; IR: incidence rates; IRR: incidence rate ratios; M: million

**Table 2.** Patient characteristics stratified by database and cohort.

| **CPRD GOLD** | **General population** | **COVID-19** | **Test negative** |
| --- | --- | --- | --- |
| # patients | 3'597'966 | 352'178 | 669'155 |
| Female, n (%) | 1'809'553 (50%) | 191'578 (54%) | 371319 (55%) |
| median age (IQR) [years] | 41 (23 - 59) | 35 (19 - 51) | 38 (20 - 54) |
| prior history (IQR) [years] | 12 (4 - 18) | 14 (7 - 18) | 12 (5 - 18) |
| follow-up (IQR) [days] | 645 (519 - 646) | 233 (163 - 335) | 339 (243 - 522) |
| **CPRD AURUM** | **General population** | **COVID-19** | **Test negative** |
| # patients | 13'709'413 | 1'023'461 | 3'297'889 |
| Female, n (%) | 6'835'656 (50%) | 538'899 (53%) | 1'782'808 (54%) |
| median age (IQR) [years] | 38 (21 - 57) | 33 (17 - 49) | 35 (18 - 52) |
| prior history (IQR) [years] | 8 (3 - 19) | 11 (5 - 19) | 11 (5 - 19) |
| follow-up (IQR) [days] | 562 (552 - 563) | 246 (174 - 434) | 394 (240 - 475) |
| **IPCI** | **General population** | **COVID-19** | **Test negative** |
| # patients | 1'533'819 | 181'815 | 1'459 |
| Female, n (%) | 781'452 (51%) | 97'711 (54%) | 895 (61%) |
| median age (IQR) [years] | 42 (22 - 60) | 39 (21 - 55) | 58 (39 - 74) |
| prior history (IQR) [years] | 5 (1 - 8) | 7 (3 - 10) | 6 (4 - 7) |
| follow-up (IQR) [days] | 851 (517 - 851) | 352 (302 - 621) | 416 (323 - 489) |
| **NLHR@UIO** | **General population** | **COVID-19** | **Test negative** |
| # patients | 5'550'904 | 698'538 | NA |
| Female, n (%) | 2'746'672 (49%) | 349'625 (50%) | NA |
| median age (IQR) [years] | 39 (21 - 58) | 33 (19 - 50) | NA |
| prior history (IQR) [years] | 4 (3 - 9) | 6 (3 - 10) | NA |
| follow-up (IQR) [days] | 486 (486 - 486) | 371 (273 - 435) | NA |
| **IMASIS** | **General population** | **COVID-19** | **Test negative** |
| # patients | 166'450 | 13'939 | 27'703 |
| Female, n (%) | 86'523 (52%) | 7'300 (52%) | 15'088 (54%) |
| median age (IQR) [years] | 49 (33 - 67) | 58 (42 - 72) | 60 (41 - 75) |
| prior history (IQR) [years] | 12 (4 - 23) | 16 (8 - 26) | 17 (8 - 26) |
| follow-up (IQR) [days] | 645 (379 - 783) | 302 (153 - 488) | 345 (185 - 521) |
| **CHUM** | **General population** | **COVID-19** | **Test negative** |
| # patients | 1'910'472 | 6'121 | 73'187 |
| Female, n (%) | 98'8147 (52%) | 3'444 (56%) | 40'668 (56%) |
| median age (IQR) [years] | 42 (25 - 64) | 49 (30 - 71) | 43 (28 - 66) |
| prior history (IQR) [years] | 31 (26 - 31) | 31 (31 - 32) | 31 (29 - 32) |
| follow-up (IQR) [days] | 967 (967 - 967) | 572 (431 - 762) | 657 (471 - 825) |
| **AUSOM** | **General population** | **COVID-19** | **Test negative** |
| # records | 398'114 | 598 | 30'065 |
| # patients | 398'114 | 598 | 30'065 |
| Female, n (%) | 201403 (51%) | 321 (54%) | 15953 (53%) |
| median age (IQR) [years] | 49 (33 - 61) | 50 (29 - 68) | 55 (38 - 68) |
| prior history (IQR) [years] | 10 (4 - 17) | 12 (5 - 20) | 11 (5 - 19) |
| follow-up (IQR) [days] | 759 (477 - 904) | 235 (112 - 278) | 470 (307 - 620) |

| **CORIVA** | **General population** | **COVID-19** | **Test negative** |
| --- | --- | --- | --- |
| # records | 438'269 | 43'215 | 133'687 |
| # patients | 438'269 | 43'215 | 133'687 |
| Female, n (%) | 228'349 (52%) | 23'351 (54%) | 71'422 (53%) |
| median age (IQR) [years] | 41 (25 - 59) | 35 (20 - 51) | 39 (25 - 55) |
| prior history (IQR) [years] | 4 (4 - 4) | 4 (4 - 4) | 4 (4 - 4) |
| follow-up (IQR) [days] | 882 (882 - 882) | 722 (680 - 774) | 731 (620 - 811) |
| **P+** | **General population** | **COVID-19** | **Test negative** |
| # records | 8'257'952 | 201'947 | NA |
| # patients | 8'244'168 | 201'947 | NA |
| Female, n (%) | 4'275'098 (52%) | 105'983 (52%) | NA |
| median age (IQR) [years] | 47 (27 - 65) | 43 (27 - 59) | NA |
| prior history (IQR) [years] | 2 (1 - 4) | 4 (2 - 4) | NA |
| follow-up (IQR) [days] | 486 (211 - 576) | 318 (196 - 450) | NA |

CPRD GOLD: Clinical Practice Research Datalink GOLD; CPRD AURUM: Clinical Practice Research Datalink AURUM; IPCI: The Integrated Primary Care Information; NLHR@UiO: Norwegian linked health registry data; IMASIS: Hospital records from Parc Salut Mar Barcelona; CHUM: Hospital records from the Montpellier University Hospital; AUSOM: Hospital records from Ajou University Medical Centre; CORIVA: Healthcare claims from Estonia; P+: PharMetrics® Plus for Academics

### Study population

We included all patients with ≥365 days of history in the database (referred to as the general population). The study period lasted from the September 2020 until the latest data release available in each of the contributing databases (details are presented in Table 1). The index date was the first eligible day for each person.

From the general database population, we further defined two cohorts for comparative analyses: a COVID-19 cohort, whose index date was a first positive SARS-Cov-2 test or clinical COVID-19 diagnosis during the study period (referred to as COVID-19), and a comparator cohort, whose index date was a first negative SARS-Cov-2 test. For the COVID-19 and test negative cohort, patients with COVID-19 before the study period, with an influenza record within 3 months before the index date, with an outcome record within 6 months before the index date, and with <120 days of follow-up were excluded.

In the general population, we followed patients from day 1 until the outcome, data collection, death, or day 365. For comparative analyses, we followed eligible patients from day 91 after the index date to disregard the acute phase in COVID-19 and applied the same criteria for the test negative cohort. Follow-up end occurred at the first of the following: the outcome, end of data collection, death, day 365, or COVID-19 infection (test negative cohort only). Patients contributed a maximum of one episode each to the general population, test negative cohort, and COVID-19 cohort, if eligible.

### Outcomes

Our outcomes were a first record of POTS diagnoses and POTS symptoms such as palpitations, sinus tachycardia, vertigo, sweating, syncope, lightheadedness, orthostatic hypotension, dyspnoea, fatigue, dizziness, and giddiness. Moreover, we assessed a first record of ME/CFS diagnoses and ME/CFS symptoms such as fatigue, malaise, tiredness, irritable bowel syndrome, asthenia, dysthymia, heavy feeling, and heavy legs. Furthermore, we assessed a first record of multi-inflammatory syndrome (MIS) and the following autoimmune diseases: rheumatoid arthritis (RA), juvenile idiopathic arthritis (JIA), systemic lupus erythematosus (SLE), inflammatory bowel disease (IBD). We selected a first record of any diabetes mellitus (DM) as a control outcome to contextualize our results. Outcome code lists were established by TB and reviewed by DPA.

### Covariates

In comparative analyses, we performed stratified analyses by age (0 – 18 years, 19 – 64 years, >64 years) and sex. In the general population, we performed stratified analyses by healthcare setting (primary care, secondary care, both), calendar year, age (0-6 years, 7-11 years, 12-18 years, 19-40 years, 41-64 years, >64 years), and sex.

### Statistical analyses

Because of the many databases and different populations in this study, we kept the baseline characteristics to a minimum and reported frequency and percentage of the sexes, median and interquartile range for age, prior history, and follow-up.

To estimate the ratio of the outcome rates following COVID-19 versus the test negative cohort, we performed a meta-analysis of crude incidence rate ratios (IRR) with 95% confidence intervals (CI) for all outcomes individually using the R meta package[35]. We fed the *metainc* function the raw outcome and person-time estimates of exposed and comparator cohort and conducted a random effects meta-analysis. We replicated these analyses for every database individually to display database specific results.

In the general population, we estimated the disease burden of each outcome that yielded higher rates after COVID-19 than after negative testing (and additionally for MIS because of a clinical interest) by calculating incidence rate (IR) with 95% CIs. We stratified our results by healthcare setting to account for differences in data capture, by calendar year to see potential time trends, and by age groups and sex to determine who is most affected. Where frequency counts were less than five, data were obscured to further enhance patient/practice confidentiality, i.e. we have not reported IRR and IR estimates for these outcomes/strata. All analyses were carried out using R Version 4.2.3.0.

### Patient and public involvement

We engaged with the public and presented the study to a patient group at the Nuffield Department of Orthopaedics, Rheumatology, and Musculoskeletal Sciences (NDORMS) at the University of Oxford. Furthermore, we discussed the study setup with a patient representative of the European Alliance of Associations for Rheumatology (EULAR).

## RESULTS

Taking all the databases, we included a total of 34’549’575 individuals of whom 2’521’812 had COVID-19, and 4’233’145 a first negative test. CPRD Aurum was the biggest database with a total of 13’709’413 patients, of whom 1’023’461 had COVID-19 and 3’297’889 a negative test. Table 1 depicts patient characteristics of all databases. Among the general database populations, sex was balanced, and median age ranged from 38 years in CPRD Aurum to 49 years in both IMASIS and AUSOM. COVID-19 cohorts were younger than the general population in the primary care databases and NLHR@UiO, but older in all other secondary care databases. Test negative cohorts were again slightly older than COVID-19 cohorts in all instances but CHUM. Regarding sex, COVID-19 and test negative cohorts were well balanced expect for IPCI. In general, both COVID-19 and test negative cohorts contained more women than the general database populations.

Meta-analysed crude IRR after COVID-19 compared after negative testing depicted IRRs of 1.24 (1.23-1.25), 1.22 (1.21-1.23), and 1.12 (1.04-1.21) for POTS symptoms, ME/CFS symptoms and diagnoses, respectively (Figure 1, numeric values in Supplementary file 1). The results for symptoms were stronger in women than in men with IRRs of 1.23 (1.21-1.24) and 1.21 (1.2-1.23) for POTS symptoms and ME/CFS symptoms, respectively. Yet, the results for diagnoses were stronger for men with IRRs of 1.07 (1.01-1.12) for POTS diagnoses and of 1.42 (1.16-1.74) for ME/CFS diagnoses. The IRR of ME/CFS diagnoses was significant among the adult population aged 18-64 at 1.10 (1.02-1.20). IRRs of POTS symptoms and ME/CFS symptoms were highest in children at 1.41 (1.36-1.46) and 1.25 (1.21-1.30), respectively, followed those in elderly at 1.18 (1.16-1.21) and 1.16 (1.13-1.2), respectively.

**Figure 1.**
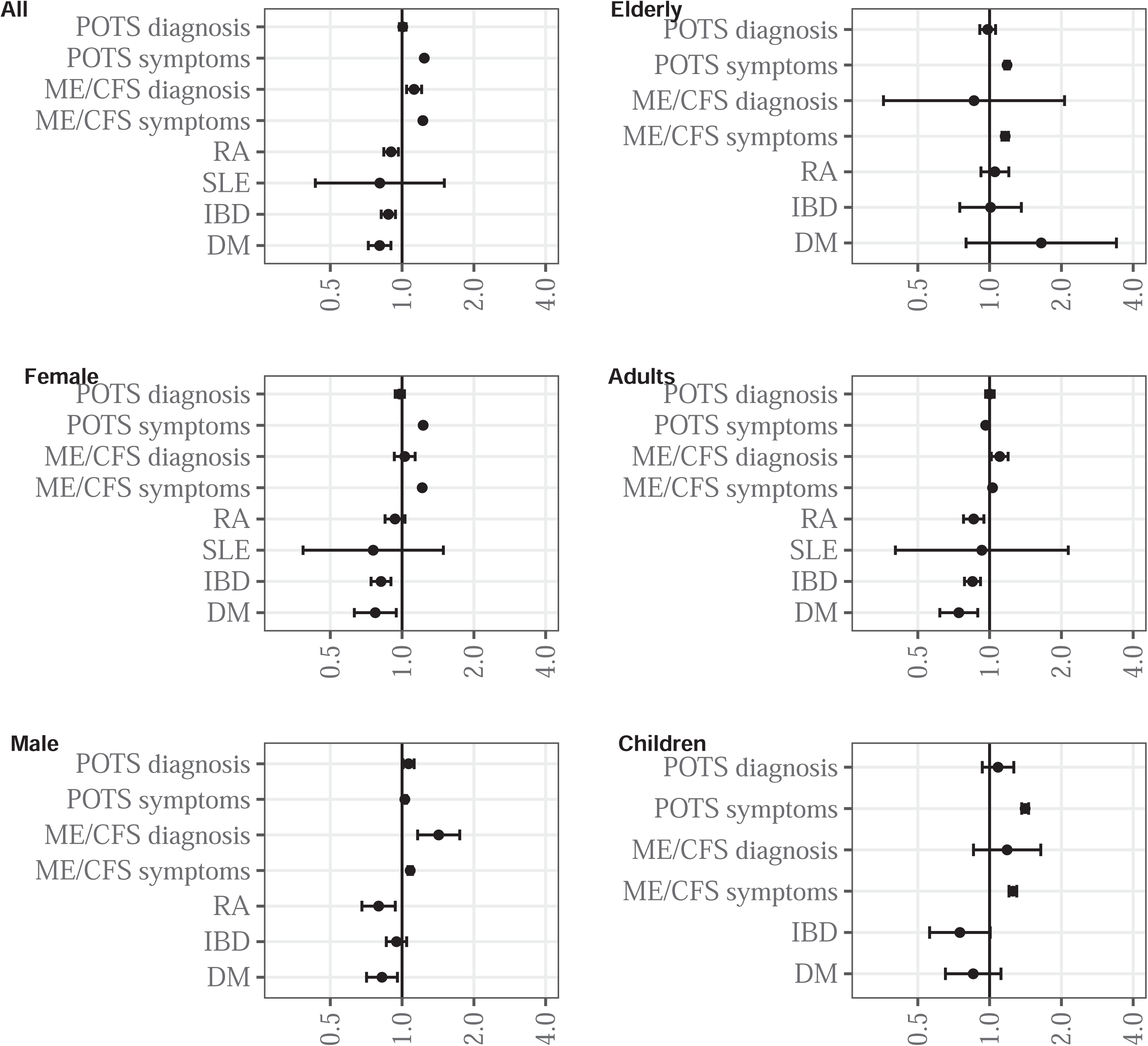
Meta-analysed crude incidence rate ratios with 95% confidence intervals for all outcomes among COVID-19 versus test negative patients, overall and stratified by age and sex. Outcomes not depicted because of too few counts: multi-inflammatory syndrome (MIS), juvenile idiopathic arthritis (JIA) IBD: inflammatory bowel disease; ME/CFS: myalgic encephalomyelitis / chronic fatigues syndrome; POTS: postural orthostatic tachycardia syndrome; RA: rheumatoid arthritis; SLE: systemic lupus erythematosus; DM: diabetes mellitus (control outcome)

For autoimmune diseases, IRRs after COVID-19 compared after negative testing of IBD and RA were 0.88 (0.82-0.94) and 0.90 (0.84-0.97), respectively. SLE yielded a null result, and JIA and MIS yielded too few counts for IRR analyses. The control outcome DM yielded an IRR of 0.81 (0.72-0.90). Input data for the meta-analyses of incidence rate ratios are depicted in Supplementary file 2. Results per database are presented in Supplementary file 3. IRR results were largely similar among individual databases though not all had sufficient counts for individual analyses.

In the general populations, we estimated the disease burden of POTS symptoms and diagnoses, ME/CFS symptoms and diagnoses, and MIS. Results are shown in Figures 2-5, with numeric values available in Supplementary files 4-7. The disease burden of each outcome was highest in databases containing primary and secondary care records, closely followed by primary care databases (for symptoms of POTS and ME/CFS). Secondary care data showed the biggest spread for diagnoses and usually included primary care estimates therein. IRs of symptoms of POTS and ME/CFS were 40-7’760/100’000 pys and 19-6’022/100’000 pys, respectively, those of diagnoses were 17-1’477/100’000 pys and 2-473/100’000 pys, respectively. Disease burden of MIS was lowest with IRs 0.4-16/100’000 pys. IRs of DM as a benchmark were 8-86/100’000 pys.

**Figure 2.**
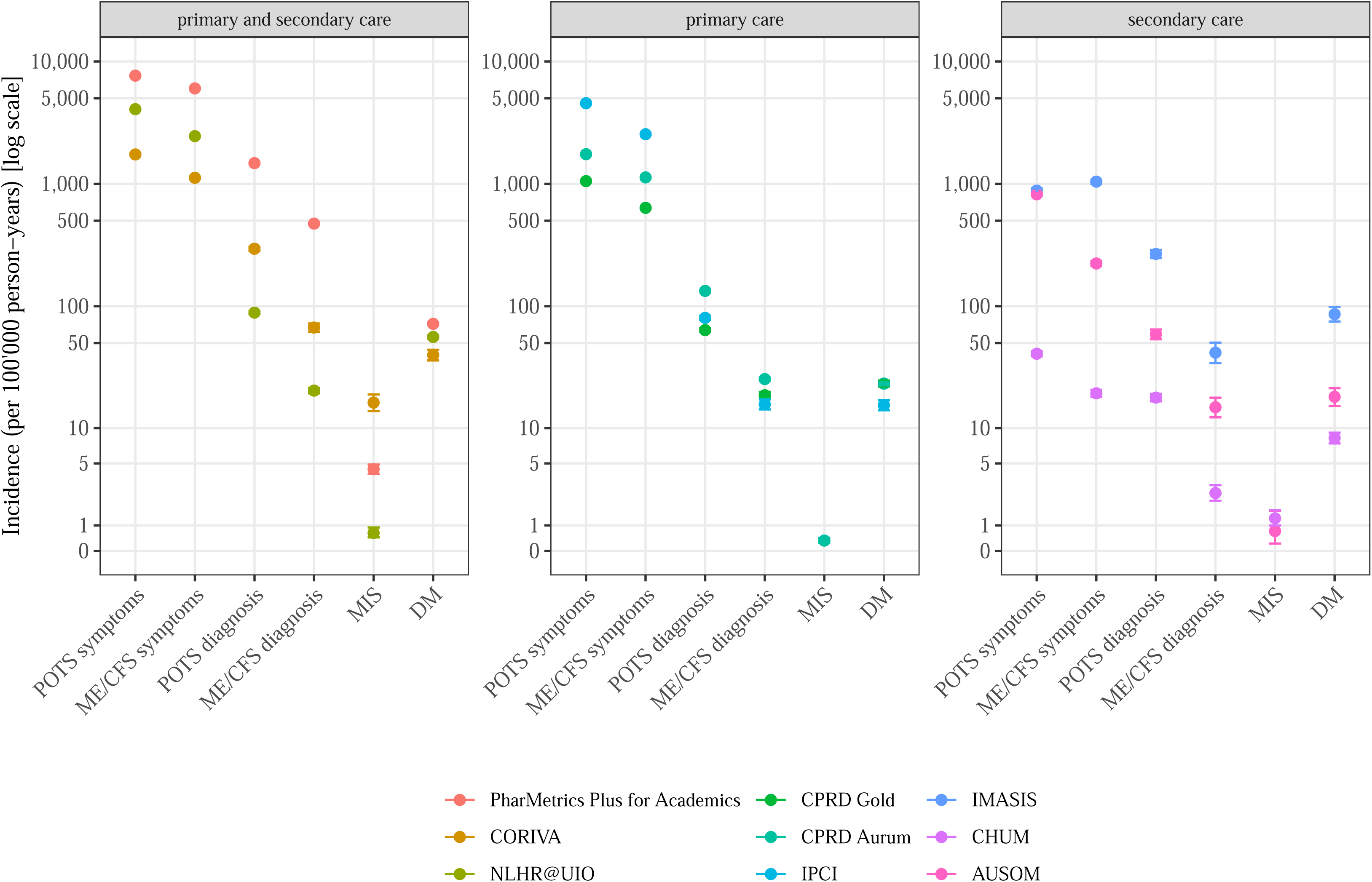
Incidence rates with 95% confidence intervals for all outcomes stratified by care setting. IBD: inflammatory bowel disease; JIA: juvenile idiopathic arthritis; ME/CFS: myalgic encephalomyelitis / chronic fatigues syndrome; MIS: multi-inflammatory syndrome, POTS: postural orthostatic tachycardia syndrome; RA: rheumatoid arthritis; SLE: systemic lupus erythematosus; DM: diabetes mellitus (control outcome)

Time trends depicted upwards trends for all assessed outcomes of interest, which was most profound in secondary care databases (Figure 3, Supplementary file 5). DM as a reference depicted a less profound upward trend and even a slight downwards trend in primary care databases.

**Figure 3.**
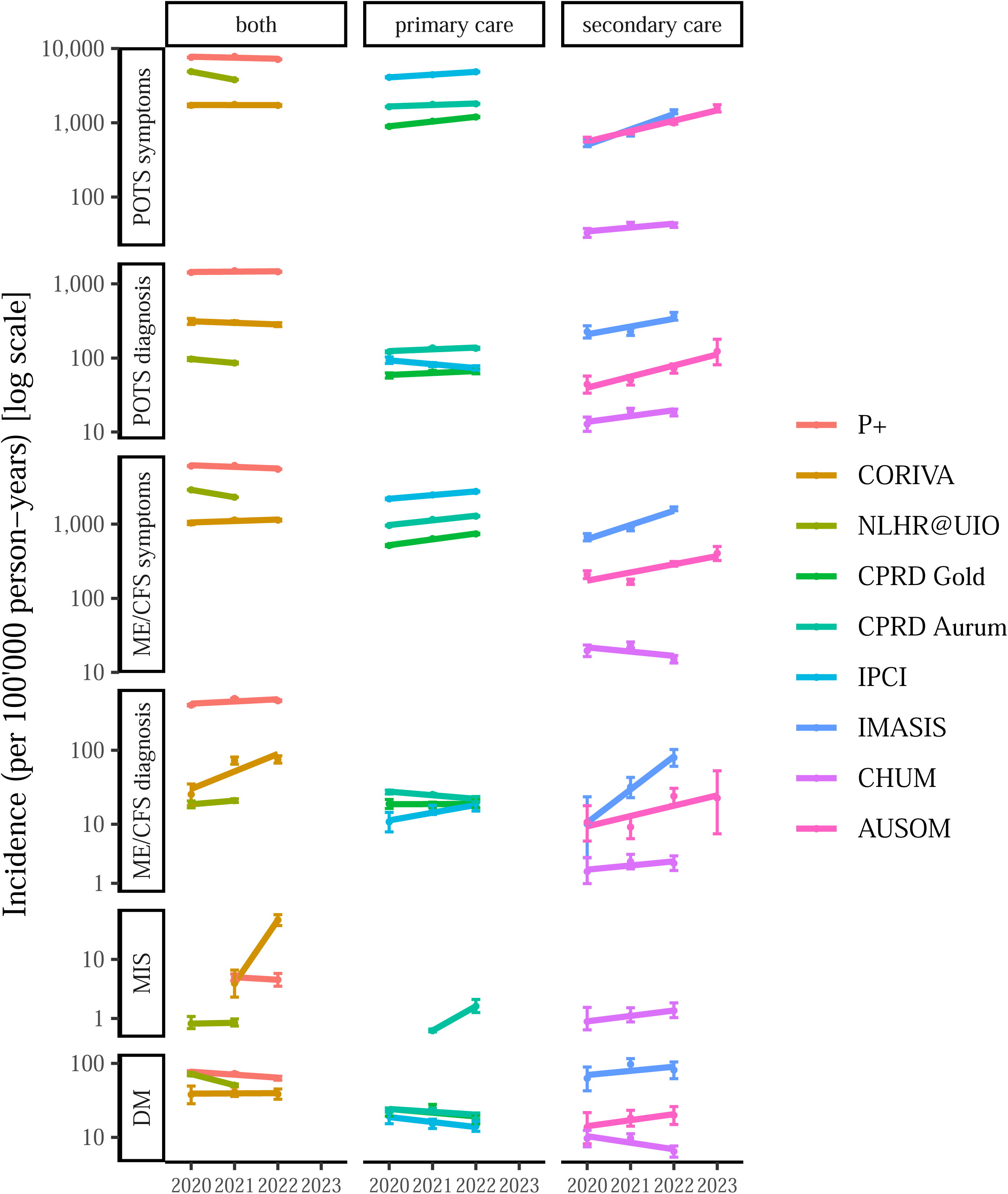
Incidence rates with 95% confidence intervals for all outcomes stratified by care setting and calendar year. IBD: inflammatory bowel disease; JIA: juvenile idiopathic arthritis; ME/CFS: myalgic encephalomyelitis / chronic fatigues syndrome; MIS: multi-inflammatory syndrome, POTS: postural orthostatic tachycardia syndrome; RA: rheumatoid arthritis; SLE: systemic lupus erythematosus; DM: diabetes mellitus (control outcome)

In age stratified analyses, IRs of POTS symptoms and diagnoses tended to increase with age (Figure 4, Supplementary file 6). For example, the age groups 19-64 years and >64 years yielded IRs of POTS diagnoses of 17-1’282/100’000 pys and 37-2’643/100’000 pys. IRs of ME/CFS symptoms plateaued >19 years and IRs of ME/CFS diagnoses followed a reversed U-shape-with highest IRs for 19-64-year-olds. IRs among children were less high at 4-609/100’000 pys and 1-153/100’000 pys for POTS and ME/CFS diagnoses, respectively.

**Figure 4.**
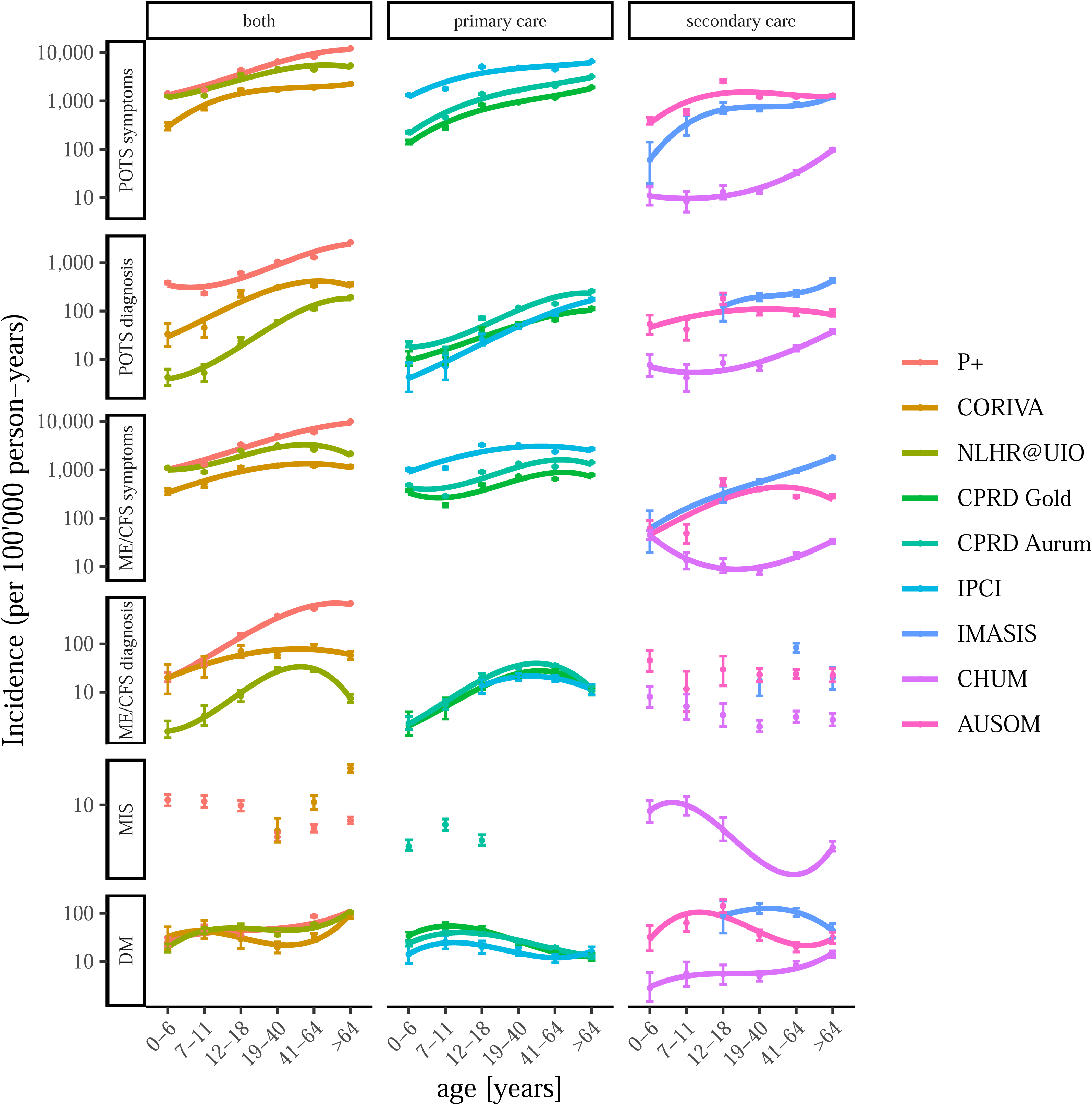
Incidence rates with 95% confidence intervals for all outcomes stratified by care setting and age. IBD: inflammatory bowel disease; JIA: juvenile idiopathic arthritis; ME/CFS: myalgic encephalomyelitis / chronic fatigues syndrome; MIS: multi-inflammatory syndrome, POTS: postural orthostatic tachycardia syndrome; RA: rheumatoid arthritis; SLE: systemic lupus erythematosus; DM: diabetes mellitus (control outcome)

Furthermore, a difference in IRs between MIS-A and MIS-C was observed. IRs of MIS-C ranged from 5-13/100’000 pys in databases contributing at least secondary care (IRs in primary care were much lower). MIS-A was captured mainly in Estonia and the US (contributing both primary and secondary care) and depicted an increasing trend with age with IRs of 2-3/100’000 pys among 19-40-year-olds to 5-58/100’000 pys for those aged >64 years.

In sex-stratified analyses, POTS and ME/CFS diagnoses and symptoms were more frequent in women than in men, while symptoms yielded more profound difference between sexes than did diagnoses (Figure 5, Supplementary file 7). MIS yielded mixed results and T2DM was more frequent in men.

**Figure 5.**
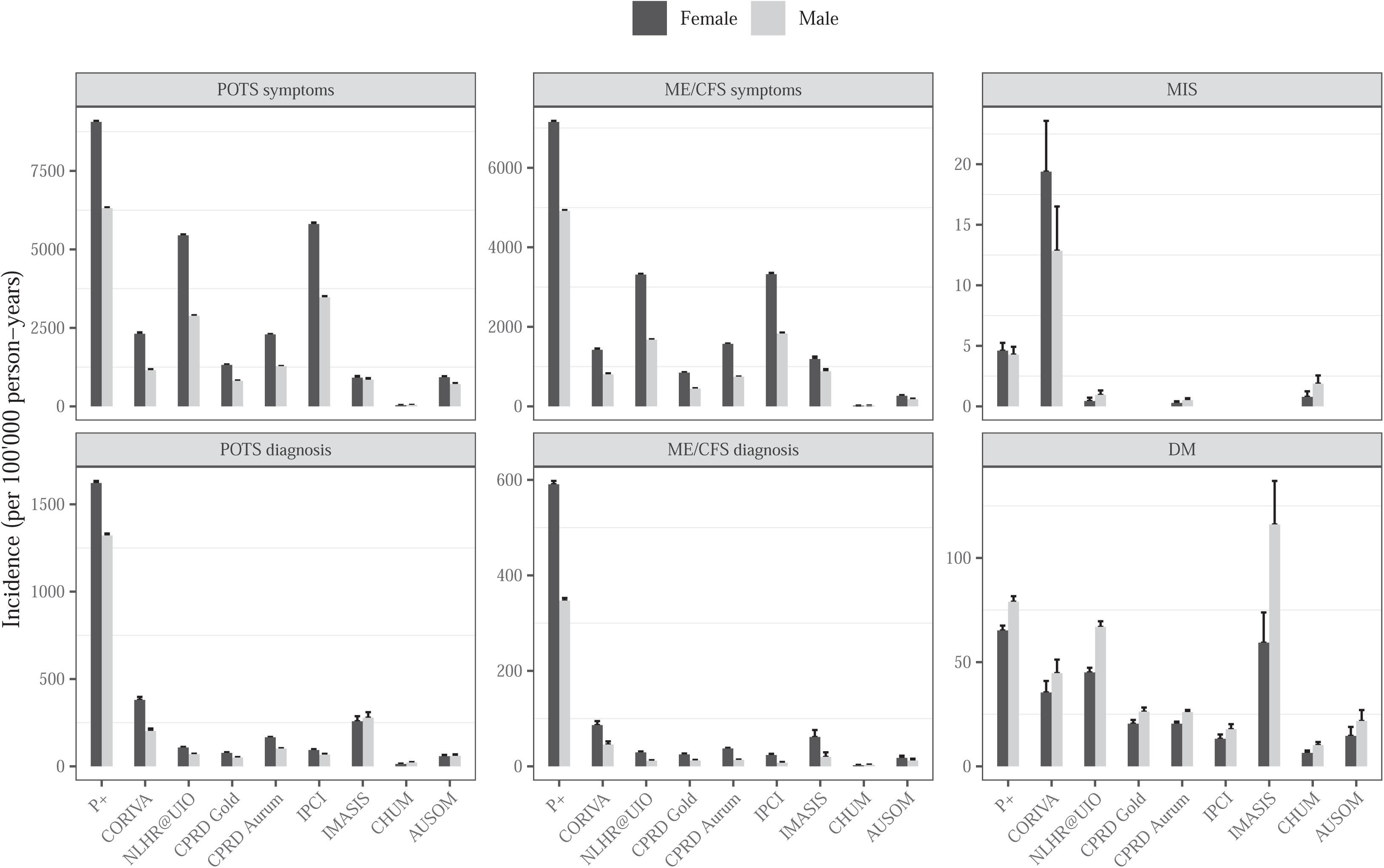
Incidence rates with 95% confidence intervals for all outcomes stratified by care setting and sex. IBD: inflammatory bowel disease; JIA: juvenile idiopathic arthritis; ME/CFS: myalgic encephalomyelitis / chronic fatigues syndrome; MIS: multi-inflammatory syndrome, POTS: postural orthostatic tachycardia syndrome; RA: rheumatoid arthritis; SLE: systemic lupus erythematosus; DM: diabetes mellitus (control outcome)

## DISCUSSION

In this large network study among around 35 million individuals from nine databases in Europe, the US and Korea, of whom 3 million patients had COVID-19 and 4 million patients a negative test, POTS symptoms, ME/CFS symptoms, and ME/CFS diagnoses after COVID-19 yielded 1.24, 1.22, and 1.12 times the rates compared to after negative testing. Among the working age group, the rates of ME/CFS diagnoses were increased, while among children and elderly, the rates of symptoms of ME/CFS and POTS were increased. The disease burden of POTS and ME/CFS diagnoses in the general population were higher among women than among men and overall similar to that of diabetes mellitus.

Our result suggesting that the rate of POTS symptoms after COVID-19 compared to after negative testing is 1.24 times increased is conservative compared to literature. A systematic review and meta-analysis observed a 2-fold increased rate of POTS in infected versus uninfected.[36] A reason for the difference can be different definitions of comparators and/or outcome. While we observed an increased rate for POTS symptoms, we observed a null result for POTS diagnoses. Yet, POTS has only gained attention recently and recording for patients may differ between care settings, countries, and databases. Therefore, POTS symptoms should be used as a proxy for POTS diagnoses.

Concerning ME/CFS, we observed that after COVID-19, the rates for ME/CFS diagnoses and symptoms were 1.12 and 1.22 times higher compared to after negative testing, respectively. There is no other literature that has estimated ME/CFS risks or rates yet. However, a systematic review and meta-analysis compared ME/CFS incidence following different virus infections (not including COVID-19) when compared to healthy individuals and observed increased odds of at least 2-fold.[37] Thus, our results would fit nicely with previously observed viral infection-related triggers and may suggest that SARS-Cov-2 also increases the absolute risks of ME/CFS. This suggestion is consistent with narrative reviews suggesting how SARS-Cov-2 may lead to ME/CFS.[38,39] Furthermore, given observed increased risks/rates following COVID-19, also our observed upward trend in rates of POTS and ME/CFS over time aligns with the increasing spread of the pandemic with time.

Furthermore, our results include age- and sex-stratified analyses while no other study has yet assessed the influence of age and sex on the relative risks/rates of POTS and ME/CFS following COVID-19. We observed that POTS/ME/CFS symptom results were mainly driven by women (around 0.2 rate increase), children (0.3-0.4 rate increase), and elderly (around 0.2 rate increase), POTS/ME/CFS diagnoses results by men (0.1-0.4 rate increase) and adults (0.1 rate increase for ME/CFS only). A study reviewing post-acute COVID-19 conditions in paediatric populations concluded the following: “Frequent symptoms include fatigue, exertion intolerance, and anxiety. Some patients present with postural tachycardia syndrome (PoTS), and a small number of cases fulfill the clinical criteria of myalgic encephalomyelitis/chronic fatigue syndrome (ME/CFS).”[40] Thus, our findings are consistent with the suggested existence of POTS and ME/CFS as post-acute COVID-19 conditions in paediatrics. In addition, our findings suggest absolute risks in paediatric populations of POTS and ME/CFS of 4-609/100’000 pys and 1-153/100’000 pys, respectively. A study undertaken in a post-COVID-19 clinic among 140 patients also suggests that women are more affected by ME/CFS and reports median age of 47 years which is consistent with our U-shaped findings of highest IRs among 19-64-year olds.[41] Moreover, a meta-analysis including studies from Japan and Korea between 1994 and January 2021 suggest that the prevalence of ME/CFS was around 0.8% and that women were more affected.[42] If we were to estimate the cumulative incidence from our results, we would yield 0.002-0.473% and prevalence would rise quickly given that ME/CFS is uncurable to date. Moreover, estimates would be around 10-fold higher for ME/CFS symptoms and up to 5% of a population seeking healthcare anew because of ME/CFS related symptoms is substantial. Most studies to date assessed post-acute COVID-19 conditions or symptoms among the infected only.[36,43] However, our work contributing absolute risks of ME/CFS and POTS among the general population is important to inform the healthcare system about the existing disease burden and to allocate the needed resources for these long-term conditions. Moreover, to put our estimated rates into perspective, we estimated the incidence of DM and observed similar incidence with POTS and ME/CFS, especially among children and elderly for POTS and for working age for ME/CFS. Our results suggest a significant strain for individuals, society, and the healthcare systems.

For autoimmune diseases and DM, we observed that after COVID-19 the rates were 0.1-0.2 times lower than after negative testing. This finding contrasts with existing literature mainly publishing increased risks of RA, IBD, and other autoimmune diseases. [18–20] There has also been reports of glucometabolic control to fail during and after COVID-19, yet it remains to be seen whether these symptoms persist.[44,45] We suggest the observed decreased rates after COVID-19 to be due to symptoms being non-specific and may have been associated with aftermaths of COVID-19 thus leading to delayed diagnoses.

Our study has the strengths of producing a large quantity of standardised results from various countries and care settings in Europe. Thus, our results are widely generalisable to primary and secondary care settings in Europe, but probably only generalisable to patients similar to that of P+ in the US and Ajou hospital in Korea. Yet, the results were largely similar among the individual databases, which underlines the robustness of our findings. Moreover, we used test negative comparators which are more reliable than comparing to patients without COVID-19 because they may have still been infected at the time. However, we must consider several limitations, especially for comparative analyses. We did not match our comparison groups and did not adjust for confounders leaving our results crude. Yet, we found age and sex mostly balanced. Moreover, our results may have been subject to bias through differential recording of post-acute COVID-19 conditions and symptoms over time especially with increasing awareness by healthcare personnel.

## CONCLUSION

In our large but unmatched comparisons, we observed that, following COVID-19, POTS and ME/CFS yielded higher rates than after negative testing. In absolute terms, we observed POTS and ME/CFS diagnoses to have a similar disease burden as DM which may imply a significant strain on the healthcare system. Well matched and large comparison analyses are needed to assess this matter. Furthermore, in contrast to current literature, we did not observe increased rates of auto-immune diseases or DM following COVID-19.

## Data Availability

Patient level data cannot be shared without approval from data custodians owing to local information governance and data protection regulations. The analytical code is available at https://github.com/oxford-pharmacoepi/LongCovidStudyathon_W1. The code for postprocessing and aggregated results are available at https://github.com/tiozab/immune_inflammatory_PACS/

## ACKNOWLEDGEMENTS

We would like to thank Mahkameh Mafi and Francesca Walker for their help in organising the studyathon that brought together all collaborators for a week in Oxford in spring 2023.

## Details of any meeting at which the work has been / will be presented

“The risks of immune- and inflammatory related post-acute sequelae of COVID-19: a network study in 6 European countries, the US, and Korea” (International Conference on *Pharmacoepidemiology* & Therapeutic Risk Management [ICPE] in Berlin, Germany, 24-28 August 2024)

“The risks of immune- and inflammatory related post-acute sequelae of COVID-19 (PASC): a network study in 6 European countries, the US, and Korea” (OHDSI Europe symposium in Rotterdam, The Netherlands, 3 June 2024)

## CONTRIBUTORS

DPA and AMJ obtained the funding for this project.

KLG, MC, DPA, JX and AMJ led the conceptualisation of the study.

KK, DPA, and AMJ led the phenotyping of COVID-19 and test negative patients

TB led the phenotyping of outcomes

AD mapped and curated CPRD data

KLG and MC wrote the analytical code

TB wrote the postprocessing code

KLG, MC, DDed, RK, JMR, MM, DDel, CK, JK, and NTHT conducted the statistical analyses on the respective databases

TB, KLG, SK, AU, DDed, JOO, MAM, TDS, JTA, AA, KK, NMB, LM, CL, RP, JMC, NTHT, HMEN, CK, JK, GM, EB, JA, LPC, RLP, BV, AM, AN, AMJ, DPA, JX clinically interpreted the results

TB wrote the first draft of the paper

All authors read, contributed to, and approved the last version of the paper.

## ETHICAL APPROVAL

The protocol for this research was approved by the independent scientific advisory committee for Medicine and Healthcare products Regulatory Agency database research (protocol number 23_002603). Informed consent of individual patients was not required as anonymised information was obtained from medical records. Ethical approval for the Norwegian data in this study was obtained from The Regional Committee for Research Ethics (approval number 155294) and the Data Protection Officer at the University of Oslo (approval number 523275). Ethical approval for CORIVA data was obtained from the Research Ethics Committee of the University of Tartu (No. 351/M-8). Ethical approval for IMASIS was obtained by the Parc de Salut Mar Research Ethics Committee CEIm-Parc de Salut Mar (number 2021/9975). Ethical approval for IPCI was obtained by the Integrated Primary Care Information review board (registration number 9/2023). For CHUM, no ethical approval was required according to French law for this study. All patients admitted to the hospital are provided with general information about the collection and secondary use of their data, and an opt-out option is offered. PharMetrics® Plus for Academics needed no approval for use of pseudoanonymised secondary data. Ethical approval for AUSOM was obtained by the Ajou University Medical Center Institutional Review Board (No. AJOUIRB-MDB-2021-694).

## DATA SHARING

Patient level data cannot be shared without approval from data custodians owing to local information governance and data protection regulations. The analytical code is available at <u>oxford-pharmacoepi/LongCovidStudyathon_W1 (github.com)</u>. The code for postprocessing and aggregated results are available at <u>immune_inflammatory_PACS/ at maintiozab/immune_inflammatory_PACS (github.com)</u>.

## TRANSPARENCY STATEMENT

The lead author (TB) affirms that this manuscript is an honest, accurate, and transparent account of the study being reported; that no important aspects of the study have been omitted; and that any discrepancies from the study as planned (and, if relevant, registered) have been explained.

## REFERENCES

1 Astin R, Banerjee A, Baker MR, et al. Long COVID: mechanisms, risk factors and recovery. Exp Physiol. 2023;108:12–27. 10.1113/EP090802

2 Lippi G, Sanchis-Gomar F, Henry BM. COVID-19 and its long-term sequelae: what do we know in 2023? Pol Arch Intern Med. 2023;133. 10.20452/pamw.16402

3 Lopez-Leon S, Wegman-Ostrosky T, Perelman C, et al. More than 50 long-term effects of COVID-19: a systematic review and meta-analysis. Sci Rep. 2021;11. doi: 10.1038/s41598-021-95565-8

4 Vos T, Hanson SW, Abbafati C, et al. Estimated Global Proportions of Individuals with Persistent Fatigue, Cognitive, and Respiratory Symptom Clusters Following Symptomatic COVID-19 in 2020 and 2021. JAMA. 2022;328:1604–15.

5 Chen C, Haupert SR, Zimmermann L, et al. Global Prevalence of Post COVID-19 Condition or Long COVID: A Meta-Analysis and Systematic Review 2 Running Title: Post COVID-19 Condition Meta-Analysis 3 4. J Infect Dis. 2022;226:1593–607.

6 Ormiston CK, Świątkiewicz I, Taub PR. Postural orthostatic tachycardia syndrome as a sequela of COVID-19. Heart Rhythm. 2022;19:1880–9.

7 Kwan AC, Ebinger JE, Wei J, et al. Apparent risks of postural orthostatic tachycardia syndrome diagnoses after COVID-19 vaccination and SARS-Cov-2 Infection. Nature Cardiovascular Research. 2022;1:1187–94.

8 Seeley MC, Gallagher C, Ong E, et al. High Incidence of Autonomic Dysfunction and Postural Orthostatic Tachycardia Syndrome in Patients with Long COVID: Implications for Management and Health Care Planning. American Journal of Medicine. Published Online First: 2023. doi: 10.1016/j.amjmed.2023.06.010

9 Chadda KR, Blakey EE, Huang CLH, et al. Long COVID-19 and Postural Orthostatic Tachycardia SyndromeIs Dysautonomia to Be Blamed? Front Cardiovasc Med. 2022;9. doi: 10.3389/fcvm.2022.860198

10 Gómez-Moyano E, Rodríguez-Capitán J, Gaitán Román D, et al. Postural orthostatic tachycardia syndrome and other related dysautonomic disorders after SARS-CoV-2 infection and after COVID-19 messenger RNA vaccination. Front Neurol. 2023;14. 10.3389/fneur.2023.1221518

11 Mallick D, Goyal L, Chourasia P, et al. COVID-19 Induced Postural Orthostatic Tachycardia Syndrome (POTS): A Review. Cureus. Published Online First: 31 March 2023. doi: 10.7759/cureus.36955

12 Malkova AM, Shoenfeld Y. Autoimmune autonomic nervous system imbalance and conditions: Chronic fatigue syndrome, fibromyalgia, silicone breast implants, COVID and post-COVID syndrome, sick building syndrome, post-orthostatic tachycardia syndrome, autoimmune diseases and autoimmune/inflammatory syndrome induced by adjuvants. Autoimmun Rev. 2023;22. 10.1016/j.autrev.2022.103230

13 Komaroff AL, Lipkin WI. ME/CFS and Long COVID share similar symptoms and biological abnormalities: road map to the literature. Front Med (Lausanne). 2023;10. 10.3389/fmed.2023.1187163

14 Morrow AK, Malone LA, Kokorelis C, et al. Long-Term COVID 19 Sequelae in Adolescents: the Overlap with Orthostatic Intolerance and ME/CFS. Curr Pediatr Rep. 2022;10:31–44. 10.1007/s40124-022-00261-4

15 Tate W, Walker M, Sweetman E, et al. Molecular Mechanisms of Neuroinflammation in ME/CFS and Long COVID to Sustain Disease and Promote Relapses. Front Neurol. 2022;13. doi: 10.3389/fneur.2022.877772

16 Qamar MA, Afzal SS, Dhillon RA, et al. A global systematic review and meta-analysis on the emerging evidence on risk factors, clinical characteristics, and prognosis of multisystem inflammatory syndrome in adults (MIS-A). Annals of Medicine & Surgery. 2023;85:4463–75.

17 Yazdanpanah N, Rezaei N. Autoimmune complications of COVID-19. J Med Virol. 2022;94:54– 62. 10.1002/jmv.27292

18 Marín JS, Mazenett-Granados EA, Salazar-Uribe JC, et al. Increased incidence of rheumatoid arthritis after COVID-19. Autoimmun Rev. 2023;22. 10.1016/j.autrev.2023.103409

19 Tesch F, Ehm F, Vivirito A, et al. Incident autoimmune diseases in association with SARS-CoV-2 infection: a matched cohort study. Clin Rheumatol. Published Online First: 2023. doi: 10.1007/s10067-023-06670-0

20 Chang R, Yen-Ting Chen T, Wang SI, et al. Risk of autoimmune diseases in patients with COVID-19: A retrospective cohort study. EClinicalMedicine. 2023;56. doi: 10.1016/j.eclinm.2022.101783

21 Voss EA, Makadia R, Matcho A, et al. Feasibility and utility of applications of the common data model to multiple, disparate observational health databases. Journal of the American Medical Informatics Association. 2015;22:553–64.

22 Marc Overhage J, Ryan PB, Reich CG, et al. Validation of a common data model for active safety surveillance research. Journal of the American Medical Informatics Association. 2012;19:54–60.

23 Hripcsak G, Duke JD, Shah NH, et al. Observational Health Data Sciences and Informatics (OHDSI): Opportunities for Observational Researchers. Stud Health Technol Inform. 2015;216:574–8.

24 Herrett E, Gallagher AM, Bhaskaran K, et al. Data Resource Profile: Clinical Practice Research Datalink (CPRD). Int J Epidemiol. 2015;44:827–36.

25 Wolf A, Dedman D, Campbell J, et al. Data resource profile: Clinical Practice Research Datalink (CPRD) Aurum. Int J Epidemiol. 2019;1–8.

26 Syder, Kirsty. Release Notes for CPRD GOLD. doi: 10.48329/ty3h-h728

27 Syder, Kirsty. Release Notes for CPRD Aurum. doi: 10.48329/8pm6-4q84

28 De Ridder MAJ, De Wilde M, De Ben C, et al. Data Resource Profile: The Integrated Primary Care Information (IPCI) database, The Netherlands. Int J Epidemiol. 2022;51:E314–23.

29 Bakken IJ, Ariansen AMS, Knudsen GP, et al. The Norwegian Patient Registry and the Norwegian Registry for Primary Health Care: Research potential of two nationwide health-care registries. Scand J Public Health. 2020;48:49–55. 10.1177/1403494819859737

30 Mayer MA, Furlong LI, Torre P, et al. Reuse of EHRs to Support Clinical Research in a Hospital of Reference. Studies in Health Technology and Informatics. IOS Press 2015:224–6. 10.3233/978-1-61499-512-8-224

31 https://www.chu-montpellier.fr/fr/plateformes-recherche/eds. Entrepôt de Données du Languedoc - Le Centre Hospitalier Universitaire du CHU de Montpellier. Entrepôt de Données du Languedoc - Le Centre Hospitalier Universitaire du CHU de Montpellier (accessed 15 November 2023)

32 Oja M, Tamm S, Mooses K, et al. Transforming Estonian health data to the Observational Medical Outcomes Partnership (OMOP) Common Data Model: lessons learned. JAMIA Open. 2023;6. doi: 10.1093/jamiaopen/ooad100

33 Yoon D, Ahn EK, Park MY, et al. Conversion and data quality assessment of electronic health record data at a korean tertiary teaching hospital to a common data model for distributed network research. Healthc Inform Res. 2016;22:54–8.

34 IQVIA. IQVIA PharMetrics ® Plus for Academics. 2022.

35 https://cran.r-project.org/web/packages/meta/index.html.

36 Yong SJ, Halim A, Liu S, et al. Pooled rates and demographics of POTS following SARS-CoV-2 infection versus COVID-19 vaccination: Systematic review and meta-analysis. Auton Neurosci. 2023;250:103132.

37 Hwang JH, Lee JS, Oh HM, et al. Evaluation of viral infection as an etiology of ME/CFS: a systematic review and meta-analysis. J Transl Med. 2023;21. 10.1186/s12967-023-04635-0

38 Komaroff AL, Lipkin WI. Insights from myalgic encephalomyelitis/chronic fatigue syndrome may help unravel the pathogenesis of postacute COVID-19 syndrome. Trends Mol Med. 2021;27:895–906. 10.1016/j.molmed.2021.06.002

39 Choutka J, Jansari V, Hornig M, et al. Unexplained post-acute infection syndromes. Nat Med. 2022;28:911–23. 10.1038/s41591-022-01810-6

40 Toepfner N, Brinkmann F, Augustin S, et al. Long COVID in pediatrics—epidemiology, diagnosis, and management. Eur J Pediatr. Published Online First: 27 January 2024. doi: 10.1007/s00431-023-05360-y

41 Bonilla H, Quach TC, Tiwari A, et al. Myalgic Encephalomyelitis/Chronic Fatigue Syndrome is common in post-acute sequelae of SARS-CoV-2 infection (PASC): Results from a post-COVID-19 multidisciplinary clinic. Front Neurol. 2023;14. doi: 10.3389/fneur.2023.1090747

42 Lim EJ, Son CG. Prevalence of chronic fatigue syndrome (Cfs) in korea and japan: A meta-analysis. J Clin Med. 2021;10. 10.3390/jcm10153204

43 Ceban F, Ling S, Lui LMW, et al. Fatigue and cognitive impairment in Post-COVID-19 Syndrome: A systematic review and meta-analysis. Brain Behav Immun. 2022;101:93–135. 10.1016/j.bbi.2021.12.020

44 Lai H, Yang M, Sun M, et al. Risk of incident diabetes after COVID-19 infection: A systematic review and meta-analysis. Metabolism. 2022;137. 10.1016/j.metabol.2022.155330

45 Montefusco L, Ben Nasr M, D’Addio F, et al. Acute and long-term disruption of glycometabolic control after SARS-CoV-2 infection. Nat Metab. 2021;3:774–85.

