## Supplementary File 1 for "The risks of autoimmune- and inflammatory post-acute COVID-19 conditions: a network cohort study in six European countries, the US, and Korea"

### Supplementary File 1. Numeric values of incidence rate ratios with 95% confidence intervals corresponding to Figure 1

###### Supplementary Table 1. Numeric values of incidence rate ratios with 95% confidence intervals corresponding to Figure 1

|  | **All** | **Female** | **Male** | **Elderly**  **(>64 years** | **Adults**  **(19-64 years)** | **Children**  **(<19 years)** |
| --- | --- | --- | --- | --- | --- | --- |
| **POTS symptoms** | 1.24 (1.23-1.25) | 1.23 (1.21-1.24) | 1.03 (1.01-1.04) | 1.18 (1.16-1.21) | 0.96 (0.95-0.97) | 1.41 (1.36-1.46) |
| **ME/CFS symptoms** | 1.22 (1.21-1.23) | 1.21 (1.2-1.23) | 1.08 (1.06-1.1) | 1.16 (1.13-1.2) | 1.03 (1.01-1.04) | 1.25 (1.21-1.3) |
| **ME/CFS diagnosis** | 1.12 (1.04-1.21) | 1.03 (0.93-1.13) | 1.42 (1.16-1.74) | 0.86 (0.36-2.06) | 1.1 (1.02-1.2) | 1.18 (0.86-1.64) |
| **POTS diagnosis** | 1.01 (0.97-1.04) | 0.98 (0.94-1.02) | 1.07 (1.01-1.12) | 0.98 (0.91-1.06) | 1 (0.96-1.05) | 1.08 (0.93-1.26) |
| **RA** | 0.9 (0.84-0.97) | 0.93 (0.85-1.03) | 0.8 (0.68-0.94) | 1.05 (0.92-1.2) | 0.86 (0.78-0.95) | NA |
| **IBD** | 0.88 (0.82-0.94) | 0.82 (0.74-0.9) | 0.95 (0.86-1.05) | 1.01 (0.75-1.36) | 0.85 (0.78-0.92) | 0.75 (0.56-1.01) |
| **SLE** | 0.81 (0.43-1.5) | 0.76 (0.38-1.49) | NA | NA | 0.93 (0.4-2.14) | NA |
| **DM**  **(control outcome)** | 0.81 (0.72-0.9) | 0.77 (0.63-0.95) | 0.82 (0.71-0.96) | 1.65 (0.8-3.4) | 0.74 (0.62-0.89) | 0.85 (0.65-1.12) |

IBD: inflammatory bowel disease; ME/CFS: myalgic encephalomyelitis / chronic fatigues syndrome; NA: results suppressed because less than 5 outcomes; POTS: postural orthostatic tachycardia syndrome; RA: rheumatoid arthritis; SLE: systemic lupus erythematosus; DM: diabetes mellitus
