## Supplementary File 2 for "The risks of autoimmune- and inflammatory post-acute COVID-19 conditions: a network cohort study in six European countries, the US, and Korea"

### Supplementary File 2. Input data for the meta-analyses of incidence rate ratios

###### Supplementary Table 2. Input data for the meta-analyses of incidence rate ratios

| **Person-years exposed** | **# events exposed** | **Person-years unexposed** | **# events unexposed** | **database** | **Age group [years]** | **sex** | **outcome** |
| --- | --- | --- | --- | --- | --- | --- | --- |
| 147246.9 | 55 | 364968.7 | 204 | CPRDGOLD | 0 to 150 | Both | IBD |
| 7746.223 | <5 | 16399.58 | 17 | IMASIS | 0 to 150 | Both | IBD |
| 0 | 0 | 20795.69 | 6 | AUSOM | 0 to 150 | Both | IBD |
| 114135.9 | 54 | 954.412 | <5 | IPCI | 0 to 150 | Both | IBD |
| 0 | 0 | 49179.63 | 24 | eDOL_CHUM | 0 to 150 | Both | IBD |
| 26838.89 | 20 | 75875.79 | 59 | CORIVA | 0 to 150 | Both | IBD |
| 475687.8 | 265 | 1822509 | 1089 | CPRDAurum | 0 to 150 | Both | IBD |
| 79645.83 | 33 | 203377.1 | 110 | CPRDGOLD | 0 to 150 | Female | IBD |
| 4065.352 | <5 | 9027.14 | 8 | IMASIS | 0 to 150 | Female | IBD |
| 0 | 0 | 11076.88 | <5 | AUSOM | 0 to 150 | Female | IBD |
| 60802.08 | 34 | 589.2293 | <5 | IPCI | 0 to 150 | Female | IBD |
| 0 | 0 | 27530.88 | 10 | eDOL_CHUM | 0 to 150 | Female | IBD |
| 14428.32 | 12 | 40530.12 | 31 | CORIVA | 0 to 150 | Female | IBD |
| 251384.2 | 126 | 989156.2 | 607 | CPRDAurum | 0 to 150 | Female | IBD |
| 67601.03 | 22 | 161591.5 | 94 | CPRDGOLD | 0 to 150 | Male | IBD |
| 3680.871 | <5 | 7372.438 | 9 | IMASIS | 0 to 150 | Male | IBD |
| 0 | 0 | 9718.801 | 5 | AUSOM | 0 to 150 | Male | IBD |
| 53333.85 | 20 | 0 | 0 | IPCI | 0 to 150 | Male | IBD |
| 0 | 0 | 365.1828 | <5 | IPCI | 0 to 150 | Male | IBD |
| 0 | 0 | 21648.75 | 14 | eDOL_CHUM | 0 to 150 | Male | IBD |
| 12410.58 | 8 | 35345.67 | 28 | CORIVA | 0 to 150 | Male | IBD |
| 224303.6 | 139 | 833353.1 | 482 | CPRDAurum | 0 to 150 | Male | IBD |
| 5429.092 | <5 | 30446.29 | <5 | CPRDGOLD | 0 to 6 | Both | IBD |
| 85.78782 | <5 | 0 | 0 | IMASIS | 0 to 6 | Both | IBD |
| 0 | 0 | 392.1424 | <5 | IMASIS | 0 to 6 | Both | IBD |
| 0 | 0 | 494.5845 | <5 | AUSOM | 0 to 6 | Both | IBD |
| 2902.144 | <5 | 0 | 0 | IPCI | 0 to 6 | Both | IBD |
| 0 | 0 | 9.336071 | <5 | IPCI | 0 to 6 | Both | IBD |
| 0 | 0 | 2503.458 | <5 | eDOL_CHUM | 0 to 6 | Both | IBD |
| 1393.837 | <5 | 0 | 0 | CORIVA | 0 to 6 | Both | IBD |
| 0 | 0 | 1916.753 | <5 | CORIVA | 0 to 6 | Both | IBD |
| 14560.7 | <5 | 138816.5 | 6 | CPRDAurum | 0 to 6 | Both | IBD |
| 15590.3 | 6 | 26160 | 14 | CPRDGOLD | 12 to 18 | Both | IBD |
| 154.6995 | <5 | 0 | 0 | IMASIS | 12 to 18 | Both | IBD |
| 0 | 0 | 271.6112 | <5 | IMASIS | 12 to 18 | Both | IBD |
| 0 | 0 | 476.1177 | <5 | AUSOM | 12 to 18 | Both | IBD |
| 12812.53 | 6 | 0 | 0 | IPCI | 12 to 18 | Both | IBD |
| 0 | 0 | 51.27447 | <5 | IPCI | 12 to 18 | Both | IBD |
| 0 | 0 | 1940.999 | <5 | eDOL_CHUM | 12 to 18 | Both | IBD |
| 2642.141 | <5 | 5882.91 | 6 | CORIVA | 12 to 18 | Both | IBD |
| 56545.72 | 14 | 155714.5 | 82 | CPRDAurum | 12 to 18 | Both | IBD |
| 49728.34 | 26 | 113618.4 | 87 | CPRDGOLD | 19 to 40 | Both | IBD |
| 1327.08 | <5 | 3068.736 | 5 | IMASIS | 19 to 40 | Both | IBD |
| 0 | 0 | 3939.086 | <5 | AUSOM | 19 to 40 | Both | IBD |
| 35595.83 | 20 | 0 | 0 | IPCI | 19 to 40 | Both | IBD |
| 0 | 0 | 174.3491 | <5 | IPCI | 19 to 40 | Both | IBD |
| 0 | 0 | 17176.63 | 6 | eDOL_CHUM | 19 to 40 | Both | IBD |
| 9797.331 | 11 | 27104.03 | 26 | CORIVA | 19 to 40 | Both | IBD |
| 177499.1 | 118 | 611422.6 | 466 | CPRDAurum | 19 to 40 | Both | IBD |
| 49465.7 | 15 | 121957.1 | 70 | CPRDGOLD | 41 to 64 | Both | IBD |
| 3065.262 | <5 | 5483.151 | 5 | IMASIS | 41 to 64 | Both | IBD |
| 0 | 0 | 8554.965 | <5 | AUSOM | 41 to 64 | Both | IBD |
| 40610.57 | 24 | 0 | 0 | IPCI | 41 to 64 | Both | IBD |
| 0 | 0 | 320.9172 | <5 | IPCI | 41 to 64 | Both | IBD |
| 0 | 0 | 13146.23 | 10 | eDOL_CHUM | 41 to 64 | Both | IBD |
| 8520.052 | <5 | 25888.58 | 19 | CORIVA | 41 to 64 | Both | IBD |
| 154445.4 | 99 | 598533.1 | 411 | CPRDAurum | 41 to 64 | Both | IBD |
| 13576.56 | 5 | 44786.2 | 22 | CPRDGOLD | 65 to 150 | Both | IBD |
| 2934.705 | <5 | 6804.608 | 7 | IMASIS | 65 to 150 | Both | IBD |
| 0 | 0 | 6562.278 | <5 | AUSOM | 65 to 150 | Both | IBD |
| 12928.92 | <5 | 369.3634 | <5 | IPCI | 65 to 150 | Both | IBD |
| 0 | 0 | 12778.16 | 8 | eDOL_CHUM | 65 to 150 | Both | IBD |
| 2715.409 | <5 | 11117.11 | 8 | CORIVA | 65 to 150 | Both | IBD |
| 38773.11 | 22 | 172330.9 | 89 | CPRDAurum | 65 to 150 | Both | IBD |
| 10645.7 | <5 | 21379.76 | <5 | CPRDGOLD | 7 to 11 | Both | IBD |
| 75.99452 | <5 | 169.2676 | <5 | IMASIS | 7 to 11 | Both | IBD |
| 0 | 0 | 431.718 | <5 | AUSOM | 7 to 11 | Both | IBD |
| 6624.583 | <5 | 0 | 0 | IPCI | 7 to 11 | Both | IBD |
| 0 | 0 | 13.57974 | <5 | IPCI | 7 to 11 | Both | IBD |
| 0 | 0 | 899.0226 | <5 | eDOL_CHUM | 7 to 11 | Both | IBD |
| 1534.355 | <5 | 0 | 0 | CORIVA | 7 to 11 | Both | IBD |
| 0 | 0 | 2884.126 | <5 | CORIVA | 7 to 11 | Both | IBD |
| 25869.5 | 8 | 114961.1 | 17 | CPRDAurum | 7 to 11 | Both | IBD |
| 147266.1 | <5 | 365037.3 | <5 | CPRDGOLD | 0 to 150 | Both | Juvenile arthritis |
| 114154.9 | <5 | 0 | 0 | IPCI | 0 to 150 | Both | Juvenile arthritis |
| 26846.78 | <5 | 75891.65 | 9 | CORIVA | 0 to 150 | Both | Juvenile arthritis |
| 475774.2 | <5 | 1822868 | 24 | CPRDAurum | 0 to 150 | Both | Juvenile arthritis |
| 79656.76 | <5 | 203415 | <5 | CPRDGOLD | 0 to 150 | Female | Juvenile arthritis |
| 60813.6 | <5 | 0 | 0 | IPCI | 0 to 150 | Female | Juvenile arthritis |
| 14433.8 | <5 | 40538.23 | 5 | CORIVA | 0 to 150 | Female | Juvenile arthritis |
| 251424.1 | <5 | 989356.3 | 15 | CPRDAurum | 0 to 150 | Female | Juvenile arthritis |
| 67609.3 | <5 | 161622.2 | <5 | CPRDGOLD | 0 to 150 | Male | Juvenile arthritis |
| 53341.25 | <5 | 0 | 0 | IPCI | 0 to 150 | Male | Juvenile arthritis |
| 12412.98 | <5 | 35353.42 | <5 | CORIVA | 0 to 150 | Male | Juvenile arthritis |
| 224350.1 | <5 | 833511.9 | 9 | CPRDAurum | 0 to 150 | Male | Juvenile arthritis |
| 5429.092 | <5 | 30446.29 | <5 | CPRDGOLD | 0 to 6 | Both | Juvenile arthritis |
| 2902.144 | <5 | 0 | 0 | IPCI | 0 to 6 | Both | Juvenile arthritis |
| 1393.837 | <5 | 0 | 0 | CORIVA | 0 to 6 | Both | Juvenile arthritis |
| 0 | 0 | 1916.632 | <5 | CORIVA | 0 to 6 | Both | Juvenile arthritis |
| 14560.78 | 0 | 138814.9 | 6 | CPRDAurum | 0 to 6 | Both | Juvenile arthritis |
| 15591.4 | <5 | 26163.06 | <5 | CPRDGOLD | 12 to 18 | Both | Juvenile arthritis |
| 12814.13 | <5 | 0 | 0 | IPCI | 12 to 18 | Both | Juvenile arthritis |
| 2642.371 | <5 | 5882.209 | 6 | CORIVA | 12 to 18 | Both | Juvenile arthritis |
| 56549.69 | <5 | 155737.2 | 12 | CPRDAurum | 12 to 18 | Both | Juvenile arthritis |
| 49738.25 | <5 | 113646.5 | <5 | CPRDGOLD | 19 to 40 | Both | Juvenile arthritis |
| 35603.93 | <5 | 0 | 0 | IPCI | 19 to 40 | Both | Juvenile arthritis |
| 9801.785 | <5 | 27112.16 | <5 | CORIVA | 19 to 40 | Both | Juvenile arthritis |
| 177538 | 0 | 611584.8 | 0 | CPRDAurum | 19 to 40 | Both | Juvenile arthritis |
| 49470.84 | <5 | 121984.4 | <5 | CPRDGOLD | 41 to 64 | Both | Juvenile arthritis |
| 40619.09 | <5 | 0 | 0 | IPCI | 41 to 64 | Both | Juvenile arthritis |
| 8521.96 | <5 | 25894.75 | <5 | CORIVA | 41 to 64 | Both | Juvenile arthritis |
| 154478.9 | 0 | 598669.7 | 0 | CPRDAurum | 41 to 64 | Both | Juvenile arthritis |
| 13578.31 | <5 | 44793.74 | <5 | CPRDGOLD | 65 to 150 | Both | Juvenile arthritis |
| 12929.62 | <5 | 0 | 0 | IPCI | 65 to 150 | Both | Juvenile arthritis |
| 2716.942 | <5 | 11119.9 | <5 | CORIVA | 65 to 150 | Both | Juvenile arthritis |
| 38780.41 | 0 | 172361.9 | 0 | CPRDAurum | 65 to 150 | Both | Juvenile arthritis |
| 10645.91 | <5 | 21379.93 | <5 | CPRDGOLD | 7 to 11 | Both | Juvenile arthritis |
| 6624.583 | <5 | 0 | 0 | IPCI | 7 to 11 | Both | Juvenile arthritis |
| 1534.114 | <5 | 0 | 0 | CORIVA | 7 to 11 | Both | Juvenile arthritis |
| 0 | 0 | 2883.548 | <5 | CORIVA | 7 to 11 | Both | Juvenile arthritis |
| 25871.44 | 0 | 114964.9 | <5 | CPRDAurum | 7 to 11 | Both | Juvenile arthritis |
| 147238.1 | 76 | 365008.8 | 93 | CPRDGOLD | 0 to 150 | Both | ME/CFS diagnosis |
| 7746.212 | 7 | 16403.35 | 7 | IMASIS | 0 to 150 | Both | ME/CFS diagnosis |
| 0 | 0 | 20796.43 | 7 | AUSOM | 0 to 150 | Both | ME/CFS diagnosis |
| 114150.8 | 13 | 0 | 0 | IPCI | 0 to 150 | Both | ME/CFS diagnosis |
| 3941.881 | <5 | 49188.89 | 5 | eDOL_CHUM | 0 to 150 | Both | ME/CFS diagnosis |
| 26844.5 | 12 | 75888.22 | 20 | CORIVA | 0 to 150 | Both | ME/CFS diagnosis |
| 475721.7 | 144 | 1822677 | 618 | CPRDAurum | 0 to 150 | Both | ME/CFS diagnosis |
| 79638.05 | 51 | 203395.1 | 64 | CPRDGOLD | 0 to 150 | Female | ME/CFS diagnosis |
| 4065.331 | <5 | 9029.101 | 5 | IMASIS | 0 to 150 | Female | ME/CFS diagnosis |
| 0 | 0 | 11076.03 | 6 | AUSOM | 0 to 150 | Female | ME/CFS diagnosis |
| 60810.08 | 10 | 0 | 0 | IPCI | 0 to 150 | Female | ME/CFS diagnosis |
| 2229.35 | <5 | 27535.21 | <5 | eDOL_CHUM | 0 to 150 | Female | ME/CFS diagnosis |
| 14431.88 | 9 | 40534.73 | 14 | CORIVA | 0 to 150 | Female | ME/CFS diagnosis |
| 251387.5 | 103 | 989201.8 | 489 | CPRDAurum | 0 to 150 | Female | ME/CFS diagnosis |
| 67600.04 | 25 | 161613.7 | 29 | CPRDGOLD | 0 to 150 | Male | ME/CFS diagnosis |
| 3680.882 | <5 | 7374.253 | <5 | IMASIS | 0 to 150 | Male | ME/CFS diagnosis |
| 0 | 0 | 9720.402 | <5 | AUSOM | 0 to 150 | Male | ME/CFS diagnosis |
| 53340.74 | <5 | 0 | 0 | IPCI | 0 to 150 | Male | ME/CFS diagnosis |
| 1712.531 | <5 | 21653.68 | <5 | eDOL_CHUM | 0 to 150 | Male | ME/CFS diagnosis |
| 12412.62 | <5 | 35353.49 | 6 | CORIVA | 0 to 150 | Male | ME/CFS diagnosis |
| 224334.1 | 41 | 833475.4 | 129 | CPRDAurum | 0 to 150 | Male | ME/CFS diagnosis |
| 5429.092 | <5 | 30446.29 | <5 | CPRDGOLD | 0 to 6 | Both | ME/CFS diagnosis |
| 85.78782 | <5 | 0 | 0 | IMASIS | 0 to 6 | Both | ME/CFS diagnosis |
| 0 | 0 | 392.1424 | <5 | IMASIS | 0 to 6 | Both | ME/CFS diagnosis |
| 0 | 0 | 494.5845 | <5 | AUSOM | 0 to 6 | Both | ME/CFS diagnosis |
| 2902.144 | <5 | 0 | 0 | IPCI | 0 to 6 | Both | ME/CFS diagnosis |
| 173.8617 | <5 | 0 | 0 | eDOL_CHUM | 0 to 6 | Both | ME/CFS diagnosis |
| 0 | 0 | 2503.458 | <5 | eDOL_CHUM | 0 to 6 | Both | ME/CFS diagnosis |
| 1393.837 | <5 | 0 | 0 | CORIVA | 0 to 6 | Both | ME/CFS diagnosis |
| 0 | 0 | 1916.717 | <5 | CORIVA | 0 to 6 | Both | ME/CFS diagnosis |
| 14560.78 | 0 | 138816.6 | 7 | CPRDAurum | 0 to 6 | Both | ME/CFS diagnosis |
| 15589.79 | 6 | 26161.19 | 8 | CPRDGOLD | 12 to 18 | Both | ME/CFS diagnosis |
| 154.6995 | <5 | 0 | 0 | IMASIS | 12 to 18 | Both | ME/CFS diagnosis |
| 0 | 0 | 271.6112 | <5 | IMASIS | 12 to 18 | Both | ME/CFS diagnosis |
| 0 | 0 | 476.1177 | <5 | AUSOM | 12 to 18 | Both | ME/CFS diagnosis |
| 12814.76 | <5 | 0 | 0 | IPCI | 12 to 18 | Both | ME/CFS diagnosis |
| 104.7803 | <5 | 1940.999 | <5 | eDOL_CHUM | 12 to 18 | Both | ME/CFS diagnosis |
| 2641.873 | <5 | 5883.874 | <5 | CORIVA | 12 to 18 | Both | ME/CFS diagnosis |
| 56545.94 | 18 | 155724.7 | 60 | CPRDAurum | 12 to 18 | Both | ME/CFS diagnosis |
| 49730.51 | 23 | 113634.8 | 34 | CPRDGOLD | 19 to 40 | Both | ME/CFS diagnosis |
| 1326.943 | <5 | 3069.922 | <5 | IMASIS | 19 to 40 | Both | ME/CFS diagnosis |
| 0 | 0 | 3939.696 | <5 | AUSOM | 19 to 40 | Both | ME/CFS diagnosis |
| 35602.63 | 5 | 0 | 0 | IPCI | 19 to 40 | Both | ME/CFS diagnosis |
| 1168.95 | <5 | 17178.92 | <5 | eDOL_CHUM | 19 to 40 | Both | ME/CFS diagnosis |
| 9800.334 | 5 | 27110.77 | 5 | CORIVA | 19 to 40 | Both | ME/CFS diagnosis |
| 177513.7 | 63 | 611503.4 | 251 | CPRDAurum | 19 to 40 | Both | ME/CFS diagnosis |
| 49454.32 | 43 | 121970.6 | 40 | CPRDGOLD | 41 to 64 | Both | ME/CFS diagnosis |
| 3064.37 | 6 | 5483.975 | 5 | IMASIS | 41 to 64 | Both | ME/CFS diagnosis |
| 0 | 0 | 8555.006 | 6 | AUSOM | 41 to 64 | Both | ME/CFS diagnosis |
| 40615.76 | 7 | 0 | 0 | IPCI | 41 to 64 | Both | ME/CFS diagnosis |
| 1140.485 | <5 | 13149.74 | <5 | eDOL_CHUM | 41 to 64 | Both | ME/CFS diagnosis |
| 8521.711 | <5 | 25892.44 | 7 | CORIVA | 41 to 64 | Both | ME/CFS diagnosis |
| 154456.2 | 55 | 598585.6 | 254 | CPRDAurum | 41 to 64 | Both | ME/CFS diagnosis |
| 13577.21 | <5 | 44792.42 | 8 | CPRDGOLD | 65 to 150 | Both | ME/CFS diagnosis |
| 2935.789 | <5 | 6806.374 | <5 | IMASIS | 65 to 150 | Both | ME/CFS diagnosis |
| 0 | 0 | 6562.374 | <5 | AUSOM | 65 to 150 | Both | ME/CFS diagnosis |
| 12929.59 | <5 | 0 | 0 | IPCI | 65 to 150 | Both | ME/CFS diagnosis |
| 1242.888 | <5 | 12781.61 | <5 | eDOL_CHUM | 65 to 150 | Both | ME/CFS diagnosis |
| 2716.654 | <5 | 11118.43 | 5 | CORIVA | 65 to 150 | Both | ME/CFS diagnosis |
| 38778.77 | 6 | 172351.3 | 31 | CPRDAurum | 65 to 150 | Both | ME/CFS diagnosis |
| 10645.91 | <5 | 21380.4 | <5 | CPRDGOLD | 7 to 11 | Both | ME/CFS diagnosis |
| 75.99452 | <5 | 169.2676 | <5 | IMASIS | 7 to 11 | Both | ME/CFS diagnosis |
| 0 | 0 | 431.718 | <5 | AUSOM | 7 to 11 | Both | ME/CFS diagnosis |
| 6624.583 | <5 | 0 | 0 | IPCI | 7 to 11 | Both | ME/CFS diagnosis |
| 63.60849 | <5 | 0 | 0 | eDOL_CHUM | 7 to 11 | Both | ME/CFS diagnosis |
| 0 | 0 | 899.0226 | <5 | eDOL_CHUM | 7 to 11 | Both | ME/CFS diagnosis |
| 1534.322 | <5 | 0 | 0 | CORIVA | 7 to 11 | Both | ME/CFS diagnosis |
| 0 | 0 | 2883.406 | <5 | CORIVA | 7 to 11 | Both | ME/CFS diagnosis |
| 25871.24 | <5 | 114963 | 9 | CPRDAurum | 7 to 11 | Both | ME/CFS diagnosis |
| 146934.6 | 1182 | 364087.9 | 3040 | CPRDGOLD | 0 to 150 | Both | ME/CFS symptoms |
| 7729.164 | 94 | 16346.43 | 261 | IMASIS | 0 to 150 | Both | ME/CFS symptoms |
| 320.8323 | <5 | 0 | 0 | AUSOM | 0 to 150 | Both | ME/CFS symptoms |
| 0 | 0 | 20772.75 | 68 | AUSOM | 0 to 150 | Both | ME/CFS symptoms |
| 113154.6 | 2933 | 945.0678 | 30 | IPCI | 0 to 150 | Both | ME/CFS symptoms |
| 3941.372 | <5 | 49170.91 | 60 | eDOL_CHUM | 0 to 150 | Both | ME/CFS symptoms |
| 26729.02 | 353 | 75586.6 | 942 | CORIVA | 0 to 150 | Both | ME/CFS symptoms |
| 473678.8 | 6761 | 1815318 | 23131 | CPRDAurum | 0 to 150 | Both | ME/CFS symptoms |
| 79435.57 | 790 | 202752.7 | 2127 | CPRDGOLD | 0 to 150 | Female | ME/CFS symptoms |
| 4054.965 | 45 | 8998.719 | 135 | IMASIS | 0 to 150 | Female | ME/CFS symptoms |
| 180.9993 | <5 | 0 | 0 | AUSOM | 0 to 150 | Female | ME/CFS symptoms |
| 0 | 0 | 11061.26 | 45 | AUSOM | 0 to 150 | Female | ME/CFS symptoms |
| 60167.83 | 1899 | 583.0198 | 20 | IPCI | 0 to 150 | Female | ME/CFS symptoms |
| 2229.35 | <5 | 27530.62 | 22 | eDOL_CHUM | 0 to 150 | Female | ME/CFS symptoms |
| 14354.09 | 241 | 40337.76 | 616 | CORIVA | 0 to 150 | Female | ME/CFS symptoms |
| 249991 | 4551 | 984110.4 | 15888 | CPRDAurum | 0 to 150 | Female | ME/CFS symptoms |
| 67499.02 | 392 | 161335.2 | 913 | CPRDGOLD | 0 to 150 | Male | ME/CFS symptoms |
| 3674.198 | 49 | 7347.713 | 126 | IMASIS | 0 to 150 | Male | ME/CFS symptoms |
| 139.833 | <5 | 0 | 0 | AUSOM | 0 to 150 | Male | ME/CFS symptoms |
| 0 | 0 | 9711.491 | 23 | AUSOM | 0 to 150 | Male | ME/CFS symptoms |
| 52986.73 | 1034 | 0 | 0 | IPCI | 0 to 150 | Male | ME/CFS symptoms |
| 0 | 0 | 362.0479 | 10 | IPCI | 0 to 150 | Male | ME/CFS symptoms |
| 1712.022 | <5 | 21640.29 | 38 | eDOL_CHUM | 0 to 150 | Male | ME/CFS symptoms |
| 12374.93 | 112 | 35248.84 | 326 | CORIVA | 0 to 150 | Male | ME/CFS symptoms |
| 223687.8 | 2210 | 831207.7 | 7243 | CPRDAurum | 0 to 150 | Male | ME/CFS symptoms |
| 5420.011 | 31 | 30395.85 | 172 | CPRDGOLD | 0 to 6 | Both | ME/CFS symptoms |
| 85.78782 | <5 | 0 | 0 | IMASIS | 0 to 6 | Both | ME/CFS symptoms |
| 0 | 0 | 392.1424 | <5 | IMASIS | 0 to 6 | Both | ME/CFS symptoms |
| 9.98768 | <5 | 0 | 0 | AUSOM | 0 to 6 | Both | ME/CFS symptoms |
| 0 | 0 | 494.5845 | <5 | AUSOM | 0 to 6 | Both | ME/CFS symptoms |
| 2888.068 | 43 | 0 | 0 | IPCI | 0 to 6 | Both | ME/CFS symptoms |
| 0 | 0 | 9.336071 | <5 | IPCI | 0 to 6 | Both | ME/CFS symptoms |
| 173.8617 | <5 | 0 | 0 | eDOL_CHUM | 0 to 6 | Both | ME/CFS symptoms |
| 0 | 0 | 2503.335 | <5 | eDOL_CHUM | 0 to 6 | Both | ME/CFS symptoms |
| 1393.155 | 6 | 0 | 0 | CORIVA | 0 to 6 | Both | ME/CFS symptoms |
| 0 | 0 | 1912.444 | 13 | CORIVA | 0 to 6 | Both | ME/CFS symptoms |
| 14534.71 | 101 | 138555.5 | 893 | CPRDAurum | 0 to 6 | Both | ME/CFS symptoms |
| 15567 | 107 | 26102.67 | 202 | CPRDGOLD | 12 to 18 | Both | ME/CFS symptoms |
| 154.6995 | <5 | 0 | 0 | IMASIS | 12 to 18 | Both | ME/CFS symptoms |
| 0 | 0 | 271.6112 | <5 | IMASIS | 12 to 18 | Both | ME/CFS symptoms |
| 10.02601 | <5 | 0 | 0 | AUSOM | 12 to 18 | Both | ME/CFS symptoms |
| 0 | 0 | 475.0062 | <5 | AUSOM | 12 to 18 | Both | ME/CFS symptoms |
| 12684.16 | 417 | 0 | 0 | IPCI | 12 to 18 | Both | ME/CFS symptoms |
| 0 | 0 | 50.65572 | <5 | IPCI | 12 to 18 | Both | ME/CFS symptoms |
| 104.7803 | <5 | 1940.597 | <5 | eDOL_CHUM | 12 to 18 | Both | ME/CFS symptoms |
| 2634.475 | 30 | 5863.71 | 67 | CORIVA | 12 to 18 | Both | ME/CFS symptoms |
| 56368.42 | 778 | 155229.1 | 1806 | CPRDAurum | 12 to 18 | Both | ME/CFS symptoms |
| 49606.13 | 470 | 113274.3 | 1143 | CPRDGOLD | 19 to 40 | Both | ME/CFS symptoms |
| 1325.027 | 7 | 3062.669 | 33 | IMASIS | 19 to 40 | Both | ME/CFS symptoms |
| 89.20739 | <5 | 0 | 0 | AUSOM | 19 to 40 | Both | ME/CFS symptoms |
| 0 | 0 | 3932.123 | 25 | AUSOM | 19 to 40 | Both | ME/CFS symptoms |
| 35239.43 | 1068 | 0 | 0 | IPCI | 19 to 40 | Both | ME/CFS symptoms |
| 0 | 0 | 173.8316 | <5 | IPCI | 19 to 40 | Both | ME/CFS symptoms |
| 1168.95 | <5 | 17176.49 | 8 | eDOL_CHUM | 19 to 40 | Both | ME/CFS symptoms |
| 9745.785 | 156 | 26983.8 | 395 | CORIVA | 19 to 40 | Both | ME/CFS symptoms |
| 176561.4 | 3068 | 608212.7 | 10110 | CPRDAurum | 19 to 40 | Both | ME/CFS symptoms |
| 49349.93 | 406 | 121680.2 | 963 | CPRDGOLD | 41 to 64 | Both | ME/CFS symptoms |
| 3058.398 | 33 | 5467.105 | 70 | IMASIS | 41 to 64 | Both | ME/CFS symptoms |
| 109.1828 | <5 | 0 | 0 | AUSOM | 41 to 64 | Both | ME/CFS symptoms |
| 0 | 0 | 8545.78 | 26 | AUSOM | 41 to 64 | Both | ME/CFS symptoms |
| 40274.85 | 962 | 0 | 0 | IPCI | 41 to 64 | Both | ME/CFS symptoms |
| 0 | 0 | 318.7981 | 7 | IPCI | 41 to 64 | Both | ME/CFS symptoms |
| 1140.485 | <5 | 13145.41 | 17 | eDOL_CHUM | 41 to 64 | Both | ME/CFS symptoms |
| 8485.684 | 115 | 25794.21 | 302 | CORIVA | 41 to 64 | Both | ME/CFS symptoms |
| 153801.9 | 2027 | 596217.5 | 7265 | CPRDAurum | 41 to 64 | Both | ME/CFS symptoms |
| 13546.42 | 113 | 44663.96 | 445 | CPRDGOLD | 65 to 150 | Both | ME/CFS symptoms |
| 2926.628 | 54 | 6773.717 | 155 | IMASIS | 65 to 150 | Both | ME/CFS symptoms |
| 77.34702 | <5 | 0 | 0 | AUSOM | 65 to 150 | Both | ME/CFS symptoms |
| 0 | 0 | 6556.772 | 14 | AUSOM | 65 to 150 | Both | ME/CFS symptoms |
| 12834.53 | 305 | 363.4962 | 17 | IPCI | 65 to 150 | Both | ME/CFS symptoms |
| 1242.379 | <5 | 12771.54 | 32 | eDOL_CHUM | 65 to 150 | Both | ME/CFS symptoms |
| 2701.27 | 42 | 11075.81 | 137 | CORIVA | 65 to 150 | Both | ME/CFS symptoms |
| 38599.23 | 566 | 171625.2 | 2332 | CPRDAurum | 65 to 150 | Both | ME/CFS symptoms |
| 10638.31 | 39 | 21363.57 | 64 | CPRDGOLD | 7 to 11 | Both | ME/CFS symptoms |
| 75.99452 | <5 | 169.2676 | <5 | IMASIS | 7 to 11 | Both | ME/CFS symptoms |
| 19.77823 | <5 | 0 | 0 | AUSOM | 7 to 11 | Both | ME/CFS symptoms |
| 0 | 0 | 431.718 | <5 | AUSOM | 7 to 11 | Both | ME/CFS symptoms |
| 6600.575 | 77 | 0 | 0 | IPCI | 7 to 11 | Both | ME/CFS symptoms |
| 0 | 0 | 13.51403 | <5 | IPCI | 7 to 11 | Both | ME/CFS symptoms |
| 63.60849 | <5 | 0 | 0 | eDOL_CHUM | 7 to 11 | Both | ME/CFS symptoms |
| 0 | 0 | 899.0226 | <5 | eDOL_CHUM | 7 to 11 | Both | ME/CFS symptoms |
| 1533.5 | <5 | 0 | 0 | CORIVA | 7 to 11 | Both | ME/CFS symptoms |
| 0 | 0 | 2879.381 | 16 | CORIVA | 7 to 11 | Both | ME/CFS symptoms |
| 25846.31 | 125 | 114839.8 | 432 | CPRDAurum | 7 to 11 | Both | ME/CFS symptoms |
| 0 | 0 | 49190.43 | <5 | eDOL_CHUM | 0 to 150 | Both | MIS |
| 26846.72 | <5 | 75893.94 | <5 | CORIVA | 0 to 150 | Both | MIS |
| 475775.6 | <5 | 1822877 | <5 | CPRDAurum | 0 to 150 | Both | MIS |
| 0 | 0 | 27535.72 | <5 | eDOL_CHUM | 0 to 150 | Female | MIS |
| 14433.8 | <5 | 40539.49 | <5 | CORIVA | 0 to 150 | Female | MIS |
| 251425.3 | <5 | 989361.2 | <5 | CPRDAurum | 0 to 150 | Female | MIS |
| 0 | 0 | 21654.71 | <5 | eDOL_CHUM | 0 to 150 | Male | MIS |
| 12412.92 | <5 | 35354.45 | <5 | CORIVA | 0 to 150 | Male | MIS |
| 224350.3 | <5 | 833515.6 | <5 | CPRDAurum | 0 to 150 | Male | MIS |
| 0 | 0 | 2503.458 | <5 | eDOL_CHUM | 0 to 6 | Both | MIS |
| 1393.837 | <5 | 0 | 0 | CORIVA | 0 to 6 | Both | MIS |
| 0 | 0 | 1916.753 | <5 | CORIVA | 0 to 6 | Both | MIS |
| 14560.66 | <5 | 138818.3 | 0 | CPRDAurum | 0 to 6 | Both | MIS |
| 0 | 0 | 1940.999 | <5 | eDOL_CHUM | 12 to 18 | Both | MIS |
| 2642.79 | <5 | 5883.89 | <5 | CORIVA | 12 to 18 | Both | MIS |
| 56551.12 | <5 | 155740.5 | <5 | CPRDAurum | 12 to 18 | Both | MIS |
| 0 | 0 | 17179.95 | <5 | eDOL_CHUM | 19 to 40 | Both | MIS |
| 9801.785 | <5 | 27112.13 | <5 | CORIVA | 19 to 40 | Both | MIS |
| 177538 | 0 | 611584.8 | 0 | CPRDAurum | 19 to 40 | Both | MIS |
| 0 | 0 | 13150.26 | <5 | eDOL_CHUM | 41 to 64 | Both | MIS |
| 8521.96 | <5 | 25894.75 | <5 | CORIVA | 41 to 64 | Both | MIS |
| 154478.9 | 0 | 598669.7 | 0 | CPRDAurum | 41 to 64 | Both | MIS |
| 0 | 0 | 12781.61 | <5 | eDOL_CHUM | 65 to 150 | Both | MIS |
| 2716.942 | <5 | 11119.72 | <5 | CORIVA | 65 to 150 | Both | MIS |
| 38780.41 | 0 | 172361.9 | 0 | CPRDAurum | 65 to 150 | Both | MIS |
| 0 | 0 | 899.0226 | <5 | eDOL_CHUM | 7 to 11 | Both | MIS |
| 1533.634 | <5 | 0 | 0 | CORIVA | 7 to 11 | Both | MIS |
| 0 | 0 | 2884.126 | <5 | CORIVA | 7 to 11 | Both | MIS |
| 25871.43 | <5 | 114965.7 | <5 | CPRDAurum | 7 to 11 | Both | MIS |
| 147234.4 | 112 | 364942.7 | 300 | CPRDGOLD | 0 to 150 | Both | POTS diagnosis |
| 7743.102 | 23 | 16392.31 | 46 | IMASIS | 0 to 150 | Both | POTS diagnosis |
| 320.95 | <5 | 0 | 0 | AUSOM | 0 to 150 | Both | POTS diagnosis |
| 0 | 0 | 20794.55 | 13 | AUSOM | 0 to 150 | Both | POTS diagnosis |
| 114126.4 | 81 | 0 | 0 | IPCI | 0 to 150 | Both | POTS diagnosis |
| 3938.719 | 8 | 49168.4 | 58 | eDOL_CHUM | 0 to 150 | Both | POTS diagnosis |
| 26816.32 | 93 | 75815 | 238 | CORIVA | 0 to 150 | Both | POTS diagnosis |
| 475549.7 | 729 | 1821978 | 2735 | CPRDAurum | 0 to 150 | Both | POTS diagnosis |
| 79636.77 | 69 | 203351.4 | 208 | CPRDGOLD | 0 to 150 | Female | POTS diagnosis |
| 4063.299 | 14 | 9022.5 | 24 | IMASIS | 0 to 150 | Female | POTS diagnosis |
| 181.117 | <5 | 0 | 0 | AUSOM | 0 to 150 | Female | POTS diagnosis |
| 0 | 0 | 11075.87 | <5 | AUSOM | 0 to 150 | Female | POTS diagnosis |
| 60796.15 | 53 | 0 | 0 | IPCI | 0 to 150 | Female | POTS diagnosis |
| 2228.6 | <5 | 27527.04 | 26 | eDOL_CHUM | 0 to 150 | Female | POTS diagnosis |
| 14413.36 | 59 | 40488.48 | 161 | CORIVA | 0 to 150 | Female | POTS diagnosis |
| 251277 | 471 | 988760.8 | 1834 | CPRDAurum | 0 to 150 | Female | POTS diagnosis |
| 67597.64 | 43 | 161591.3 | 92 | CPRDGOLD | 0 to 150 | Male | POTS diagnosis |
| 3679.803 | 9 | 7369.81 | 22 | IMASIS | 0 to 150 | Male | POTS diagnosis |
| 139.833 | <5 | 0 | 0 | AUSOM | 0 to 150 | Male | POTS diagnosis |
| 0 | 0 | 9718.683 | 9 | AUSOM | 0 to 150 | Male | POTS diagnosis |
| 53330.26 | 28 | 0 | 0 | IPCI | 0 to 150 | Male | POTS diagnosis |
| 1710.119 | 6 | 21641.37 | 32 | eDOL_CHUM | 0 to 150 | Male | POTS diagnosis |
| 12402.96 | 34 | 35326.52 | 77 | CORIVA | 0 to 150 | Male | POTS diagnosis |
| 224272.7 | 258 | 833217.3 | 901 | CPRDAurum | 0 to 150 | Male | POTS diagnosis |
| 5429.092 | <5 | 30444.45 | 6 | CPRDGOLD | 0 to 6 | Both | POTS diagnosis |
| 85.78782 | <5 | 0 | 0 | IMASIS | 0 to 6 | Both | POTS diagnosis |
| 0 | 0 | 392.1424 | <5 | IMASIS | 0 to 6 | Both | POTS diagnosis |
| 9.98768 | <5 | 0 | 0 | AUSOM | 0 to 6 | Both | POTS diagnosis |
| 0 | 0 | 494.5845 | <5 | AUSOM | 0 to 6 | Both | POTS diagnosis |
| 2902.144 | <5 | 0 | 0 | IPCI | 0 to 6 | Both | POTS diagnosis |
| 173.8617 | <5 | 0 | 0 | eDOL_CHUM | 0 to 6 | Both | POTS diagnosis |
| 0 | 0 | 2503.176 | <5 | eDOL_CHUM | 0 to 6 | Both | POTS diagnosis |
| 1393.837 | <5 | 0 | 0 | CORIVA | 0 to 6 | Both | POTS diagnosis |
| 0 | 0 | 1916.753 | <5 | CORIVA | 0 to 6 | Both | POTS diagnosis |
| 14559.64 | <5 | 138806.3 | 41 | CPRDAurum | 0 to 6 | Both | POTS diagnosis |
| 15589.54 | 8 | 26159.55 | 12 | CPRDGOLD | 12 to 18 | Both | POTS diagnosis |
| 154.6995 | <5 | 0 | 0 | IMASIS | 12 to 18 | Both | POTS diagnosis |
| 0 | 0 | 271.6112 | <5 | IMASIS | 12 to 18 | Both | POTS diagnosis |
| 10.02601 | <5 | 0 | 0 | AUSOM | 12 to 18 | Both | POTS diagnosis |
| 0 | 0 | 476.1177 | <5 | AUSOM | 12 to 18 | Both | POTS diagnosis |
| 12813.99 | <5 | 0 | 0 | IPCI | 12 to 18 | Both | POTS diagnosis |
| 104.7803 | <5 | 1939.556 | <5 | eDOL_CHUM | 12 to 18 | Both | POTS diagnosis |
| 2640.871 | 6 | 5879.113 | 17 | CORIVA | 12 to 18 | Both | POTS diagnosis |
| 56535.79 | 55 | 155693.3 | 153 | CPRDAurum | 12 to 18 | Both | POTS diagnosis |
| 49728.41 | 37 | 113617.9 | 99 | CPRDGOLD | 19 to 40 | Both | POTS diagnosis |
| 1326.702 | <5 | 3067.477 | 9 | IMASIS | 19 to 40 | Both | POTS diagnosis |
| 89.20739 | <5 | 0 | 0 | AUSOM | 19 to 40 | Both | POTS diagnosis |
| 0 | 0 | 3939.696 | <5 | AUSOM | 19 to 40 | Both | POTS diagnosis |
| 35597.04 | 20 | 0 | 0 | IPCI | 19 to 40 | Both | POTS diagnosis |
| 1168.95 | <5 | 17177.14 | 7 | eDOL_CHUM | 19 to 40 | Both | POTS diagnosis |
| 9788.742 | 37 | 27084.53 | 83 | CORIVA | 19 to 40 | Both | POTS diagnosis |
| 177454.2 | 262 | 611272.7 | 951 | CPRDAurum | 19 to 40 | Both | POTS diagnosis |
| 49458.37 | 43 | 121947.1 | 111 | CPRDGOLD | 41 to 64 | Both | POTS diagnosis |
| 3063.83 | 9 | 5481.344 | 12 | IMASIS | 41 to 64 | Both | POTS diagnosis |
| 109.1828 | <5 | 0 | 0 | AUSOM | 41 to 64 | Both | POTS diagnosis |
| 0 | 0 | 8553.908 | 7 | AUSOM | 41 to 64 | Both | POTS diagnosis |
| 40605.63 | 35 | 0 | 0 | IPCI | 41 to 64 | Both | POTS diagnosis |
| 1140.485 | <5 | 13143.9 | 17 | eDOL_CHUM | 41 to 64 | Both | POTS diagnosis |
| 8510.894 | 35 | 25865.43 | 92 | CORIVA | 41 to 64 | Both | POTS diagnosis |
| 154393.4 | 282 | 598327.7 | 989 | CPRDAurum | 41 to 64 | Both | POTS diagnosis |
| 13571.93 | 20 | 44772.15 | 66 | CPRDGOLD | 65 to 150 | Both | POTS diagnosis |
| 2933.394 | 12 | 6800.405 | 25 | IMASIS | 65 to 150 | Both | POTS diagnosis |
| 77.46475 | <5 | 0 | 0 | AUSOM | 65 to 150 | Both | POTS diagnosis |
| 0 | 0 | 6561.588 | 6 | AUSOM | 65 to 150 | Both | POTS diagnosis |
| 12923.52 | 18 | 0 | 0 | IPCI | 65 to 150 | Both | POTS diagnosis |
| 1240.298 | 7 | 12770.55 | 30 | eDOL_CHUM | 65 to 150 | Both | POTS diagnosis |
| 2712.468 | 15 | 11103.9 | 43 | CORIVA | 65 to 150 | Both | POTS diagnosis |
| 38745.54 | 110 | 172194 | 535 | CPRDAurum | 65 to 150 | Both | POTS diagnosis |
| 10645.62 | <5 | 21379.51 | <5 | CPRDGOLD | 7 to 11 | Both | POTS diagnosis |
| 75.99452 | <5 | 169.2676 | <5 | IMASIS | 7 to 11 | Both | POTS diagnosis |
| 19.77823 | <5 | 0 | 0 | AUSOM | 7 to 11 | Both | POTS diagnosis |
| 0 | 0 | 431.718 | <5 | AUSOM | 7 to 11 | Both | POTS diagnosis |
| 6623.641 | <5 | 0 | 0 | IPCI | 7 to 11 | Both | POTS diagnosis |
| 63.60849 | <5 | 0 | 0 | eDOL_CHUM | 7 to 11 | Both | POTS diagnosis |
| 0 | 0 | 899.0226 | <5 | eDOL_CHUM | 7 to 11 | Both | POTS diagnosis |
| 1534.355 | <5 | 0 | 0 | CORIVA | 7 to 11 | Both | POTS diagnosis |
| 0 | 0 | 2884.09 | <5 | CORIVA | 7 to 11 | Both | POTS diagnosis |
| 25869.51 | 6 | 114960.9 | 26 | CPRDAurum | 7 to 11 | Both | POTS diagnosis |
| 146832.3 | 1552 | 363762.8 | 4043 | CPRDGOLD | 0 to 150 | Both | POTS symptoms |
| 7723.918 | 95 | 16357.43 | 181 | IMASIS | 0 to 150 | Both | POTS symptoms |
| 319.1923 | 6 | 0 | 0 | AUSOM | 0 to 150 | Both | POTS symptoms |
| 0 | 0 | 20731.23 | 189 | AUSOM | 0 to 150 | Both | POTS symptoms |
| 112692.6 | 4309 | 943.4661 | 34 | IPCI | 0 to 150 | Both | POTS symptoms |
| 3936.698 | 13 | 49138.57 | 145 | eDOL_CHUM | 0 to 150 | Both | POTS symptoms |
| 26683.98 | 467 | 75432.08 | 1402 | CORIVA | 0 to 150 | Both | POTS symptoms |
| 473086.4 | 8350 | 1812730 | 30085 | CPRDAurum | 0 to 150 | Both | POTS symptoms |
| 79385.42 | 995 | 202568.3 | 2653 | CPRDGOLD | 0 to 150 | Female | POTS symptoms |
| 4050.281 | 58 | 9002.672 | 99 | IMASIS | 0 to 150 | Female | POTS symptoms |
| 180.4682 | <5 | 0 | 0 | AUSOM | 0 to 150 | Female | POTS symptoms |
| 0 | 0 | 11040.23 | 108 | AUSOM | 0 to 150 | Female | POTS symptoms |
| 59899.99 | 2699 | 582.5243 | 23 | IPCI | 0 to 150 | Female | POTS symptoms |
| 2227.762 | <5 | 27512.43 | 69 | eDOL_CHUM | 0 to 150 | Female | POTS symptoms |
| 14324.68 | 315 | 40229.69 | 938 | CORIVA | 0 to 150 | Female | POTS symptoms |
| 249678.5 | 5343 | 982673.3 | 19568 | CPRDAurum | 0 to 150 | Female | POTS symptoms |
| 67446.88 | 557 | 161194.5 | 1390 | CPRDGOLD | 0 to 150 | Male | POTS symptoms |
| 3673.637 | 37 | 7354.757 | 82 | IMASIS | 0 to 150 | Male | POTS symptoms |
| 138.7242 | <5 | 0 | 0 | AUSOM | 0 to 150 | Male | POTS symptoms |
| 0 | 0 | 9691.001 | 81 | AUSOM | 0 to 150 | Male | POTS symptoms |
| 52792.66 | 1610 | 0 | 0 | IPCI | 0 to 150 | Male | POTS symptoms |
| 0 | 0 | 360.9418 | 11 | IPCI | 0 to 150 | Male | POTS symptoms |
| 1708.936 | 9 | 21626.13 | 76 | eDOL_CHUM | 0 to 150 | Male | POTS symptoms |
| 12359.3 | 152 | 35202.38 | 464 | CORIVA | 0 to 150 | Male | POTS symptoms |
| 223407.9 | 3007 | 830057 | 10517 | CPRDAurum | 0 to 150 | Male | POTS symptoms |
| 5425.87 | 14 | 30429.34 | 64 | CPRDGOLD | 0 to 6 | Both | POTS symptoms |
| 85.68652 | <5 | 0 | 0 | IMASIS | 0 to 6 | Both | POTS symptoms |
| 0 | 0 | 392.0411 | <5 | IMASIS | 0 to 6 | Both | POTS symptoms |
| 9.98768 | <5 | 0 | 0 | AUSOM | 0 to 6 | Both | POTS symptoms |
| 0 | 0 | 494.371 | <5 | AUSOM | 0 to 6 | Both | POTS symptoms |
| 2884.862 | 53 | 0 | 0 | IPCI | 0 to 6 | Both | POTS symptoms |
| 0 | 0 | 9.336071 | <5 | IPCI | 0 to 6 | Both | POTS symptoms |
| 173.8617 | <5 | 0 | 0 | eDOL_CHUM | 0 to 6 | Both | POTS symptoms |
| 0 | 0 | 2503.176 | <5 | eDOL_CHUM | 0 to 6 | Both | POTS symptoms |
| 1392.966 | <5 | 0 | 0 | CORIVA | 0 to 6 | Both | POTS symptoms |
| 0 | 0 | 1912.802 | 9 | CORIVA | 0 to 6 | Both | POTS symptoms |
| 14546.19 | 57 | 138692.4 | 398 | CPRDAurum | 0 to 6 | Both | POTS symptoms |
| 15550.49 | 178 | 26074.07 | 291 | CPRDGOLD | 12 to 18 | Both | POTS symptoms |
| 154.2149 | <5 | 0 | 0 | IMASIS | 12 to 18 | Both | POTS symptoms |
| 0 | 0 | 270.883 | <5 | IMASIS | 12 to 18 | Both | POTS symptoms |
| 10.02601 | <5 | 0 | 0 | AUSOM | 12 to 18 | Both | POTS symptoms |
| 0 | 0 | 474.1684 | 6 | AUSOM | 12 to 18 | Both | POTS symptoms |
| 12616.83 | 599 | 0 | 0 | IPCI | 12 to 18 | Both | POTS symptoms |
| 0 | 0 | 50.65572 | <5 | IPCI | 12 to 18 | Both | POTS symptoms |
| 104.7803 | <5 | 1939.556 | <5 | eDOL_CHUM | 12 to 18 | Both | POTS symptoms |
| 2632.096 | 37 | 5856.181 | 96 | CORIVA | 12 to 18 | Both | POTS symptoms |
| 56286.94 | 1053 | 154934.2 | 2622 | CPRDAurum | 12 to 18 | Both | POTS symptoms |
| 49591.59 | 533 | 113201.2 | 1402 | CPRDGOLD | 19 to 40 | Both | POTS symptoms |
| 1324.621 | 9 | 3062.352 | 30 | IMASIS | 19 to 40 | Both | POTS symptoms |
| 89.10062 | <5 | 0 | 0 | AUSOM | 19 to 40 | Both | POTS symptoms |
| 0 | 0 | 3925.593 | 34 | AUSOM | 19 to 40 | Both | POTS symptoms |
| 35113.92 | 1421 | 0 | 0 | IPCI | 19 to 40 | Both | POTS symptoms |
| 0 | 0 | 172.6242 | 6 | IPCI | 19 to 40 | Both | POTS symptoms |
| 1168.63 | <5 | 17173.57 | 16 | eDOL_CHUM | 19 to 40 | Both | POTS symptoms |
| 9732.808 | 196 | 26939.19 | 524 | CORIVA | 19 to 40 | Both | POTS symptoms |
| 176400 | 3374 | 607556 | 11702 | CPRDAurum | 19 to 40 | Both | POTS symptoms |
| 49309.45 | 544 | 121523.4 | 1450 | CPRDGOLD | 41 to 64 | Both | POTS symptoms |
| 3054.335 | 45 | 5471.179 | 51 | IMASIS | 41 to 64 | Both | POTS symptoms |
| 108.0739 | <5 | 0 | 0 | AUSOM | 41 to 64 | Both | POTS symptoms |
| 0 | 0 | 8526.248 | 86 | AUSOM | 41 to 64 | Both | POTS symptoms |
| 40116.45 | 1466 | 0 | 0 | IPCI | 41 to 64 | Both | POTS symptoms |
| 0 | 0 | 319.0335 | 8 | IPCI | 41 to 64 | Both | POTS symptoms |
| 1140.485 | <5 | 13138.64 | 35 | eDOL_CHUM | 41 to 64 | Both | POTS symptoms |
| 8464.331 | 163 | 25736.34 | 472 | CORIVA | 41 to 64 | Both | POTS symptoms |
| 153547.1 | 2739 | 595017.5 | 10588 | CPRDAurum | 41 to 64 | Both | POTS symptoms |
| 13520.25 | 200 | 44578.61 | 689 | CPRDGOLD | 65 to 150 | Both | POTS symptoms |
| 2926.502 | 36 | 6781.818 | 95 | IMASIS | 65 to 150 | Both | POTS symptoms |
| 77.27858 | <5 | 0 | 0 | AUSOM | 65 to 150 | Both | POTS symptoms |
| 0 | 0 | 6545.029 | 56 | AUSOM | 65 to 150 | Both | POTS symptoms |
| 12758.77 | 549 | 362.8884 | 17 | IPCI | 65 to 150 | Both | POTS symptoms |
| 1238.598 | 11 | 12751.24 | 88 | eDOL_CHUM | 65 to 150 | Both | POTS symptoms |
| 2695.337 | 62 | 11030.15 | 267 | CORIVA | 65 to 150 | Both | POTS symptoms |
| 38517.81 | 815 | 171165 | 3712 | CPRDAurum | 65 to 150 | Both | POTS symptoms |
| 10633.34 | 54 | 21354.38 | 89 | CPRDGOLD | 7 to 11 | Both | POTS symptoms |
| 75.99452 | <5 | 169.2676 | <5 | IMASIS | 7 to 11 | Both | POTS symptoms |
| 19.77823 | <5 | 0 | 0 | AUSOM | 7 to 11 | Both | POTS symptoms |
| 0 | 0 | 431.1786 | <5 | AUSOM | 7 to 11 | Both | POTS symptoms |
| 6583.83 | 131 | 0 | 0 | IPCI | 7 to 11 | Both | POTS symptoms |
| 0 | 0 | 13.57974 | <5 | IPCI | 7 to 11 | Both | POTS symptoms |
| 63.60849 | <5 | 0 | 0 | eDOL_CHUM | 7 to 11 | Both | POTS symptoms |
| 0 | 0 | 899.0226 | <5 | eDOL_CHUM | 7 to 11 | Both | POTS symptoms |
| 1532.851 | <5 | 0 | 0 | CORIVA | 7 to 11 | Both | POTS symptoms |
| 0 | 0 | 2879.433 | 22 | CORIVA | 7 to 11 | Both | POTS symptoms |
| 25827.94 | 188 | 114769.9 | 658 | CPRDAurum | 7 to 11 | Both | POTS symptoms |
| 147254.6 | 40 | 364997.6 | 123 | CPRDGOLD | 0 to 150 | Both | RA |
| 7745.15 | 7 | 16402.12 | 9 | IMASIS | 0 to 150 | Both | RA |
| 0 | 0 | 20786.25 | 24 | AUSOM | 0 to 150 | Both | RA |
| 114130.6 | 65 | 953.629 | <5 | IPCI | 0 to 150 | Both | RA |
| 0 | 0 | 49184.26 | 17 | eDOL_CHUM | 0 to 150 | Both | RA |
| 26838.16 | 25 | 75862.4 | 94 | CORIVA | 0 to 150 | Both | RA |
| 475721 | 170 | 1822633 | 707 | CPRDAurum | 0 to 150 | Both | RA |
| 79647.43 | 33 | 203388.6 | 88 | CPRDGOLD | 0 to 150 | Female | RA |
| 4064.747 | 5 | 9029.276 | <5 | IMASIS | 0 to 150 | Female | RA |
| 0 | 0 | 11072.19 | 12 | AUSOM | 0 to 150 | Female | RA |
| 60800.74 | 36 | 588.2519 | <5 | IPCI | 0 to 150 | Female | RA |
| 0 | 0 | 27532.26 | 8 | eDOL_CHUM | 0 to 150 | Female | RA |
| 14426.42 | 19 | 40519.17 | 65 | CORIVA | 0 to 150 | Female | RA |
| 251387 | 122 | 989186.4 | 512 | CPRDAurum | 0 to 150 | Female | RA |
| 67607.18 | 7 | 161609 | 35 | CPRDGOLD | 0 to 150 | Male | RA |
| 3680.402 | <5 | 7372.846 | 5 | IMASIS | 0 to 150 | Male | RA |
| 0 | 0 | 9714.062 | 12 | AUSOM | 0 to 150 | Male | RA |
| 53329.87 | 29 | 0 | 0 | IPCI | 0 to 150 | Male | RA |
| 0 | 0 | 365.3771 | <5 | IPCI | 0 to 150 | Male | RA |
| 0 | 0 | 21651.99 | 9 | eDOL_CHUM | 0 to 150 | Male | RA |
| 12411.75 | 6 | 35343.23 | 29 | CORIVA | 0 to 150 | Male | RA |
| 224334.1 | 48 | 833447 | 195 | CPRDAurum | 0 to 150 | Male | RA |
| 5429.092 | <5 | 30446.29 | <5 | CPRDGOLD | 0 to 6 | Both | RA |
| 85.78782 | <5 | 0 | 0 | IMASIS | 0 to 6 | Both | RA |
| 0 | 0 | 392.1424 | <5 | IMASIS | 0 to 6 | Both | RA |
| 0 | 0 | 494.5845 | <5 | AUSOM | 0 to 6 | Both | RA |
| 2902.144 | <5 | 0 | 0 | IPCI | 0 to 6 | Both | RA |
| 0 | 0 | 9.336071 | <5 | IPCI | 0 to 6 | Both | RA |
| 0 | 0 | 2503.458 | <5 | eDOL_CHUM | 0 to 6 | Both | RA |
| 1393.837 | <5 | 0 | 0 | CORIVA | 0 to 6 | Both | RA |
| 0 | 0 | 1916.753 | <5 | CORIVA | 0 to 6 | Both | RA |
| 14560.78 | 0 | 138818.3 | 0 | CPRDAurum | 0 to 6 | Both | RA |
| 15592.04 | <5 | 26163.9 | <5 | CPRDGOLD | 12 to 18 | Both | RA |
| 154.6995 | <5 | 0 | 0 | IMASIS | 12 to 18 | Both | RA |
| 0 | 0 | 271.6112 | <5 | IMASIS | 12 to 18 | Both | RA |
| 0 | 0 | 476.1177 | <5 | AUSOM | 12 to 18 | Both | RA |
| 12814.76 | <5 | 0 | 0 | IPCI | 12 to 18 | Both | RA |
| 0 | 0 | 51.27447 | <5 | IPCI | 12 to 18 | Both | RA |
| 0 | 0 | 1940.92 | <5 | eDOL_CHUM | 12 to 18 | Both | RA |
| 2642.79 | <5 | 5883.89 | <5 | CORIVA | 12 to 18 | Both | RA |
| 56551.28 | 0 | 155740.5 | 0 | CPRDAurum | 12 to 18 | Both | RA |
| 49734.56 | 10 | 113637.9 | 23 | CPRDGOLD | 19 to 40 | Both | RA |
| 1327.08 | <5 | 3070.251 | <5 | IMASIS | 19 to 40 | Both | RA |
| 0 | 0 | 3938.672 | <5 | AUSOM | 19 to 40 | Both | RA |
| 35597.97 | 13 | 0 | 0 | IPCI | 19 to 40 | Both | RA |
| 0 | 0 | 174.5435 | <5 | IPCI | 19 to 40 | Both | RA |
| 0 | 0 | 17179.61 | <5 | eDOL_CHUM | 19 to 40 | Both | RA |
| 9800.441 | <5 | 27106.22 | 15 | CORIVA | 19 to 40 | Both | RA |
| 177528.7 | 30 | 611540.1 | 137 | CPRDAurum | 19 to 40 | Both | RA |
| 49463.67 | 21 | 121965.6 | 56 | CPRDGOLD | 41 to 64 | Both | RA |
| 3064.46 | <5 | 5483.767 | <5 | IMASIS | 41 to 64 | Both | RA |
| 0 | 0 | 8549.969 | 12 | AUSOM | 41 to 64 | Both | RA |
| 40606.49 | 35 | 0 | 0 | IPCI | 41 to 64 | Both | RA |
| 0 | 0 | 320.5695 | <5 | IPCI | 41 to 64 | Both | RA |
| 0 | 0 | 13149.86 | <5 | eDOL_CHUM | 41 to 64 | Both | RA |
| 8517.076 | 15 | 25880.88 | 48 | CORIVA | 41 to 64 | Both | RA |
| 154449.2 | 90 | 598538.9 | 384 | CPRDAurum | 41 to 64 | Both | RA |
| 13577.62 | 7 | 44780.56 | 42 | CPRDGOLD | 65 to 150 | Both | RA |
| 2934.434 | 5 | 6805.021 | 5 | IMASIS | 65 to 150 | Both | RA |
| 0 | 0 | 6558.861 | 9 | AUSOM | 65 to 150 | Both | RA |
| 12923.32 | 17 | 368.7337 | <5 | IPCI | 65 to 150 | Both | RA |
| 0 | 0 | 12776.68 | 11 | eDOL_CHUM | 65 to 150 | Both | RA |
| 2713.895 | 6 | 11108.11 | 31 | CORIVA | 65 to 150 | Both | RA |
| 38764.72 | 49 | 172295.8 | 180 | CPRDAurum | 65 to 150 | Both | RA |
| 10645.91 | <5 | 21380.52 | <5 | CPRDGOLD | 7 to 11 | Both | RA |
| 75.99452 | <5 | 169.2676 | <5 | IMASIS | 7 to 11 | Both | RA |
| 0 | 0 | 431.718 | <5 | AUSOM | 7 to 11 | Both | RA |
| 6624.583 | <5 | 0 | 0 | IPCI | 7 to 11 | Both | RA |
| 0 | 0 | 13.57974 | <5 | IPCI | 7 to 11 | Both | RA |
| 0 | 0 | 899.0226 | <5 | eDOL_CHUM | 7 to 11 | Both | RA |
| 1534.355 | <5 | 0 | 0 | CORIVA | 7 to 11 | Both | RA |
| 0 | 0 | 2884.126 | <5 | CORIVA | 7 to 11 | Both | RA |
| 25871.44 | 0 | 114966.1 | 0 | CPRDAurum | 7 to 11 | Both | RA |
| 147265.3 | <5 | 365032.7 | 15 | CPRDGOLD | 0 to 150 | Both | SLE |
| 7746.5 | <5 | 16404.06 | <5 | IMASIS | 0 to 150 | Both | SLE |
| 0 | 0 | 20793.97 | 8 | AUSOM | 0 to 150 | Both | SLE |
| 3941.979 | <5 | 49188.39 | 5 | eDOL_CHUM | 0 to 150 | Both | SLE |
| 0 | 0 | 75891.99 | 7 | CORIVA | 0 to 150 | Both | SLE |
| 475772 | 12 | 1822852 | 57 | CPRDAurum | 0 to 150 | Both | SLE |
| 79657.13 | <5 | 203411.3 | 13 | CPRDGOLD | 0 to 150 | Female | SLE |
| 4065.706 | <5 | 9029.498 | <5 | IMASIS | 0 to 150 | Female | SLE |
| 0 | 0 | 11073.57 | 7 | AUSOM | 0 to 150 | Female | SLE |
| 2229.309 | <5 | 27533.69 | 5 | eDOL_CHUM | 0 to 150 | Female | SLE |
| 0 | 0 | 40537.8 | 6 | CORIVA | 0 to 150 | Female | SLE |
| 251422 | 10 | 989339.3 | 52 | CPRDAurum | 0 to 150 | Female | SLE |
| 67608.17 | <5 | 161621.4 | <5 | CPRDGOLD | 0 to 150 | Male | SLE |
| 3680.794 | <5 | 7374.565 | <5 | IMASIS | 0 to 150 | Male | SLE |
| 0 | 0 | 9720.402 | <5 | AUSOM | 0 to 150 | Male | SLE |
| 1712.671 | <5 | 21654.71 | <5 | eDOL_CHUM | 0 to 150 | Male | SLE |
| 0 | 0 | 35354.19 | <5 | CORIVA | 0 to 150 | Male | SLE |
| 224350 | <5 | 833513.1 | 5 | CPRDAurum | 0 to 150 | Male | SLE |
| 5429.092 | <5 | 30446.29 | <5 | CPRDGOLD | 0 to 6 | Both | SLE |
| 85.78782 | <5 | 0 | 0 | IMASIS | 0 to 6 | Both | SLE |
| 0 | 0 | 392.1424 | <5 | IMASIS | 0 to 6 | Both | SLE |
| 0 | 0 | 494.5845 | <5 | AUSOM | 0 to 6 | Both | SLE |
| 173.8617 | <5 | 0 | 0 | eDOL_CHUM | 0 to 6 | Both | SLE |
| 0 | 0 | 2503.458 | <5 | eDOL_CHUM | 0 to 6 | Both | SLE |
| 0 | 0 | 1916.753 | <5 | CORIVA | 0 to 6 | Both | SLE |
| 14560.78 | 0 | 138818.3 | 0 | CPRDAurum | 0 to 6 | Both | SLE |
| 15592.04 | <5 | 26163.9 | <5 | CPRDGOLD | 12 to 18 | Both | SLE |
| 154.6995 | <5 | 0 | 0 | IMASIS | 12 to 18 | Both | SLE |
| 0 | 0 | 271.6112 | <5 | IMASIS | 12 to 18 | Both | SLE |
| 0 | 0 | 475.373 | <5 | AUSOM | 12 to 18 | Both | SLE |
| 104.7803 | <5 | 1940.999 | <5 | eDOL_CHUM | 12 to 18 | Both | SLE |
| 0 | 0 | 5883.89 | <5 | CORIVA | 12 to 18 | Both | SLE |
| 56551.28 | 0 | 155740.5 | <5 | CPRDAurum | 12 to 18 | Both | SLE |
| 49737.98 | <5 | 113644.2 | 5 | CPRDGOLD | 19 to 40 | Both | SLE |
| 1326.782 | <5 | 3070.338 | <5 | IMASIS | 19 to 40 | Both | SLE |
| 0 | 0 | 3939.696 | <5 | AUSOM | 19 to 40 | Both | SLE |
| 1168.909 | <5 | 17178.95 | <5 | eDOL_CHUM | 19 to 40 | Both | SLE |
| 0 | 0 | 27112.16 | <5 | CORIVA | 19 to 40 | Both | SLE |
| 177536 | 7 | 611573.7 | 26 | CPRDAurum | 19 to 40 | Both | SLE |
| 49469.71 | <5 | 121982.2 | 7 | CPRDGOLD | 41 to 64 | Both | SLE |
| 3064.753 | <5 | 5484.268 | <5 | IMASIS | 41 to 64 | Both | SLE |
| 0 | 0 | 8553.728 | 5 | AUSOM | 41 to 64 | Both | SLE |
| 1140.485 | <5 | 13149.68 | <5 | eDOL_CHUM | 41 to 64 | Both | SLE |
| 0 | 0 | 25893.36 | 5 | CORIVA | 41 to 64 | Both | SLE |
| 154477.1 | <5 | 598657.6 | 27 | CPRDAurum | 41 to 64 | Both | SLE |
| 13578.31 | <5 | 44792.94 | <5 | CPRDGOLD | 65 to 150 | Both | SLE |
| 2935.789 | <5 | 6806.374 | <5 | IMASIS | 65 to 150 | Both | SLE |
| 0 | 0 | 6561.936 | <5 | AUSOM | 65 to 150 | Both | SLE |
| 1243.028 | <5 | 12781.15 | <5 | eDOL_CHUM | 65 to 150 | Both | SLE |
| 0 | 0 | 11119.12 | <5 | CORIVA | 65 to 150 | Both | SLE |
| 38780.33 | <5 | 172360.8 | <5 | CPRDAurum | 65 to 150 | Both | SLE |
| 10645.91 | <5 | 21379.82 | <5 | CPRDGOLD | 7 to 11 | Both | SLE |
| 75.99452 | <5 | 169.2676 | <5 | IMASIS | 7 to 11 | Both | SLE |
| 0 | 0 | 431.718 | <5 | AUSOM | 7 to 11 | Both | SLE |
| 63.60849 | <5 | 0 | 0 | eDOL_CHUM | 7 to 11 | Both | SLE |
| 0 | 0 | 899.0226 | <5 | eDOL_CHUM | 7 to 11 | Both | SLE |
| 0 | 0 | 2884.126 | <5 | CORIVA | 7 to 11 | Both | SLE |
| 25871.44 | 0 | 114965.7 | <5 | CPRDAurum | 7 to 11 | Both | SLE |
| 147259.5 | 26 | 365002.8 | 104 | CPRDGOLD | 0 to 150 | Both | DM |
| 7747.08 | <5 | 16402.72 | 9 | IMASIS | 0 to 150 | Both | DM |
| 0 | 0 | 20795.2 | 7 | AUSOM | 0 to 150 | Both | DM |
| 114149.8 | 17 | 0 | 0 | IPCI | 0 to 150 | Both | DM |
| 3940.517 | <5 | 49179.98 | 27 | eDOL_CHUM | 0 to 150 | Both | DM |
| 26843.92 | 8 | 75887.41 | 23 | CORIVA | 0 to 150 | Both | DM |
| 475744 | 98 | 1822725 | 444 | CPRDAurum | 0 to 150 | Both | DM |
| 79654 | 11 | 203400.4 | 50 | CPRDGOLD | 0 to 150 | Female | DM |
| 4065.996 | <5 | 9029.103 | <5 | IMASIS | 0 to 150 | Female | DM |
| 0 | 0 | 11075.7 | <5 | AUSOM | 0 to 150 | Female | DM |
| 60812.33 | 8 | 0 | 0 | IPCI | 0 to 150 | Female | DM |
| 2227.847 | <5 | 27530.31 | 12 | eDOL_CHUM | 0 to 150 | Female | DM |
| 14432.86 | <5 | 40537.26 | 9 | CORIVA | 0 to 150 | Female | DM |
| 251411.2 | 46 | 989284.2 | 216 | CPRDAurum | 0 to 150 | Female | DM |
| 67605.46 | 15 | 161602.4 | 54 | CPRDGOLD | 0 to 150 | Male | DM |
| 3681.084 | <5 | 7373.615 | 5 | IMASIS | 0 to 150 | Male | DM |
| 0 | 0 | 9719.496 | <5 | AUSOM | 0 to 150 | Male | DM |
| 53337.47 | 9 | 0 | 0 | IPCI | 0 to 150 | Male | DM |
| 1712.671 | <5 | 21649.67 | 15 | eDOL_CHUM | 0 to 150 | Male | DM |
| 12411.06 | 5 | 35350.15 | 14 | CORIVA | 0 to 150 | Male | DM |
| 224332.8 | 52 | 833440.3 | 228 | CPRDAurum | 0 to 150 | Male | DM |
| 5429.092 | <5 | 30441.43 | 11 | CPRDGOLD | 0 to 6 | Both | DM |
| 85.78782 | <5 | 0 | 0 | IMASIS | 0 to 6 | Both | DM |
| 0 | 0 | 392.1424 | <5 | IMASIS | 0 to 6 | Both | DM |
| 0 | 0 | 494.5845 | <5 | AUSOM | 0 to 6 | Both | DM |
| 2902.144 | <5 | 0 | 0 | IPCI | 0 to 6 | Both | DM |
| 173.8617 | <5 | 0 | 0 | eDOL_CHUM | 0 to 6 | Both | DM |
| 0 | 0 | 2503.458 | <5 | eDOL_CHUM | 0 to 6 | Both | DM |
| 1393.837 | <5 | 0 | 0 | CORIVA | 0 to 6 | Both | DM |
| 0 | 0 | 1916.753 | <5 | CORIVA | 0 to 6 | Both | DM |
| 14559.51 | <5 | 138808.4 | 31 | CPRDAurum | 0 to 6 | Both | DM |
| 15591.1 | <5 | 26158.59 | 16 | CPRDGOLD | 12 to 18 | Both | DM |
| 154.6995 | <5 | 0 | 0 | IMASIS | 12 to 18 | Both | DM |
| 0 | 0 | 271.6112 | <5 | IMASIS | 12 to 18 | Both | DM |
| 0 | 0 | 476.0329 | <5 | AUSOM | 12 to 18 | Both | DM |
| 12814.76 | <5 | 0 | 0 | IPCI | 12 to 18 | Both | DM |
| 104.7803 | <5 | 1940.441 | <5 | eDOL_CHUM | 12 to 18 | Both | DM |
| 2642.79 | <5 | 5883.288 | <5 | CORIVA | 12 to 18 | Both | DM |
| 56548.58 | 11 | 155720.3 | 60 | CPRDAurum | 12 to 18 | Both | DM |
| 49736.55 | 6 | 113637.5 | 28 | CPRDGOLD | 19 to 40 | Both | DM |
| 1327.08 | <5 | 3070.283 | <5 | IMASIS | 19 to 40 | Both | DM |
| 0 | 0 | 3939.198 | <5 | AUSOM | 19 to 40 | Both | DM |
| 35600.99 | 8 | 0 | 0 | IPCI | 19 to 40 | Both | DM |
| 1168.676 | <5 | 17177.51 | 5 | eDOL_CHUM | 19 to 40 | Both | DM |
| 9801.292 | <5 | 27111.15 | 5 | CORIVA | 19 to 40 | Both | DM |
| 177527.7 | 30 | 611529 | 160 | CPRDAurum | 19 to 40 | Both | DM |
| 49468.53 | 7 | 121974.9 | 27 | CPRDGOLD | 41 to 64 | Both | DM |
| 3065.035 | <5 | 5483.888 | 5 | IMASIS | 41 to 64 | Both | DM |
| 0 | 0 | 8555.113 | <5 | AUSOM | 41 to 64 | Both | DM |
| 40617.61 | <5 | 0 | 0 | IPCI | 41 to 64 | Both | DM |
| 1139.756 | <5 | 13148.56 | <5 | eDOL_CHUM | 41 to 64 | Both | DM |
| 8520.597 | <5 | 25892.43 | 8 | CORIVA | 41 to 64 | Both | DM |
| 154470.1 | 27 | 598633.3 | 101 | CPRDAurum | 41 to 64 | Both | DM |
| 13578.31 | <5 | 44790.86 | 9 | CPRDGOLD | 65 to 150 | Both | DM |
| 2935.789 | <5 | 6805.692 | <5 | IMASIS | 65 to 150 | Both | DM |
| 0 | 0 | 6561.615 | <5 | AUSOM | 65 to 150 | Both | DM |
| 12929.21 | <5 | 0 | 0 | IPCI | 65 to 150 | Both | DM |
| 1242.527 | <5 | 12775.86 | 17 | eDOL_CHUM | 65 to 150 | Both | DM |
| 2715.28 | <5 | 11117.88 | 5 | CORIVA | 65 to 150 | Both | DM |
| 38776.3 | 10 | 172353 | 27 | CPRDAurum | 65 to 150 | Both | DM |
| 10643.86 | 9 | 21377.18 | 12 | CPRDGOLD | 7 to 11 | Both | DM |
| 75.99452 | <5 | 169.2676 | <5 | IMASIS | 7 to 11 | Both | DM |
| 0 | 0 | 431.718 | <5 | AUSOM | 7 to 11 | Both | DM |
| 6623.745 | <5 | 0 | 0 | IPCI | 7 to 11 | Both | DM |
| 63.60849 | <5 | 0 | 0 | eDOL_CHUM | 7 to 11 | Both | DM |
| 0 | 0 | 899.0226 | <5 | eDOL_CHUM | 7 to 11 | Both | DM |
| 1534.355 | <5 | 0 | 0 | CORIVA | 7 to 11 | Both | DM |
| 0 | 0 | 2883.608 | <5 | CORIVA | 7 to 11 | Both | DM |
| 25867.91 | 14 | 114948.3 | 53 | CPRDAurum | 7 to 11 | Both | DM |
