## Supplementary File 3 for "The risks of autoimmune- and inflammatory post-acute COVID-19 conditions: a network cohort study in six European countries, the US, and Korea"

### Supplementary File 3. Incidence rate ratio results per database with numeric values

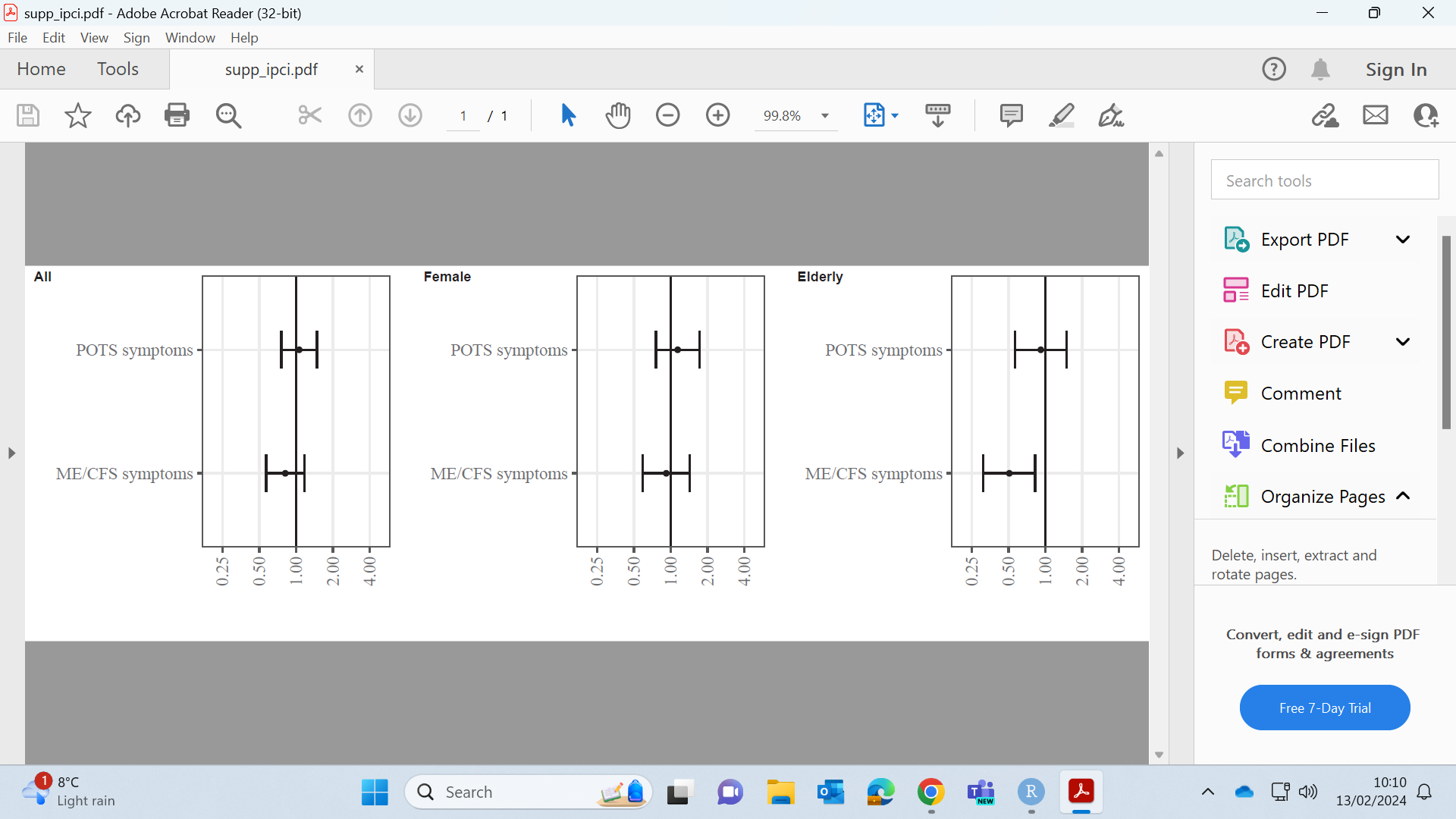

###### Supplementary Figure 1. Incidence rate ratio results in IPCI database.

Numeric values available in Supplementary Table 3.

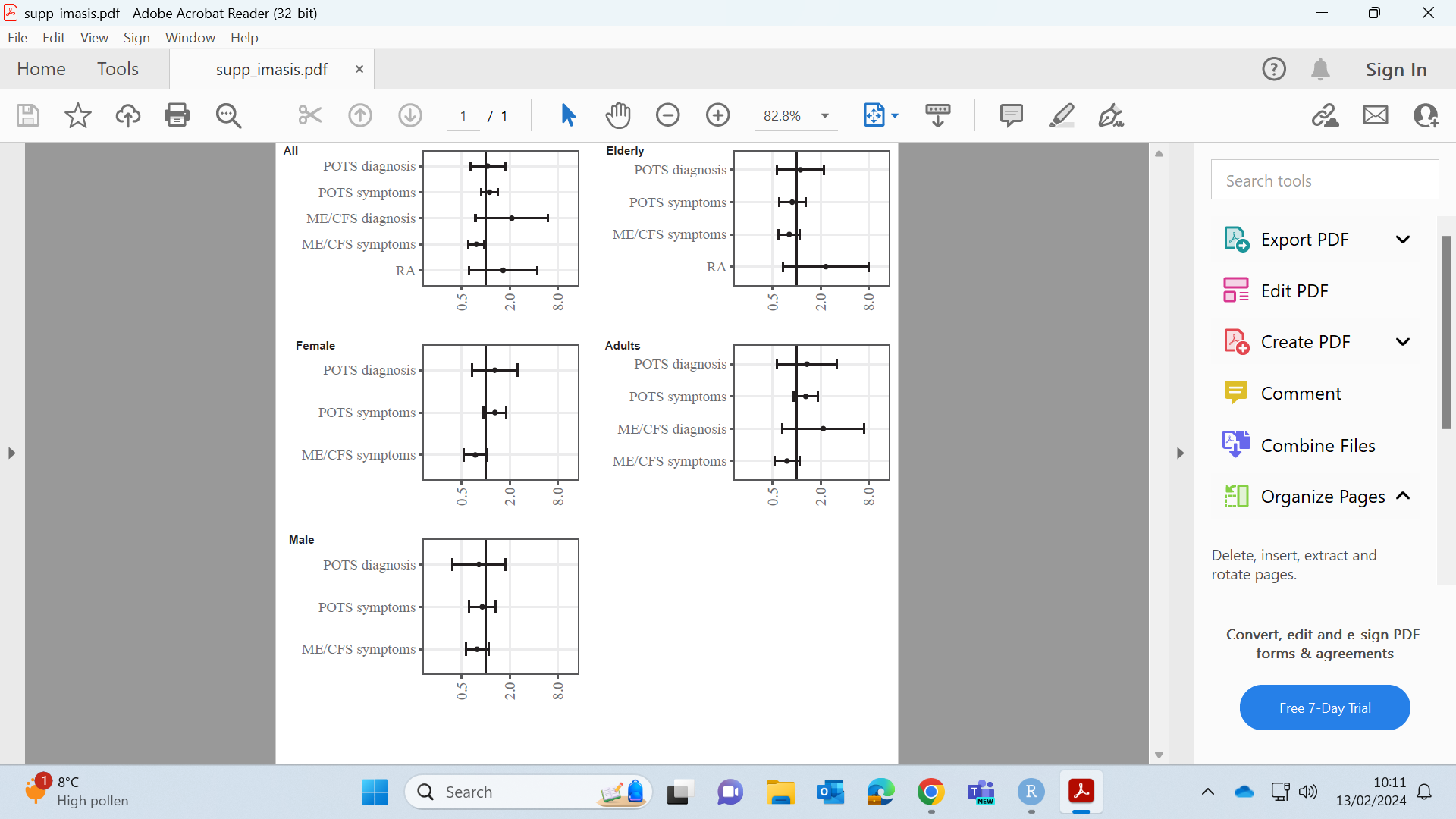

###### Supplementary Figure 2. Incidence rate ratio results in IMASIS database.

Numeric values available in Supplementary Table 3

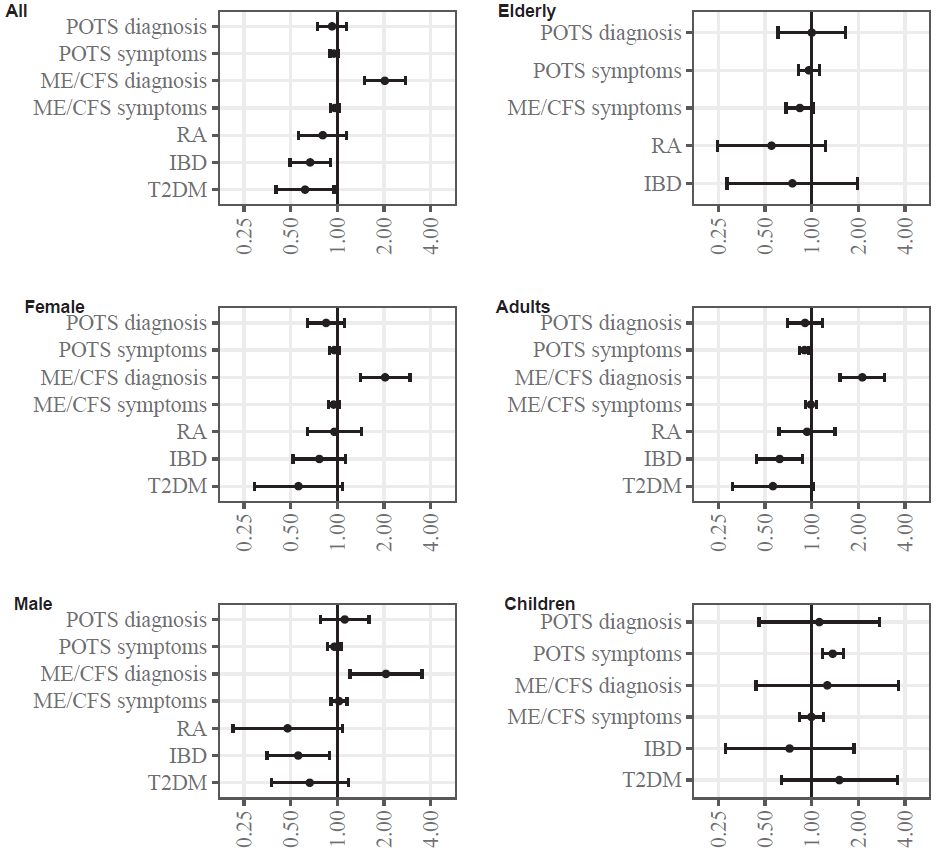

###### Supplementary Figure 3. Incidence rate ratio results in CPRD GOLD database.

Numeric values available in Supplementary Table 3

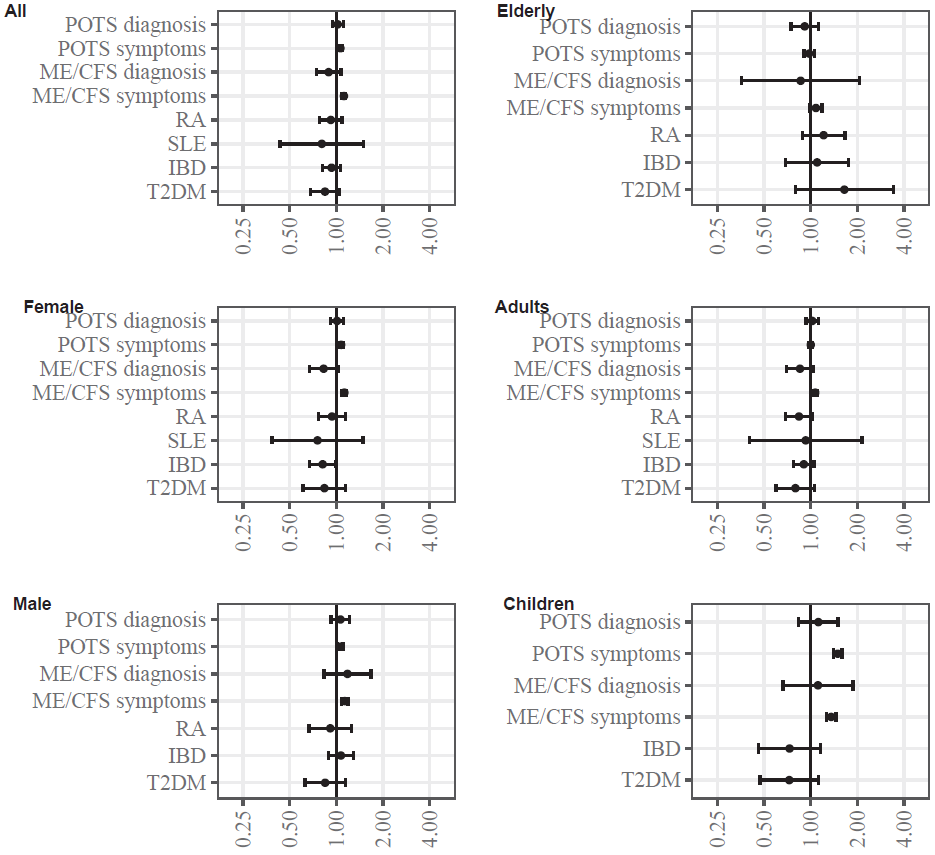

###### Supplementary Figure 4. Incidence rate ratio results in CPRD Aurum database.

Numeric values available in Supplementary Table 3

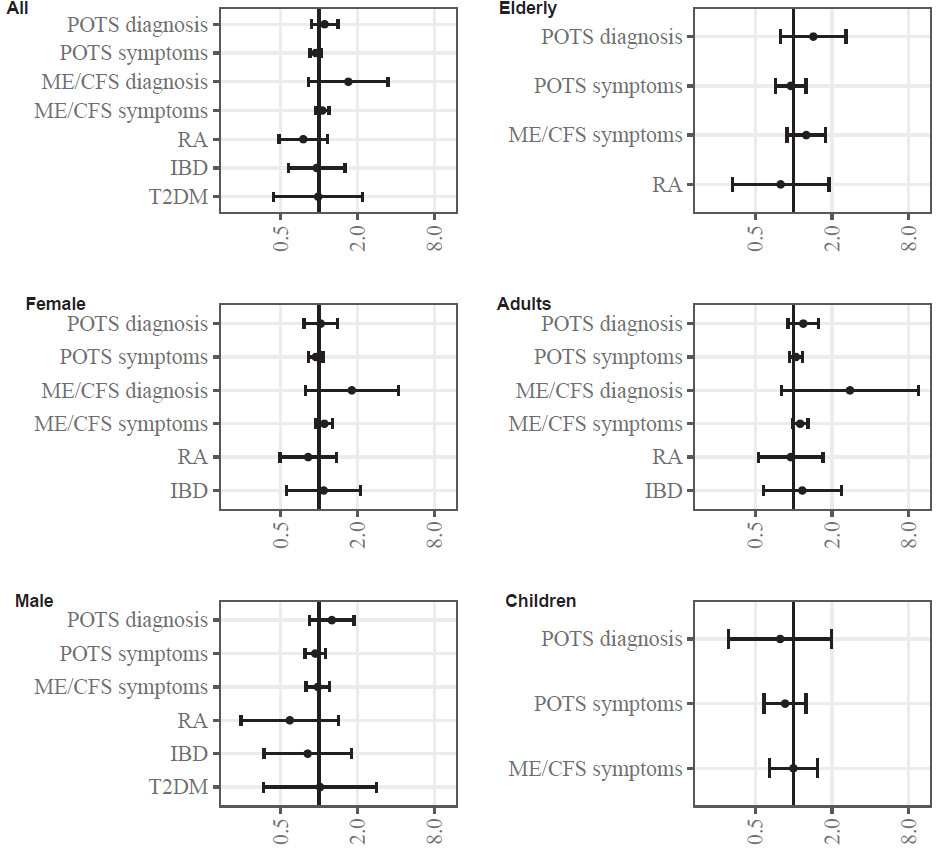

###### Supplementary Figure 5. Incidence rate ratio results in CORIVA database.

Numeric values available in Supplementary Table 3

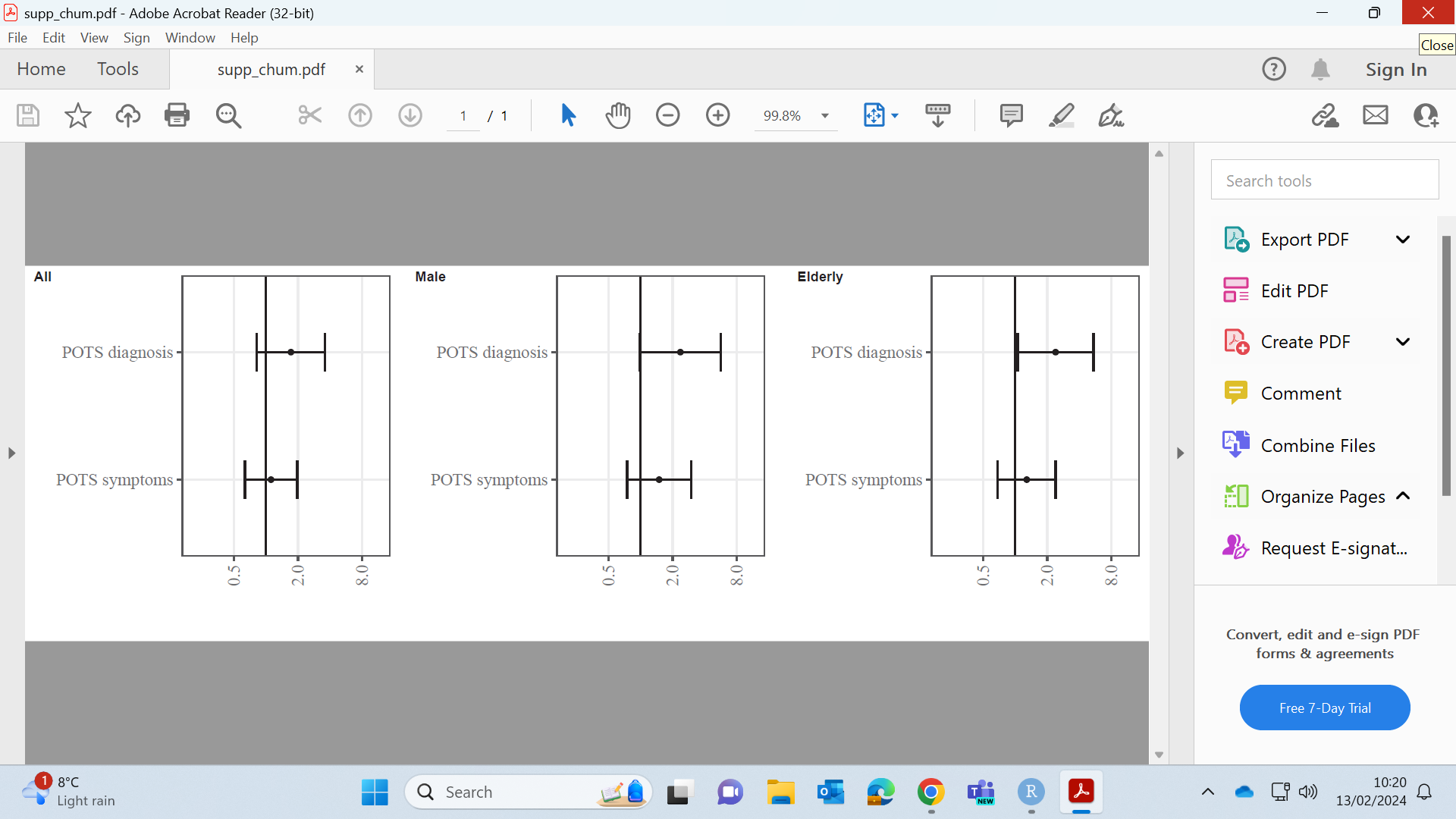

###### Supplementary Figure 6. Incidence rate ratio results in CHUM database.

Numeric values available in Supplementary Table 3

AUSOM had too few counts for individual results.

###### Supplementary Table 3. Numeric values of incidence rate ratios with 95% confidence intervals corresponding to Supplementary Figures 1-6

| **Outcomes per database** | **All** | **Female** | **Male** | **Elderly**  **(>64 years** | **Adults**  **(19-64 years)** | **Children**  **(<19 years)** |
| --- | --- | --- | --- | --- | --- | --- |
| **IPCI** |  |  |  |  |  |  |
| POTS symptoms | 1.06 (0.76-1.49) | 1.14 (0.76-1.72) | NA | 0.92 (0.57-1.49) | NA | NA |
| ME/CFS symptoms | 0.82 (0.57-1.17) | 0.92 (0.59-1.43) | NA | 0.51 (0.31-0.83) | NA | NA |
| **IMASIS** |  |  |  |  |  |  |
| POTS symptoms | 1.11 (0.87-1.42) | 1.3 (0.94-1.8) | 1.3 (0.94-1.8) | 0.88 (0.6-1.29) | 1.3 (0.92-1.83) | NA |
| ME/CFS diagnosis | 2.12 (0.74-6.04) | NA | NA | NA | 2.15 (0.66-7.04) | NA |
| ME/CFS symptoms | 0.76 (0.6-0.96) | 0.74 (0.53-1.04) | 0.74 (0.53-1.04) | 0.81 (0.59-1.1) | 0.76 (0.52-1.09) | NA |
| RA | 1.65 (0.61-4.42) | NA | NA | 2.32 (0.67-8.01) | NA | NA |
| POTS diagnosis | 1.06 (0.64-1.75) | 1.3 (0.67-2.5) | 1.3 (0.67-2.5) | 1.11 (0.56-2.21) | 1.34 (0.57-3.18) | NA |
| **CPRD GOLD** |  |  |  |  |  |  |
| POTS symptoms | 0.95 (0.9-1.01) | 0.96 (0.89-1.03) | 0.96 (0.87-1.06) | 0.96 (0.82-1.12) | 0.9 (0.84-0.96) | 1.36 (1.17-1.59) |
| IBD | 0.67 (0.5-0.9) | 0.77 (0.52-1.13) | 0.56 (0.35-0.89) | 0.75 (0.28-1.98) | 0.62 (0.44-0.87) | 0.72 (0.28-1.87) |
| ME/CFS diagnosis | 2.03 (1.5-2.74) | 2.04 (1.41-2.94) | 2.06 (1.21-3.52) | NA | 2.12 (1.52-2.95) | 1.26 (0.44-3.63) |
| ME/CFS symptoms | 0.96 (0.9-1.03) | 0.95 (0.87-1.03) | 1.03 (0.91-1.16) | 0.84 (0.68-1.03) | 0.99 (0.91-1.07) | 0.99 (0.84-1.18) |
| RA | 0.81 (0.56-1.15) | 0.96 (0.64-1.43) | 0.48 (0.21-1.08) | 0.55 (0.25-1.22) | 0.93 (0.62-1.41) | NA |
| POTS diagnosis | 0.93 (0.74-1.15) | 0.85 (0.65-1.11) | 1.12 (0.78-1.6) | 1 (0.61-1.65) | 0.9 (0.7-1.17) | 1.12 (0.46-2.74) |
| DM | 0.62 (0.4-0.95) | 0.56 (0.29-1.08) | 0.66 (0.37-1.18) | NA | 0.56 (0.31-1.03) | 1.51 (0.63-3.57) |
| **CPRD Aurum** |  |  |  |  |  |  |
| POTS symptoms | 1.06 (1.04-1.09) | 1.07 (1.04-1.11) | 1.06 (1.02-1.11) | 0.98 (0.9-1.05) | 1 (0.97-1.03) | 1.49 (1.4-1.59) |
| IBD | 0.93 (0.82-1.07) | 0.82 (0.67-0.99) | 1.07 (0.89-1.29) | 1.1 (0.69-1.75) | 0.9 (0.78-1.05) | 0.73 (0.46-1.16) |
| ME/CFS diagnosis | 0.89 (0.74-1.07) | 0.83 (0.67-1.03) | 1.18 (0.83-1.68) | 0.86 (0.36-2.06) | 0.85 (0.7-1.04) | 1.11 (0.66-1.87) |
| ME/CFS symptoms | 1.12 (1.09-1.15) | 1.13 (1.09-1.17) | 1.13 (1.08-1.19) | 1.08 (0.98-1.18) | 1.07 (1.04-1.1) | 1.35 (1.26-1.45) |
| RA | 0.92 (0.78-1.09) | 0.94 (0.77-1.14) | 0.91 (0.67-1.25) | 1.21 (0.88-1.66) | 0.84 (0.69-1.02) | NA |
| SLE | 0.81 (0.43-1.5) | 0.76 (0.38-1.49) | NA | NA | 0.93 (0.4-2.14) | NA |
| POTS diagnosis | 1.02 (0.94-1.11) | 1.01 (0.91-1.12) | 1.06 (0.93-1.22) | 0.91 (0.74-1.12) | 1.02 (0.93-1.12) | 1.12 (0.84-1.5) |
| DM | 0.85 (0.68-1.05) | 0.84 (0.61-1.15) | 0.85 (0.63-1.15) | 1.65 (0.8-3.4) | 0.8 (0.6-1.06) | 0.73 (0.47-1.12) |
| **CORIVA** |  |  |  |  |  |  |
| POTS symptoms | 0.94 (0.85-1.05) | 0.94 (0.83-1.07) | 0.93 (0.78-1.12) | 0.95 (0.72-1.25) | 1.04 (0.92-1.18) | 0.86 (0.59-1.25) |
| IBD | 0.96 (0.58-1.59) | 1.09 (0.56-2.12) | 0.81 (0.37-1.79) | NA | 1.17 (0.58-2.37) | NA |
| ME/CFS diagnosis | 1.7 (0.83-3.47) | 1.81 (0.78-4.17) | NA | NA | 2.77 (0.8-9.56) | NA |
| ME/CFS symptoms | 1.06 (0.94-1.2) | 1.1 (0.95-1.28) | 0.98 (0.79-1.21) | 1.26 (0.89-1.78) | 1.13 (0.98-1.3) | 1 (0.65-1.53) |
| RA | 0.75 (0.48-1.17) | 0.82 (0.49-1.37) | 0.59 (0.24-1.42) | 0.79 (0.33-1.9) | 0.95 (0.53-1.7) | NA |
| POTS diagnosis | 1.1 (0.87-1.4) | 1.03 (0.76-1.39) | 1.26 (0.84-1.88) | 1.43 (0.79-2.57) | 1.19 (0.9-1.57) | 0.79 (0.31-1.99) |
| DM | 0.98 (0.44-2.2) | NA | 1.02 (0.37-2.82) | NA | NA | NA |
| **CHUM** |  |  |  |  |  |  |
| POTS symptoms | 1.12 (0.63-1.97) | NA | 1.5 (0.75-2.99) | 1.29 (0.69-2.41) | NA | NA |
| POTS diagnosis | 1.72 (0.82-3.61) | NA | 2.37 (0.99-5.67) | 2.4 (1.06-5.47) | NA | NA |

IBD: inflammatory bowel disease; ME/CFS: myalgic encephalomyelitis / chronic fatigues syndrome; NA: results suppressed because less than 5 outcomes; POTS: postural orthostatic tachycardia syndrome; RA: rheumatoid arthritis; SLE: systemic lupus erythematosus; DM: diabetes mellitus
