## Supplementary File 4 for "The risks of autoimmune- and inflammatory post-acute COVID-19 conditions: a network cohort study in six European countries, the US, and Korea"

### Supplementary File 4. Numeric values of incidence rates with 95% confidence intervals corresponding to Figure 2

###### Supplementary Table 4. Numeric values of incidence rates with 95% confidence intervals corresponding to Figure 2

| **outcome** | **# events** | **Person years** | **Incidence**  **(per 100000 pys)** | **95% CI**  **lower bound** | **95% CI**  **upper bound** | **database** | **Care sector** |
| --- | --- | --- | --- | --- | --- | --- | --- |
| POTS symptoms | 532063 | 6945424.159 | 7660.626448 | 7640.056037 | 7681.239 | PharMetrics Plus for Academics | both |
| POTS symptoms | 15890 | 913621.5743 | 1739.232134 | 1712.293649 | 1766.488 | CORIVA | both |
| POTS symptoms | 249779 | 6124302.738 | 4078.48878 | 4062.509797 | 4094.515 | NLHR@UIO | both |
| ME/CFS symptoms | 450065 | 7472824.772 | 6022.68906 | 6005.106266 | 6040.311 | PharMetrics Plus for Academics | both |
| ME/CFS symptoms | 10579 | 943558.371 | 1121.181299 | 1099.916919 | 1142.753 | CORIVA | both |
| ME/CFS symptoms | 161542 | 6580942.448 | 2454.694009 | 2442.738172 | 2466.694 | NLHR@UIO | both |
| POTS diagnosis | 132018 | 8937028.496 | 1477.202406 | 1469.244603 | 1485.193 | PharMetrics Plus for Academics | both |
| POTS diagnosis | 2879 | 976222.4805 | 294.9122826 | 284.2370409 | 305.8859 | CORIVA | both |
| POTS diagnosis | 6465 | 7294946.672 | 88.62299192 | 86.47572384 | 90.8101 | NLHR@UIO | both |
| ME/CFS diagnosis | 43679 | 9221913.062 | 473.643589 | 469.2120234 | 478.1066 | PharMetrics Plus for Academics | both |
| ME/CFS diagnosis | 662 | 988757.1417 | 66.95274017 | 61.9489949 | 72.25304 | CORIVA | both |
| ME/CFS diagnosis | 1495 | 7329130.012 | 20.3980554 | 19.37704876 | 21.4589 | NLHR@UIO | both |
| MIS | 417 | 9386691.001 | 4.442460074 | 4.026251735 | 4.89001 | PharMetrics Plus for Academics | both |
| MIS | 161 | 990314.7269 | 16.25745792 | 13.843186 | 18.9717 | CORIVA | both |
| MIS | 51 | 7330945.823 | 0.695681038 | 0.517980227 | 0.914692 | NLHR@UIO | both |
| DM | 6695 | 9324592.151 | 71.79938695 | 70.08970522 | 73.54024 | PharMetrics Plus for Academics | both |
| DM | 4079 | 7271439.247 | 56.09618483 | 54.38775705 | 57.84463 | NLHR@UIO | both |
| DM | 393 | 984917.0595 | 39.90183703 | 36.05388635 | 44.04863 | CORIVA | Both |
| POTS symptoms | 44852 | 4258910 | 1053.133 | 1043.409 | 1062.926 | CPRD GOLD | primary care |
| POTS symptoms | 90826 | 1992188 | 4559.109 | 4529.506 | 4588.856 | IPCI | primary care |
| POTS symptoms | 254753 | 14576245 | 1747.727 | 1740.947 | 1754.527 | CPRD Aurum | primary care |
| ME/CFS symptoms | 28989 | 4553029 | 636.697 | 629.3885 | 644.0692 | CPRD GOLD | primary care |
| ME/CFS symptoms | 57867 | 2273981 | 2544.745 | 2524.053 | 2565.564 | IPCI | primary care |
| ME/CFS symptoms | 175410 | 15535365 | 1129.101 | 1123.823 | 1134.398 | CPRD Aurum | primary care |
| POTS diagnosis | 3377 | 5291398 | 63.82056 | 61.68602 | 66.01012 | CPRD GOLD | primary care |
| POTS diagnosis | 2200 | 2739721 | 80.30015 | 76.97938 | 83.72731 | IPCI | primary care |
| POTS diagnosis | 24420 | 18299595 | 133.4456 | 131.777 | 135.1299 | CPRD Aurum | primary care |
| ME/CFS diagnosis | 993 | 5315136 | 18.6825 | 17.53841 | 19.88162 | CPRD GOLD | primary care |
| ME/CFS diagnosis | 433 | 2753401 | 15.726 | 14.27947 | 17.27936 | IPCI | primary care |
| ME/CFS diagnosis | 4672 | 18421361 | 25.36186 | 24.63978 | 26.09973 | CPRD Aurum | primary care |
| MIS | 73 | 18487136 | 0.394869 | 0.309514 | 0.496489 | CPRD Aurum | primary care |
| DM | 1238 | 5309349 | 23.31736 | 22.03642 | 24.65334 | CPRD GOLD | primary care |
| DM | 4282 | 18409910 | 23.25921 | 22.56771 | 23.96651 | CPRD Aurum | primary care |
| DM | 426 | 2751955 | 15.47991 | 14.04464 | 17.02206 | IPCI | primary care |
| POTS symptoms | 2148 | 243901.1 | 880.6849 | 843.8311 | 918.7339 | IMASIS | secondary care |
| POTS symptoms | 5990 | 729342.1 | 821.2881 | 800.6199 | 842.3549 | AUSOM | secondary care |
| POTS symptoms | 1806 | 4416177 | 40.8951 | 39.03056 | 42.8257 | CHUM | secondary care |
| ME/CFS symptoms | 2545 | 244173.3 | 1042.292 | 1002.187 | 1083.591 | IMASIS | secondary care |
| ME/CFS symptoms | 1704 | 762086 | 223.5968 | 213.1052 | 234.4713 | AUSOM | secondary care |
| ME/CFS symptoms | 857 | 4422087 | 19.37999 | 18.10403 | 20.72216 | CHUM | secondary care |
| POTS diagnosis | 680 | 254100.3 | 267.6109 | 247.8723 | 288.5033 | IMASIS | secondary care |
| POTS diagnosis | 458 | 777371.6 | 58.91648 | 53.64358 | 64.56755 | AUSOM | secondary care |
| POTS diagnosis | 791 | 4425447 | 17.8739 | 16.64984 | 19.16415 | CHUM | secondary care |
| ME/CFS diagnosis | 107 | 256319.8 | 41.74472 | 34.21081 | 50.44422 | IMASIS | secondary care |
| ME/CFS diagnosis | 116 | 779815.8 | 14.87531 | 12.29175 | 17.84151 | AUSOM | secondary care |
| ME/CFS diagnosis | 117 | 4432171 | 2.63979 | 2.183178 | 3.163719 | CHUM | secondary care |
| MIS | 6 | 780684.9 | 0.768556 | 0.282046 | 1.672823 | AUSOM | secondary care |
| MIS | 58 | 4433631 | 1.308183 | 0.993358 | 1.69113 | CHUM | secondary care |
| DM | 220 | 255439.8 | 86.12598 | 75.12043 | 98.29019 | IMASIS | secondary care |
| DM | 141 | 777767 | 18.12882 | 15.26008 | 21.38017 | AUSOM | secondary care |
| DM | 366 | 4426368 | 8.26863 | 7.443114 | 9.160682 | CHUM | secondary care |
