## Supplementary File 5 for "The risks of autoimmune- and inflammatory post-acute COVID-19 conditions: a network cohort study in six European countries, the US, and Korea"

### Supplementary File 5. Numeric values of incidence rates with 95% confidence intervals corresponding to Figure 3

###### Supplementary Table 5. Numeric values of incidence rates with 95% confidence intervals corresponding to Figure 3

| **outcome** | **# events** | **Person years** | **Incidence**  **(per 100000 pys)** | **95% CI**  **lower bound** | **95% CI**  **upper bound** | **Year** | **database** | **Care sector** |
| --- | --- | --- | --- | --- | --- | --- | --- | --- |
| POTS symptoms | 107456 | 1412562 | 7607.173 | 7561.756 | 7652.795 | 2020 | P+ | both |
| POTS symptoms | 334045 | 4264583 | 7833.005 | 7806.465 | 7859.614 | 2021 | P+ | both |
| POTS symptoms | 90562 | 1268280 | 7140.539 | 7094.108 | 7187.198 | 2022 | P+ | both |
| POTS symptoms | 2341 | 136300.5 | 1717.529 | 1648.652 | 1788.544 | 2020 | CORIVA | both |
| POTS symptoms | 7116 | 402245.2 | 1769.07 | 1728.203 | 1810.66 | 2021 | CORIVA | both |
| POTS symptoms | 6433 | 375075.8 | 1715.12 | 1673.461 | 1757.553 | 2022 | CORIVA | both |
| POTS symptoms | 76568 | 1565803 | 4890.014 | 4855.438 | 4924.775 | 2020 | NLHR@UIO | both |
| POTS symptoms | 173211 | 4558499 | 3799.737 | 3781.864 | 3817.674 | 2021 | NLHR@UIO | both |
| POTS diagnosis | 25600 | 1797231 | 1424.413 | 1407.017 | 1441.971 | 2020 | P+ | both |
| POTS diagnosis | 82689 | 5508924 | 1501.001 | 1490.788 | 1511.267 | 2021 | P+ | both |
| POTS diagnosis | 23729 | 1630874 | 1454.987 | 1436.532 | 1473.619 | 2022 | P+ | both |
| POTS diagnosis | 448 | 143671.3 | 311.8229 | 283.6131 | 342.0794 | 2020 | CORIVA | both |
| POTS diagnosis | 1289 | 427954.6 | 301.2002 | 284.9798 | 318.1033 | 2021 | CORIVA | both |
| POTS diagnosis | 1142 | 404596.6 | 282.2564 | 266.1214 | 299.114 | 2022 | CORIVA | both |
| POTS diagnosis | 1770 | 1826416 | 96.91109 | 92.44841 | 101.5335 | 2020 | NLHR@UIO | both |
| POTS diagnosis | 4695 | 5468530 | 85.85488 | 83.41643 | 88.34651 | 2021 | NLHR@UIO | both |
| ME/CFS symptoms | 91842 | 1514266 | 6065.119 | 6025.956 | 6104.473 | 2020 | P+ | both |
| ME/CFS symptoms | 283212 | 4593346 | 6165.702 | 6143.015 | 6188.452 | 2021 | P+ | both |
| ME/CFS symptoms | 75011 | 1365214 | 5494.451 | 5455.201 | 5533.914 | 2022 | P+ | both |
| ME/CFS symptoms | 1452 | 139969 | 1037.373 | 984.6948 | 1092.137 | 2020 | CORIVA | both |
| ME/CFS symptoms | 4715 | 414708.4 | 1136.944 | 1104.72 | 1169.868 | 2021 | CORIVA | both |
| ME/CFS symptoms | 4412 | 388881.1 | 1134.537 | 1101.304 | 1168.518 | 2022 | CORIVA | both |
| ME/CFS symptoms | 48298 | 1668493 | 2894.708 | 2868.949 | 2920.641 | 2020 | NLHR@UIO | both |
| ME/CFS symptoms | 113244 | 4912450 | 2305.245 | 2291.838 | 2318.711 | 2021 | NLHR@UIO | both |
| ME/CFS diagnosis | 7514 | 1849880 | 406.1885 | 397.0557 | 415.4785 | 2020 | P+ | both |
| ME/CFS diagnosis | 28326 | 5687980 | 497.9975 | 492.2148 | 503.8312 | 2021 | P+ | both |
| ME/CFS diagnosis | 7839 | 1684053 | 465.4842 | 455.2362 | 475.9048 | 2022 | P+ | both |
| ME/CFS diagnosis | 37 | 145216.3 | 25.47923 | 17.93973 | 35.11977 | 2020 | CORIVA | both |
| ME/CFS diagnosis | 315 | 433182.3 | 72.71764 | 64.90816 | 81.20792 | 2021 | CORIVA | both |
| ME/CFS diagnosis | 310 | 410358.5 | 75.5437 | 67.36743 | 84.43874 | 2022 | CORIVA | both |
| ME/CFS diagnosis | 343 | 1834364 | 18.69858 | 16.77187 | 20.78593 | 2020 | NLHR@UIO | both |
| ME/CFS diagnosis | 1152 | 5494766 | 20.96541 | 19.77207 | 22.21194 | 2021 | NLHR@UIO | both |
| MIS | 327 | 5791495 | 5.64621 | 5.050753 | 6.29257 | 2021 | P+ | both |
| MIS | 89 | 1714089 | 5.192261 | 4.169812 | 6.389524 | 2022 | P+ | both |
| MIS | 20 | 433778.9 | 4.610643 | 2.816301 | 7.120765 | 2021 | CORIVA | both |
| MIS | 141 | 411180.6 | 34.2915 | 28.86514 | 40.44158 | 2022 | CORIVA | both |
| MIS | 12 | 1834625 | 0.654085 | 0.337975 | 1.142554 | 2020 | NLHR@UIO | both |
| MIS | 39 | 5496321 | 0.709566 | 0.504571 | 0.97 | 2021 | NLHR@UIO | both |
| DM | 1418 | 1868528 | 75.88862 | 71.98965 | 79.94388 | 2020 | P+ | both |
| DM | 4201 | 5753017 | 73.02255 | 70.83091 | 75.26477 | 2021 | P+ | both |
| DM | 1076 | 1703047 | 63.18087 | 59.4617 | 67.07174 | 2022 | P+ | both |
| DM | 55 | 144584.6 | 38.04 | 28.65694 | 49.51429 | 2020 | CORIVA | both |
| DM | 180 | 431424.8 | 41.72222 | 35.84956 | 48.28249 | 2021 | CORIVA | both |
| DM | 158 | 408907.6 | 38.63953 | 32.84948 | 45.15624 | 2022 | CORIVA | both |
| DM | 1310 | 1820071 | 71.97522 | 68.12993 | 75.981 | 2020 | NLHR@UIO | both |
| DM | 2769 | 5451368 | 50.79459 | 48.9201 | 52.72251 | 2021 | NLHR@UIO | both |
| POTS symptoms | 7648 | 857089.9 | 892.3218 | 872.4341 | 912.5484 | 2020 | CPRD GOLD | primary care |
| POTS symptoms | 25256 | 2403701 | 1050.713 | 1037.794 | 1063.753 | 2021 | CPRD GOLD | primary care |
| POTS symptoms | 11948 | 998118.6 | 1197.052 | 1175.683 | 1218.712 | 2022 | CPRD GOLD | primary care |
| POTS symptoms | 12127 | 295803.6 | 4099.68 | 4027.035 | 4173.307 | 2020 | IPCI | primary care |
| POTS symptoms | 39732 | 894648.3 | 4441.075 | 4397.512 | 4484.961 | 2021 | IPCI | primary care |
| POTS symptoms | 38967 | 801735.8 | 4860.33 | 4812.19 | 4908.831 | 2022 | IPCI | primary care |
| POTS symptoms | 52565 | 3196137 | 1644.642 | 1630.612 | 1658.762 | 2020 | CPRD Aurum | primary care |
| POTS symptoms | 167199 | 9437807 | 1771.587 | 1763.106 | 1780.1 | 2021 | CPRD Aurum | primary care |
| POTS symptoms | 34989 | 1942301 | 1801.42 | 1782.593 | 1820.396 | 2022 | CPRD Aurum | primary care |
| POTS diagnosis | 622 | 1066934 | 58.29792 | 53.80584 | 63.06492 | 2020 | CPRD GOLD | primary care |
| POTS diagnosis | 1940 | 2986910 | 64.95006 | 62.09171 | 67.90606 | 2021 | CPRD GOLD | primary care |
| POTS diagnosis | 815 | 1237554 | 65.85569 | 61.41141 | 70.5366 | 2022 | CPRD GOLD | primary care |
| POTS diagnosis | 380 | 405244.1 | 93.77064 | 84.57842 | 103.6894 | 2020 | IPCI | primary care |
| POTS diagnosis | 1004 | 1228021 | 81.75756 | 76.77793 | 86.97538 | 2021 | IPCI | primary care |
| POTS diagnosis | 816 | 1106456 | 73.74899 | 68.77503 | 78.98762 | 2022 | IPCI | primary care |
| POTS diagnosis | 4868 | 4004745 | 121.5558 | 118.1648 | 125.0194 | 2020 | CPRD Aurum | primary care |
| POTS diagnosis | 16244 | 11853866 | 137.0355 | 134.9361 | 139.1593 | 2021 | CPRD Aurum | primary care |
| POTS diagnosis | 3308 | 2440984 | 135.5191 | 130.9399 | 140.2176 | 2022 | CPRD Aurum | primary care |
| ME/CFS symptoms | 4718 | 915770.3 | 515.1947 | 500.5976 | 530.1094 | 2020 | CPRD GOLD | primary care |
| ME/CFS symptoms | 16385 | 2570130 | 637.5165 | 627.7919 | 647.354 | 2021 | CPRD GOLD | primary care |
| ME/CFS symptoms | 7886 | 1067129 | 738.9922 | 722.771 | 755.4858 | 2022 | CPRD GOLD | primary care |
| ME/CFS symptoms | 7380 | 336837 | 2190.971 | 2141.266 | 2241.539 | 2020 | IPCI | primary care |
| ME/CFS symptoms | 25238 | 1020193 | 2473.846 | 2443.418 | 2504.557 | 2021 | IPCI | primary care |
| ME/CFS symptoms | 25249 | 916950.6 | 2753.584 | 2719.723 | 2787.761 | 2022 | IPCI | primary care |
| ME/CFS symptoms | 32636 | 3401103 | 959.5711 | 949.1883 | 970.0391 | 2020 | CPRD Aurum | primary care |
| ME/CFS symptoms | 116235 | 10062210 | 1155.164 | 1148.532 | 1161.824 | 2021 | CPRD Aurum | primary care |
| ME/CFS symptoms | 26539 | 2072051 | 1280.808 | 1265.444 | 1296.312 | 2022 | CPRD Aurum | primary care |
| ME/CFS diagnosis | 203 | 1071539 | 18.94472 | 16.42812 | 21.73778 | 2020 | CPRD GOLD | primary care |
| ME/CFS diagnosis | 552 | 3000296 | 18.39818 | 16.89518 | 19.99903 | 2021 | CPRD GOLD | primary care |
| ME/CFS diagnosis | 238 | 1243301 | 19.14259 | 16.78766 | 21.73537 | 2022 | CPRD GOLD | primary care |
| ME/CFS diagnosis | 44 | 407236.6 | 10.80453 | 7.850588 | 14.50458 | 2020 | IPCI | primary care |
| ME/CFS diagnosis | 194 | 1234243 | 15.71814 | 13.58403 | 18.09238 | 2021 | IPCI | primary care |
| ME/CFS diagnosis | 195 | 1111922 | 17.53721 | 15.16201 | 20.17894 | 2022 | IPCI | primary care |
| ME/CFS diagnosis | 1105 | 4030028 | 27.41916 | 25.82612 | 29.08475 | 2020 | CPRD Aurum | primary care |
| ME/CFS diagnosis | 3029 | 11933180 | 25.38301 | 24.48703 | 26.30339 | 2021 | CPRD Aurum | primary care |
| ME/CFS diagnosis | 538 | 2458153 | 21.88635 | 20.07578 | 23.81637 | 2022 | CPRD Aurum | primary care |
| MIS | 25 | 11975945 | 0.208752 | 0.135093 | 0.308159 | 2021 | CPRD Aurum | primary care |
| MIS | 48 | 2467048 | 1.945645 | 1.434565 | 2.579644 | 2022 | CPRD Aurum | primary care |
| DM | 237 | 1070402 | 22.14122 | 19.41185 | 25.14687 | 2020 | CPRD GOLD | primary care |
| DM | 783 | 2997040 | 26.12578 | 24.32764 | 28.02165 | 2021 | CPRD GOLD | primary care |
| DM | 218 | 1241907 | 17.55365 | 15.30066 | 20.04499 | 2022 | CPRD GOLD | primary care |
| DM | 79 | 406987.2 | 19.41093 | 15.36781 | 24.19181 | 2020 | IPCI | primary care |
| DM | 189 | 1233407 | 15.32341 | 13.21659 | 17.67062 | 2021 | IPCI | primary care |
| DM | 158 | 1111561 | 14.21425 | 12.08427 | 16.61154 | 2022 | IPCI | primary care |
| DM | 943 | 4027586 | 23.41353 | 21.94281 | 24.9569 | 2020 | CPRD Aurum | primary care |
| DM | 2857 | 11925738 | 23.95659 | 23.08611 | 24.85149 | 2021 | CPRD Aurum | primary care |
| DM | 482 | 2456586 | 19.62072 | 17.90797 | 21.4531 | 2022 | CPRD Aurum | primary care |
| POTS symptoms | 256 | 47212.38 | 542.2307 | 477.837 | 612.8833 | 2020 | IMASIS | secondary care |
| POTS symptoms | 890 | 125531.8 | 708.9838 | 663.164 | 757.1354 | 2021 | IMASIS | secondary care |
| POTS symptoms | 1002 | 71156.94 | 1408.155 | 1322.304 | 1498.117 | 2022 | IMASIS | secondary care |
| POTS symptoms | 735 | 123621.1 | 594.5588 | 552.3468 | 639.1412 | 2020 | AUSOM | secondary care |
| POTS symptoms | 2478 | 338726.3 | 731.5642 | 703.041 | 760.9476 | 2021 | AUSOM | secondary care |
| POTS symptoms | 2464 | 247061.4 | 997.323 | 958.3288 | 1037.497 | 2022 | AUSOM | secondary care |
| POTS symptoms | 313 | 19933.37 | 1570.231 | 1401.074 | 1754.183 | 2023 | AUSOM | secondary care |
| POTS symptoms | 207 | 629028.6 | 32.90789 | 28.57737 | 37.70924 | 2020 | CHUM | secondary care |
| POTS symptoms | 806 | 1890291 | 42.63893 | 39.7457 | 45.68709 | 2021 | CHUM | secondary care |
| POTS symptoms | 793 | 1896857 | 41.806 | 38.94653 | 44.81987 | 2022 | CHUM | secondary care |
| POTS diagnosis | 111 | 49018.86 | 226.4434 | 186.282 | 272.6963 | 2020 | IMASIS | secondary care |
| POTS diagnosis | 296 | 130550.7 | 226.7319 | 201.6355 | 254.0886 | 2021 | IMASIS | secondary care |
| POTS diagnosis | 273 | 74530.73 | 366.2919 | 324.1259 | 412.4201 | 2022 | IMASIS | secondary care |
| POTS diagnosis | 58 | 130862.9 | 44.32119 | 33.65494 | 57.29541 | 2020 | AUSOM | secondary care |
| POTS diagnosis | 181 | 359642.8 | 50.32771 | 43.26262 | 58.21759 | 2021 | AUSOM | secondary care |
| POTS diagnosis | 192 | 264959.8 | 72.46382 | 62.57596 | 83.47044 | 2022 | AUSOM | secondary care |
| POTS diagnosis | 27 | 21906.03 | 123.2537 | 81.22499 | 179.3277 | 2023 | AUSOM | secondary care |
| POTS diagnosis | 81 | 630233.3 | 12.85238 | 10.20665 | 15.97434 | 2020 | CHUM | secondary care |
| POTS diagnosis | 359 | 1894133 | 18.95327 | 17.04316 | 21.01889 | 2021 | CHUM | secondary care |
| POTS diagnosis | 351 | 1901081 | 18.46318 | 16.58195 | 20.49939 | 2022 | CHUM | secondary care |
| ME/CFS symptoms | 316 | 47347.06 | 667.4121 | 595.846 | 745.2067 | 2020 | IMASIS | secondary care |
| ME/CFS symptoms | 1087 | 125760.3 | 864.3427 | 813.717 | 917.2934 | 2021 | IMASIS | secondary care |
| ME/CFS symptoms | 1142 | 71065.95 | 1606.958 | 1515.097 | 1702.932 | 2022 | IMASIS | secondary care |
| ME/CFS symptoms | 269 | 128612.5 | 209.1554 | 184.9055 | 235.7018 | 2020 | AUSOM | secondary care |
| ME/CFS symptoms | 592 | 353077 | 167.6688 | 154.4327 | 181.7361 | 2021 | AUSOM | secondary care |
| ME/CFS symptoms | 757 | 259160.2 | 292.0973 | 271.6573 | 313.6678 | 2022 | AUSOM | secondary care |
| ME/CFS symptoms | 86 | 21236.32 | 404.9666 | 323.921 | 500.1305 | 2023 | AUSOM | secondary care |
| ME/CFS symptoms | 124 | 629800.1 | 19.68879 | 16.37611 | 23.4748 | 2020 | CHUM | secondary care |
| ME/CFS symptoms | 446 | 1892687 | 23.56438 | 21.4279 | 25.85621 | 2021 | CHUM | secondary care |
| ME/CFS symptoms | 287 | 1899600 | 15.10845 | 13.41089 | 16.96139 | 2022 | CHUM | secondary care |
| ME/CFS diagnosis | 5 | 49377.74 | 10.12602 | 3.287892 | 23.63076 | 2020 | IMASIS | secondary care |
| ME/CFS diagnosis | 42 | 131634.8 | 31.90645 | 22.99536 | 43.12825 | 2021 | IMASIS | secondary care |
| ME/CFS diagnosis | 60 | 75307.29 | 79.67356 | 60.79932 | 102.5557 | 2022 | IMASIS | secondary care |
| ME/CFS diagnosis | 14 | 131223.6 | 10.66881 | 5.832739 | 17.90046 | 2020 | AUSOM | secondary care |
| ME/CFS diagnosis | 33 | 360685.8 | 9.149238 | 6.297914 | 12.84893 | 2021 | AUSOM | secondary care |
| ME/CFS diagnosis | 64 | 265892.1 | 24.06991 | 18.53676 | 30.73674 | 2022 | AUSOM | secondary care |
| ME/CFS diagnosis | 5 | 22014.28 | 22.71254 | 7.374698 | 53.00348 | 2023 | AUSOM | secondary care |
| ME/CFS diagnosis | 12 | 631120.4 | 1.90138 | 0.982471 | 3.321329 | 2020 | CHUM | secondary care |
| ME/CFS diagnosis | 54 | 1896924 | 2.846714 | 2.138539 | 3.714344 | 2021 | CHUM | secondary care |
| ME/CFS diagnosis | 51 | 1904127 | 2.678393 | 1.994239 | 3.521592 | 2022 | CHUM | secondary care |
| MIS | 5 | 631322 | 0.791989 | 0.257157 | 1.848238 | 2020 | CHUM | secondary care |
| MIS | 23 | 1897547 | 1.212091 | 0.768362 | 1.818732 | 2021 | CHUM | secondary care |
| MIS | 30 | 1904762 | 1.575 | 1.062646 | 2.24841 | 2022 | CHUM | secondary care |
| DM | 31 | 49263.5 | 62.92691 | 42.75578 | 89.31972 | 2020 | IMASIS | secondary care |
| DM | 128 | 131239.2 | 97.5318 | 81.36845 | 115.9656 | 2021 | IMASIS | secondary care |
| DM | 61 | 74937.02 | 81.40169 | 62.26584 | 104.5639 | 2022 | IMASIS | secondary care |
| DM | 18 | 130949.8 | 13.74573 | 8.146591 | 21.72418 | 2020 | AUSOM | secondary care |
| DM | 66 | 359856.8 | 18.34063 | 14.18464 | 23.33378 | 2021 | AUSOM | secondary care |
| DM | 53 | 265065.5 | 19.99506 | 14.97767 | 26.15403 | 2022 | AUSOM | secondary care |
| DM | 61 | 630314.2 | 9.677713 | 7.402684 | 12.43143 | 2020 | CHUM | secondary care |
| DM | 184 | 1894444 | 9.712613 | 8.359878 | 11.2219 | 2021 | CHUM | secondary care |
| DM | 121 | 1901610 | 6.363029 | 5.279873 | 7.603007 | 2022 | CHUM | secondary care |

IBD: inflammatory bowel disease; ME/CFS: myalgic encephalomyelitis / chronic fatigues syndrome; NA: results suppressed because less than 5 outcomes; POTS: postural orthostatic tachycardia syndrome; RA: rheumatoid arthritis; SLE: systemic lupus erythematosus; DM: diabetes mellitus

### Supplementary File 6. Numeric values of incidence rates with 95% confidence intervals corresponding to Figure 4

###### Supplementary Table 6. Numeric values of incidence rates with 95% confidence intervals corresponding to Figure 4

| **outcome** | **# events** | **Person years** | **Incidence**  **(per 100000 pys)** | **95% CI**  **lower bound** | **95% CI**  **upper bound** | **Age [years]** | **database** | **Care sector** |
| --- | --- | --- | --- | --- | --- | --- | --- | --- |
| POTS symptoms | 5396 | 378126.7 | 1427.035 | 1389.211 | 1465.628 | 0-6 | P+ | both |
| POTS symptoms | 6265 | 374755.8 | 1671.755 | 1630.612 | 1713.674 | 7-11 | P+ | both |
| POTS symptoms | 24267 | 556144.8 | 4363.432 | 4308.703 | 4418.683 | 12-18 | P+ | both |
| POTS symptoms | 117322 | 1791431 | 6549.068 | 6511.646 | 6586.651 | 19-40 | P+ | both |
| POTS symptoms | 180642 | 2221492 | 8131.563 | 8094.107 | 8169.149 | 41-64 | P+ | both |
| POTS symptoms | 198171 | 1623475 | 12206.6 | 12152.91 | 12260.46 | >64 | P+ | both |
| POTS symptoms | 134 | 44566.34 | 300.6753 | 251.9239 | 356.1086 | 0-6 | CORIVA | both |
| POTS symptoms | 343 | 47538.93 | 721.514 | 647.169 | 802.0577 | 7-11 | CORIVA | both |
| POTS symptoms | 1194 | 71285.58 | 1674.953 | 1581.283 | 1772.723 | 12-18 | CORIVA | both |
| POTS symptoms | 4797 | 282288.2 | 1699.327 | 1651.575 | 1748.11 | 19-40 | CORIVA | both |
| POTS symptoms | 5723 | 304151.3 | 1881.629 | 1833.192 | 1931.022 | 41-64 | CORIVA | both |
| POTS symptoms | 3699 | 163791.2 | 2258.363 | 2186.165 | 2332.337 | >64 | CORIVA | both |
| POTS symptoms | 6610 | 515102.6 | 1283.24 | 1252.488 | 1314.555 | 0-6 | NLHR@UIO | both |
| POTS symptoms | 5265 | 408059.4 | 1290.253 | 1255.634 | 1325.585 | 7-11 | NLHR@UIO | both |
| POTS symptoms | 18914 | 534299.1 | 3539.965 | 3489.693 | 3590.78 | 12-18 | NLHR@UIO | both |
| POTS symptoms | 81355 | 1790784 | 4542.983 | 4511.819 | 4574.31 | 19-40 | NLHR@UIO | both |
| POTS symptoms | 82667 | 1852959 | 4461.351 | 4430.99 | 4491.869 | 41-64 | NLHR@UIO | both |
| POTS symptoms | 54968 | 1023099 | 5372.694 | 5327.873 | 5417.799 | >64 | NLHR@UIO | both |
| POTS diagnosis | 1483 | 385710.4 | 384.4854 | 365.1636 | 404.5641 | 0-6 | P+ | both |
| POTS diagnosis | 903 | 389456.2 | 231.8618 | 216.9836 | 247.4915 | 7-11 | P+ | both |
| POTS diagnosis | 3747 | 614313.6 | 609.9491 | 590.5738 | 629.7981 | 12-18 | P+ | both |
| POTS diagnosis | 22139 | 2135672 | 1036.629 | 1023.018 | 1050.375 | 19-40 | P+ | both |
| POTS diagnosis | 37042 | 2888134 | 1282.558 | 1269.53 | 1295.687 | 41-64 | P+ | both |
| POTS diagnosis | 66704 | 2523742 | 2643.059 | 2623.039 | 2663.194 | >64 | P+ | both |
| POTS diagnosis | 15 | 44858.83 | 33.43824 | 18.71513 | 55.15129 | 0-6 | CORIVA | both |
| POTS diagnosis | 22 | 48404.35 | 45.45046 | 28.48356 | 68.81255 | 7-11 | CORIVA | both |
| POTS diagnosis | 170 | 74510.66 | 228.1553 | 195.1467 | 265.1479 | 12-18 | CORIVA | both |
| POTS diagnosis | 919 | 300442.6 | 305.8821 | 286.4229 | 326.3153 | 19-40 | CORIVA | both |
| POTS diagnosis | 1107 | 327374.5 | 338.1448 | 318.5163 | 358.6664 | 41-64 | CORIVA | both |
| POTS diagnosis | 646 | 180631.6 | 357.6341 | 330.5836 | 386.3081 | >64 | CORIVA | both |
| POTS diagnosis | 22 | 537729 | 4.09128 | 2.563983 | 6.194247 | 0-6 | NLHR@UIO | both |
| POTS diagnosis | 22 | 430139.4 | 5.114621 | 3.205305 | 7.743597 | 7-11 | NLHR@UIO | both |
| POTS diagnosis | 146 | 605709.8 | 24.10395 | 20.35274 | 28.34622 | 12-18 | NLHR@UIO | both |
| POTS diagnosis | 1298 | 2179011 | 59.56831 | 56.37139 | 62.8993 | 19-40 | NLHR@UIO | both |
| POTS diagnosis | 2486 | 2253171 | 110.3334 | 106.0384 | 114.7577 | 41-64 | NLHR@UIO | both |
| POTS diagnosis | 2491 | 1289186 | 193.2227 | 185.7085 | 200.9628 | >64 | NLHR@UIO | both |
| ME/CFS symptoms | 4158 | 381978.9 | 1088.542 | 1055.704 | 1122.141 | 0-6 | P+ | both |
| ME/CFS symptoms | 4882 | 378831.6 | 1288.699 | 1252.8 | 1325.366 | 7-11 | P+ | both |
| ME/CFS symptoms | 18969 | 572033.8 | 3316.063 | 3269.039 | 3363.594 | 12-18 | P+ | both |
| ME/CFS symptoms | 93681 | 1874670 | 4997.2 | 4965.25 | 5029.303 | 19-40 | P+ | both |
| ME/CFS symptoms | 143517 | 2397578 | 5985.915 | 5954.985 | 6016.965 | 41-64 | P+ | both |
| ME/CFS symptoms | 184858 | 1867732 | 9897.459 | 9852.391 | 9942.682 | >64 | P+ | both |
| ME/CFS symptoms | 159 | 44439.41 | 357.7905 | 304.3383 | 417.9285 | 0-6 | CORIVA | both |
| ME/CFS symptoms | 235 | 47713.25 | 492.5257 | 431.5623 | 559.6878 | 7-11 | CORIVA | both |
| ME/CFS symptoms | 804 | 72603.07 | 1107.391 | 1032.158 | 1186.658 | 12-18 | CORIVA | both |
| ME/CFS symptoms | 3525 | 289219.5 | 1218.798 | 1178.891 | 1259.71 | 19-40 | CORIVA | both |
| ME/CFS symptoms | 3835 | 315338.3 | 1216.154 | 1177.965 | 1255.266 | 41-64 | CORIVA | both |
| ME/CFS symptoms | 2021 | 174244.8 | 1159.862 | 1109.84 | 1211.557 | >64 | CORIVA | both |
| ME/CFS symptoms | 5696 | 517044.1 | 1101.647 | 1073.221 | 1130.635 | 0-6 | NLHR@UIO | both |
| ME/CFS symptoms | 3735 | 413673.5 | 902.8859 | 874.1597 | 932.3156 | 7-11 | NLHR@UIO | both |
| ME/CFS symptoms | 13745 | 552781.3 | 2486.517 | 2445.12 | 2528.439 | 12-18 | NLHR@UIO | both |
| ME/CFS symptoms | 60327 | 1900513 | 3174.248 | 3148.968 | 3199.681 | 19-40 | NLHR@UIO | both |
| ME/CFS symptoms | 52387 | 2010273 | 2605.964 | 2583.696 | 2628.377 | 41-64 | NLHR@UIO | both |
| ME/CFS symptoms | 25652 | 1186657 | 2161.702 | 2135.329 | 2188.32 | >64 | NLHR@UIO | both |
| ME/CFS diagnosis | 81 | 390314.7 | 20.75249 | 16.48047 | 25.79345 | 0-6 | P+ | both |
| ME/CFS diagnosis | 165 | 392204.8 | 42.06986 | 35.89551 | 49.00138 | 7-11 | P+ | both |
| ME/CFS diagnosis | 955 | 620694.4 | 153.8599 | 144.2552 | 163.9361 | 12-18 | P+ | both |
| ME/CFS diagnosis | 8240 | 2174013 | 379.0226 | 370.8826 | 387.2963 | 19-40 | P+ | both |
| ME/CFS diagnosis | 15711 | 2947148 | 533.0916 | 524.7879 | 541.4937 | 41-64 | P+ | both |
| ME/CFS diagnosis | 18527 | 2697538 | 686.8115 | 676.9569 | 696.7736 | >64 | P+ | both |
| ME/CFS diagnosis | 9 | 44911.45 | 20.03943 | 9.163305 | 38.04109 | 0-6 | CORIVA | both |
| ME/CFS diagnosis | 17 | 48448.62 | 35.08872 | 20.44047 | 56.18044 | 7-11 | CORIVA | both |
| ME/CFS diagnosis | 53 | 74884.94 | 70.77524 | 53.01551 | 92.57576 | 12-18 | CORIVA | both |
| ME/CFS diagnosis | 182 | 303891.4 | 59.88982 | 51.50463 | 69.25105 | 19-40 | CORIVA | both |
| ME/CFS diagnosis | 293 | 332205.5 | 88.19841 | 78.38755 | 98.8976 | 41-64 | CORIVA | both |
| ME/CFS diagnosis | 108 | 184415.2 | 58.5635 | 48.04077 | 70.70605 | >64 | CORIVA | both |
| ME/CFS diagnosis | 5 | 537785.3 | 0.929739 | 0.301884 | 2.169701 | 0-6 | NLHR@UIO | both |
| ME/CFS diagnosis | 13 | 430218.3 | 3.021722 | 1.60894 | 5.167236 | 7-11 | NLHR@UIO | both |
| ME/CFS diagnosis | 51 | 606062.9 | 8.414968 | 6.265496 | 11.06413 | 12-18 | NLHR@UIO | both |
| ME/CFS diagnosis | 667 | 2184270 | 30.53651 | 28.26275 | 32.9445 | 19-40 | NLHR@UIO | both |
| ME/CFS diagnosis | 662 | 2265815 | 29.21686 | 27.03332 | 31.52981 | 41-64 | NLHR@UIO | both |
| ME/CFS diagnosis | 97 | 1304978 | 7.433075 | 6.027723 | 9.067722 | >64 | NLHR@UIO | both |
| MIS | 39 | 2198367 | 1.774044 | 1.261519 | 2.425176 | 19-40 | P+ | both |
| MIS | 91 | 3004942 | 3.028344 | 2.438234 | 3.718136 | 41-64 | P+ | both |
| MIS | 129 | 2776919 | 4.645436 | 3.878429 | 5.519736 | >64 | P+ | both |
| MIS | 50 | 390564.6 | 12.80198 | 9.501876 | 16.87781 | 0-6 | P+ | both |
| MIS | 47 | 392777.2 | 11.96607 | 8.792219 | 15.91234 | 7-11 | P+ | both |
| MIS | 61 | 623120.7 | 9.789436 | 7.488143 | 12.57494 | 12-18 | P+ | both |
| MIS | 8 | 304339.4 | 2.628644 | 1.134862 | 5.179477 | 19-40 | CORIVA | both |
| MIS | 38 | 332907.6 | 11.41458 | 8.077636 | 15.6674 | 41-64 | CORIVA | both |
| MIS | 107 | 184650.4 | 57.94735 | 47.48925 | 70.02344 | >64 | CORIVA | both |
| DM | 100 | 390360.7 | 25.61733 | 20.84328 | 31.15754 | 0-6 | P+ | both |
| DM | 196 | 391951.6 | 50.00617 | 43.25013 | 57.51832 | 7-11 | P+ | both |
| DM | 240 | 620390.3 | 38.68533 | 33.94546 | 43.90182 | 12-18 | P+ | both |
| DM | 809 | 2185476 | 37.01711 | 34.50992 | 39.6583 | 19-40 | P+ | both |
| DM | 2598 | 2981336 | 87.14214 | 83.82316 | 90.55885 | 41-64 | P+ | both |
| DM | 2752 | 2755078 | 99.88829 | 96.19081 | 103.6915 | >64 | P+ | both |
| DM | 14 | 44893.42 | 31.18497 | 17.04911 | 52.32308 | 0-6 | CORIVA | both |
| DM | 23 | 48353.79 | 47.56608 | 30.15281 | 71.37247 | 7-11 | CORIVA | both |
| DM | 22 | 74691.04 | 29.45467 | 18.45908 | 44.59472 | 12-18 | CORIVA | both |
| DM | 60 | 303036.1 | 19.79962 | 15.1092 | 25.48604 | 19-40 | CORIVA | both |
| DM | 106 | 330962.4 | 32.02781 | 26.22175 | 38.73671 | 41-64 | CORIVA | both |
| DM | 168 | 182980.3 | 91.81316 | 78.45432 | 106.7946 | >64 | CORIVA | both |
| DM | 104 | 537570.8 | 19.34629 | 15.80732 | 23.44127 | 0-6 | NLHR@UIO | both |
| DM | 186 | 429374.9 | 43.31879 | 37.31681 | 50.01138 | 7-11 | NLHR@UIO | both |
| DM | 328 | 603726.3 | 54.32925 | 48.60811 | 60.53869 | 12-18 | NLHR@UIO | both |
| DM | 787 | 2172252 | 36.22969 | 33.74238 | 38.85184 | 19-40 | NLHR@UIO | both |
| DM | 1332 | 2244251 | 59.35166 | 56.20673 | 62.62675 | 41-64 | NLHR@UIO | both |
| DM | 1342 | 1284265 | 104.4956 | 98.97894 | 110.2396 | >64 | NLHR@UIO | Both |
| POTS symptoms | 441 | 310059 | 142.231 | 129.2644 | 156.146 | 0-6 | CPRD GOLD | primary care |
| POTS symptoms | 854 | 304658.1 | 280.3142 | 261.8267 | 299.7626 | 7-11 | CPRD GOLD | primary care |
| POTS symptoms | 3142 | 377629.3 | 832.0328 | 803.1916 | 861.645 | 12-18 | CPRD GOLD | primary care |
| POTS symptoms | 11664 | 1240465 | 940.2929 | 923.3051 | 957.5147 | 19-40 | CPRD GOLD | primary care |
| POTS symptoms | 15729 | 1346029 | 1168.548 | 1150.357 | 1186.955 | 41-64 | CPRD GOLD | primary care |
| POTS symptoms | 13022 | 680069.2 | 1914.805 | 1882.057 | 1947.98 | >64 | CPRD GOLD | primary care |
| POTS symptoms | 2166 | 162558.9 | 1332.441 | 1276.912 | 1389.762 | 0-6 | IPCI | primary care |
| POTS symptoms | 2538 | 141802.9 | 1789.808 | 1720.846 | 1860.825 | 7-11 | IPCI | primary care |
| POTS symptoms | 9141 | 178587 | 5118.514 | 5014.117 | 5224.538 | 12-18 | IPCI | primary care |
| POTS symptoms | 24828 | 515151.4 | 4819.554 | 4759.789 | 4879.882 | 19-40 | IPCI | primary care |
| POTS symptoms | 28826 | 640784.3 | 4498.55 | 4446.766 | 4550.785 | 41-64 | IPCI | primary care |
| POTS symptoms | 23327 | 353303.2 | 6602.545 | 6518.084 | 6687.826 | >64 | IPCI | primary care |
| POTS symptoms | 2515 | 1115189 | 225.5223 | 216.7936 | 234.5123 | 0-6 | CPRD Aurum | primary care |
| POTS symptoms | 5160 | 1121829 | 459.9633 | 447.4979 | 472.6879 | 7-11 | CPRD Aurum | primary care |
| POTS symptoms | 19991 | 1432397 | 1395.632 | 1376.352 | 1415.115 | 12-18 | CPRD Aurum | primary care |
| POTS symptoms | 78713 | 4725118 | 1665.842 | 1654.225 | 1677.521 | 19-40 | CPRD Aurum | primary care |
| POTS symptoms | 87967 | 4300148 | 2045.674 | 2032.177 | 2059.237 | 41-64 | CPRD Aurum | primary care |
| POTS symptoms | 60407 | 1881564 | 3210.467 | 3184.915 | 3236.173 | >64 | CPRD Aurum | primary care |
| POTS diagnosis | 33 | 311483 | 10.59448 | 7.292751 | 14.87859 | 0-6 | CPRD GOLD | primary care |
| POTS diagnosis | 35 | 310747.3 | 11.26317 | 7.845211 | 15.66434 | 7-11 | CPRD GOLD | primary care |
| POTS diagnosis | 162 | 402427.4 | 40.25571 | 34.29537 | 46.95416 | 12-18 | CPRD GOLD | primary care |
| POTS diagnosis | 785 | 1454403 | 53.97404 | 50.26387 | 57.8856 | 19-40 | CPRD GOLD | primary care |
| POTS diagnosis | 1168 | 1750492 | 66.72408 | 62.95191 | 70.66323 | 41-64 | CPRD GOLD | primary care |
| POTS diagnosis | 1194 | 1061845 | 112.4458 | 106.1574 | 119.0094 | >64 | CPRD GOLD | primary care |
| POTS diagnosis | 7 | 168606.1 | 4.151689 | 1.669194 | 8.554065 | 0-6 | IPCI | primary care |
| POTS diagnosis | 11 | 156430.6 | 7.031874 | 3.510286 | 12.58196 | 7-11 | IPCI | primary care |
| POTS diagnosis | 59 | 225575.3 | 26.15535 | 19.91066 | 33.73849 | 12-18 | IPCI | primary care |
| POTS diagnosis | 347 | 714572.6 | 48.5605 | 43.58495 | 53.94839 | 19-40 | IPCI | primary care |
| POTS diagnosis | 799 | 912122.2 | 87.59792 | 81.6285 | 93.88843 | 41-64 | IPCI | primary care |
| POTS diagnosis | 977 | 562414.4 | 173.7153 | 162.9919 | 184.9589 | >64 | IPCI | primary care |
| POTS diagnosis | 233 | 1125427 | 20.70326 | 18.13006 | 23.53928 | 0-6 | CPRD Aurum | primary care |
| POTS diagnosis | 180 | 1155981 | 15.57119 | 13.37945 | 18.01956 | 7-11 | CPRD Aurum | primary care |
| POTS diagnosis | 1113 | 1551425 | 71.74049 | 67.58718 | 76.08223 | 12-18 | CPRD Aurum | primary care |
| POTS diagnosis | 6462 | 5516382 | 117.142 | 114.3031 | 120.0336 | 19-40 | CPRD Aurum | primary care |
| POTS diagnosis | 8234 | 5786859 | 142.2879 | 139.231 | 145.395 | 41-64 | CPRD Aurum | primary care |
| POTS diagnosis | 8198 | 3163522 | 259.1415 | 253.5619 | 264.8129 | >64 | CPRD Aurum | primary care |
| ME/CFS symptoms | 1147 | 303353.9 | 378.1062 | 356.5384 | 400.6376 | 0-6 | CPRD GOLD | primary care |
| ME/CFS symptoms | 552 | 296942.9 | 185.8943 | 170.7081 | 202.0692 | 7-11 | CPRD GOLD | primary care |
| ME/CFS symptoms | 1849 | 374949.3 | 493.1333 | 470.9097 | 516.1349 | 12-18 | CPRD GOLD | primary care |
| ME/CFS symptoms | 9277 | 1267052 | 732.1721 | 717.348 | 747.2255 | 19-40 | CPRD GOLD | primary care |
| ME/CFS symptoms | 9363 | 1447612 | 646.7894 | 633.754 | 660.0255 | 41-64 | CPRD GOLD | primary care |
| ME/CFS symptoms | 6801 | 863119.1 | 787.9561 | 769.3393 | 806.9096 | >64 | CPRD GOLD | primary care |
| ME/CFS symptoms | 1658 | 164356.2 | 1008.785 | 960.8061 | 1058.539 | 0-6 | IPCI | primary care |
| ME/CFS symptoms | 1585 | 145490.6 | 1089.418 | 1036.439 | 1144.402 | 7-11 | IPCI | primary care |
| ME/CFS symptoms | 6271 | 191915.2 | 3267.589 | 3187.21 | 3349.483 | 12-18 | IPCI | primary care |
| ME/CFS symptoms | 18247 | 565656 | 3225.812 | 3179.175 | 3272.962 | 19-40 | IPCI | primary care |
| ME/CFS symptoms | 17540 | 742103.2 | 2363.553 | 2328.702 | 2398.794 | 41-64 | IPCI | primary care |
| ME/CFS symptoms | 12566 | 464459.4 | 2705.511 | 2658.411 | 2753.236 | >64 | IPCI | primary care |
| ME/CFS symptoms | 5315 | 1089585 | 487.8006 | 474.7736 | 501.0944 | 0-6 | CPRD Aurum | primary care |
| ME/CFS symptoms | 3142 | 1089439 | 288.4054 | 278.4083 | 298.6698 | 7-11 | CPRD Aurum | primary care |
| ME/CFS symptoms | 12844 | 1423935 | 902.0075 | 886.4747 | 917.7442 | 12-18 | CPRD Aurum | primary care |
| ME/CFS symptoms | 64552 | 4802570 | 1344.114 | 1333.765 | 1354.523 | 19-40 | CPRD Aurum | primary care |
| ME/CFS symptoms | 54246 | 4648968 | 1166.84 | 1157.041 | 1176.701 | 41-64 | CPRD Aurum | primary care |
| ME/CFS symptoms | 35311 | 2480868 | 1423.332 | 1408.525 | 1438.257 | >64 | CPRD Aurum | primary care |
| ME/CFS diagnosis | 5 | 311744.5 | 1.603878 | 0.520775 | 3.742916 | 0-6 | CPRD GOLD | primary care |
| ME/CFS diagnosis | 14 | 311051.4 | 4.500863 | 2.460664 | 7.551684 | 7-11 | CPRD GOLD | primary care |
| ME/CFS diagnosis | 60 | 402967.4 | 14.88954 | 11.36229 | 19.16579 | 12-18 | CPRD GOLD | primary care |
| ME/CFS diagnosis | 304 | 1458134 | 20.84857 | 18.57056 | 23.32889 | 19-40 | CPRD GOLD | primary care |
| ME/CFS diagnosis | 497 | 1756646 | 28.29256 | 25.85953 | 30.89284 | 41-64 | CPRD GOLD | primary care |
| ME/CFS diagnosis | 113 | 1074593 | 10.51561 | 8.666349 | 12.64266 | >64 | CPRD GOLD | primary care |
| ME/CFS diagnosis | 31 | 225710.2 | 13.73442 | 9.331875 | 19.49492 | 12-18 | IPCI | primary care |
| ME/CFS diagnosis | 150 | 715438 | 20.96618 | 17.74523 | 24.60271 | 19-40 | IPCI | primary care |
| ME/CFS diagnosis | 179 | 916631.3 | 19.52803 | 16.77196 | 22.60774 | 41-64 | IPCI | primary care |
| ME/CFS diagnosis | 65 | 570477.6 | 11.39396 | 8.793618 | 14.52254 | >64 | IPCI | primary care |
| ME/CFS diagnosis | 21 | 1126786 | 1.863709 | 1.153665 | 2.848877 | 0-6 | CPRD Aurum | primary care |
| ME/CFS diagnosis | 63 | 1157180 | 5.44427 | 4.183527 | 6.965587 | 7-11 | CPRD Aurum | primary care |
| ME/CFS diagnosis | 344 | 1553651 | 22.1414 | 19.86317 | 24.60929 | 12-18 | CPRD Aurum | primary care |
| ME/CFS diagnosis | 1757 | 5536201 | 31.73656 | 30.26979 | 33.25604 | 19-40 | CPRD Aurum | primary care |
| ME/CFS diagnosis | 2104 | 5819536 | 36.15409 | 34.62559 | 37.73268 | 41-64 | CPRD Aurum | primary care |
| ME/CFS diagnosis | 383 | 3228008 | 11.8649 | 10.70625 | 13.11476 | >64 | CPRD Aurum | primary care |
| MIS | 8 | 1126854 | 0.709941 | 0.306502 | 1.398867 | 0-6 | CPRD Aurum | primary care |
| MIS | 43 | 1157449 | 3.715066 | 2.688611 | 5.004169 | 7-11 | CPRD Aurum | primary care |
| MIS | 21 | 1555617 | 1.349947 | 0.835638 | 2.063537 | 12-18 | CPRD Aurum | primary care |
| DM | 106 | 311493.6 | 34.02959 | 27.86064 | 41.1578 | 0-6 | CPRD GOLD | primary care |
| DM | 173 | 310352 | 55.74316 | 47.74595 | 64.69668 | 7-11 | CPRD GOLD | primary care |
| DM | 188 | 401385 | 46.83782 | 40.38159 | 54.03275 | 12-18 | CPRD GOLD | primary care |
| DM | 328 | 1454435 | 22.55172 | 20.17691 | 25.12921 | 19-40 | CPRD GOLD | primary care |
| DM | 311 | 1756606 | 17.7046 | 15.79138 | 19.78572 | 41-64 | CPRD GOLD | primary care |
| DM | 132 | 1075077 | 12.27819 | 10.27306 | 14.56036 | >64 | CPRD GOLD | primary care |
| DM | 24 | 168599.8 | 14.2349 | 9.120567 | 21.1804 | 0-6 | IPCI | primary care |
| DM | 40 | 156317.2 | 25.589 | 18.28116 | 34.84496 | 7-11 | IPCI | primary care |
| DM | 45 | 225447.6 | 19.96029 | 14.55917 | 26.70844 | 12-18 | IPCI | primary care |
| DM | 116 | 715430.2 | 16.21402 | 13.39796 | 19.44717 | 19-40 | IPCI | primary care |
| DM | 107 | 916280.1 | 11.67765 | 9.570117 | 14.11125 | 41-64 | IPCI | primary care |
| DM | 94 | 569879.6 | 16.49471 | 13.32941 | 20.18535 | >64 | IPCI | primary care |
| DM | 267 | 1126274 | 23.70648 | 20.94794 | 26.72727 | 0-6 | CPRD Aurum | primary care |
| DM | 460 | 1155465 | 39.81081 | 36.2554 | 43.62063 | 7-11 | CPRD Aurum | primary care |
| DM | 585 | 1550058 | 37.74052 | 34.74379 | 40.92658 | 12-18 | CPRD Aurum | primary care |
| DM | 1413 | 5526765 | 25.56649 | 24.25066 | 26.93517 | 19-40 | CPRD Aurum | primary care |
| DM | 1140 | 5820907 | 19.58458 | 18.46407 | 20.7553 | 41-64 | CPRD Aurum | primary care |
| DM | 417 | 3230439 | 12.90846 | 11.69908 | 14.20891 | >64 | CPRD Aurum | primary care |
| POTS symptoms | 5 | 8171.083 | 61.1914 | 19.86868 | 142.8003 | 0-6 | IMASIS | secondary care |
| POTS symptoms | 19 | 5925.676 | 320.6385 | 193.0454 | 500.7168 | 7-11 | IMASIS | secondary care |
| POTS symptoms | 62 | 8623.379 | 718.9757 | 551.2346 | 921.6945 | 12-18 | IMASIS | secondary care |
| POTS symptoms | 384 | 56053.98 | 685.0539 | 618.2406 | 757.1192 | 19-40 | IMASIS | secondary care |
| POTS symptoms | 786 | 91782.53 | 856.3721 | 797.542 | 918.3935 | 41-64 | IMASIS | secondary care |
| POTS symptoms | 892 | 73344.44 | 1216.18 | 1137.668 | 1298.682 | >64 | IMASIS | secondary care |
| POTS symptoms | 145 | 37101.72 | 390.8174 | 329.7956 | 459.8568 | 0-6 | AUSOM | secondary care |
| POTS symptoms | 252 | 42359.67 | 594.9054 | 523.7156 | 673.0723 | 7-11 | AUSOM | secondary care |
| POTS symptoms | 755 | 29441.19 | 2564.435 | 2384.751 | 2754.07 | 12-18 | AUSOM | secondary care |
| POTS symptoms | 2282 | 188635.9 | 1209.738 | 1160.607 | 1260.413 | 19-40 | AUSOM | secondary care |
| POTS symptoms | 4316 | 352856.3 | 1223.161 | 1186.939 | 1260.208 | 41-64 | AUSOM | secondary care |
| POTS symptoms | 2127 | 164181.2 | 1295.52 | 1241.043 | 1351.773 | >64 | AUSOM | secondary care |
| POTS symptoms | 22 | 197929.7 | 11.11505 | 6.965746 | 16.82833 | 0-6 | CHUM | secondary care |
| POTS symptoms | 17 | 201384.3 | 8.441571 | 4.917526 | 13.51577 | 7-11 | CHUM | secondary care |
| POTS symptoms | 42 | 320820.3 | 13.09144 | 9.435161 | 17.69583 | 12-18 | CHUM | secondary care |
| POTS symptoms | 186 | 1300010 | 14.30758 | 12.32522 | 16.51805 | 19-40 | CHUM | secondary care |
| POTS symptoms | 416 | 1252786 | 33.20599 | 30.09132 | 36.5555 | 41-64 | CHUM | secondary care |
| POTS symptoms | 1123 | 1143247 | 98.229 | 92.56718 | 104.1465 | >64 | CHUM | secondary care |
| POTS diagnosis | 11 | 8849.415 | 124.302 | 62.05111 | 222.4106 | 12-18 | IMASIS | secondary care |
| POTS diagnosis | 113 | 57621.47 | 196.1074 | 161.6203 | 235.7752 | 19-40 | IMASIS | secondary care |
| POTS diagnosis | 222 | 94936.88 | 233.8396 | 204.0888 | 266.7076 | 41-64 | IMASIS | secondary care |
| POTS diagnosis | 333 | 78535.58 | 424.0116 | 379.6882 | 472.0882 | >64 | IMASIS | secondary care |
| POTS diagnosis | 20 | 37182.25 | 53.78911 | 32.85579 | 83.07292 | 0-6 | AUSOM | secondary care |
| POTS diagnosis | 18 | 42636.43 | 42.21742 | 25.02072 | 66.72172 | 7-11 | AUSOM | secondary care |
| POTS diagnosis | 55 | 30262.71 | 181.7418 | 136.9128 | 236.562 | 12-18 | AUSOM | secondary care |
| POTS diagnosis | 186 | 194086.4 | 95.83359 | 82.5555 | 110.6395 | 19-40 | AUSOM | secondary care |
| POTS diagnosis | 332 | 376457.8 | 88.19049 | 78.95815 | 98.20583 | 41-64 | AUSOM | secondary care |
| POTS diagnosis | 168 | 183261.8 | 91.67213 | 78.33381 | 106.6305 | >64 | AUSOM | secondary care |
| POTS diagnosis | 15 | 197979.3 | 7.576551 | 4.240538 | 12.49637 | 0-6 | CHUM | secondary care |
| POTS diagnosis | 8 | 201505.8 | 3.970109 | 1.714011 | 7.822698 | 7-11 | CHUM | secondary care |
| POTS diagnosis | 27 | 321133.7 | 8.407713 | 5.540736 | 12.23278 | 12-18 | CHUM | secondary care |
| POTS diagnosis | 93 | 1301210 | 7.147193 | 5.768707 | 8.755795 | 19-40 | CHUM | secondary care |
| POTS diagnosis | 214 | 1254615 | 17.05702 | 14.84812 | 19.5019 | 41-64 | CHUM | secondary care |
| POTS diagnosis | 434 | 1149003 | 37.77189 | 34.3014 | 41.49835 | >64 | CHUM | secondary care |
| ME/CFS symptoms | 5 | 8167.502 | 61.21823 | 19.87739 | 142.8629 | 0-6 | IMASIS | secondary care |
| ME/CFS symptoms | 28 | 8818.319 | 317.5208 | 210.9903 | 458.906 | 12-18 | IMASIS | secondary care |
| ME/CFS symptoms | 324 | 56737.76 | 571.0482 | 510.5544 | 636.7385 | 19-40 | IMASIS | secondary care |
| ME/CFS symptoms | 865 | 91184.74 | 948.6236 | 886.4516 | 1014.006 | 41-64 | IMASIS | secondary care |
| ME/CFS symptoms | 1319 | 73297.64 | 1799.512 | 1703.697 | 1899.313 | >64 | IMASIS | secondary care |
| ME/CFS symptoms | 22 | 37176.36 | 59.1774 | 37.08616 | 89.59529 | 0-6 | AUSOM | secondary care |
| ME/CFS symptoms | 21 | 42604.62 | 49.29042 | 30.51155 | 75.34565 | 7-11 | AUSOM | secondary care |
| ME/CFS symptoms | 169 | 30056.72 | 562.2703 | 480.6926 | 653.7254 | 12-18 | AUSOM | secondary care |
| ME/CFS symptoms | 766 | 192018.1 | 398.9208 | 371.1671 | 428.2001 | 19-40 | AUSOM | secondary care |
| ME/CFS symptoms | 1028 | 368048.5 | 279.311 | 262.4956 | 296.9211 | 41-64 | AUSOM | secondary care |
| ME/CFS symptoms | 510 | 178413.2 | 285.8532 | 261.5795 | 311.7734 | >64 | AUSOM | secondary care |
| ME/CFS symptoms | 90 | 196947.5 | 45.69745 | 36.74615 | 56.16993 | 0-6 | CHUM | secondary care |
| ME/CFS symptoms | 27 | 200548.3 | 13.46309 | 8.872261 | 19.58809 | 7-11 | CHUM | secondary care |
| ME/CFS symptoms | 34 | 320391 | 10.61203 | 7.349141 | 14.82925 | 12-18 | CHUM | secondary care |
| ME/CFS symptoms | 108 | 1300692 | 8.303271 | 6.811334 | 10.02487 | 19-40 | CHUM | secondary care |
| ME/CFS symptoms | 212 | 1254009 | 16.90577 | 14.70652 | 19.34115 | 41-64 | CHUM | secondary care |
| ME/CFS symptoms | 386 | 1149499 | 33.57986 | 30.31309 | 37.10272 | >64 | CHUM | secondary care |
| ME/CFS diagnosis | 10 | 57872.82 | 17.27927 | 8.286081 | 31.77719 | 19-40 | IMASIS | secondary care |
| ME/CFS diagnosis | 80 | 95557.26 | 83.71944 | 66.38431 | 104.1961 | 41-64 | IMASIS | secondary care |
| ME/CFS diagnosis | 16 | 79856.63 | 20.03591 | 11.45225 | 32.53706 | >64 | IMASIS | secondary care |
| ME/CFS diagnosis | 17 | 37197.04 | 45.70256 | 26.62343 | 73.17423 | 0-6 | AUSOM | secondary care |
| ME/CFS diagnosis | 5 | 42645.44 | 11.72458 | 3.80694 | 27.36127 | 7-11 | AUSOM | secondary care |
| ME/CFS diagnosis | 9 | 30327.71 | 29.67583 | 13.56968 | 56.33397 | 12-18 | AUSOM | secondary care |
| ME/CFS diagnosis | 45 | 194321 | 23.15756 | 16.89128 | 30.98664 | 19-40 | AUSOM | secondary care |
| ME/CFS diagnosis | 91 | 377587.9 | 24.10035 | 19.4041 | 29.58989 | 41-64 | AUSOM | secondary care |
| ME/CFS diagnosis | 42 | 184343.5 | 22.78355 | 16.42038 | 30.79674 | >64 | AUSOM | secondary care |
| ME/CFS diagnosis | 16 | 198067 | 8.078074 | 4.617317 | 13.11829 | 0-6 | CHUM | secondary care |
| ME/CFS diagnosis | 10 | 201631.1 | 4.959551 | 2.378298 | 9.120791 | 7-11 | CHUM | secondary care |
| ME/CFS diagnosis | 10 | 321186.9 | 3.113452 | 1.493021 | 5.72575 | 12-18 | CHUM | secondary care |
| ME/CFS diagnosis | 19 | 1301877 | 1.459432 | 0.878673 | 2.279083 | 19-40 | CHUM | secondary care |
| ME/CFS diagnosis | 35 | 1255999 | 2.786625 | 1.940987 | 3.875521 | 41-64 | CHUM | secondary care |
| ME/CFS diagnosis | 27 | 1153410 | 2.340884 | 1.542657 | 3.405863 | >64 | CHUM | secondary care |
| MIS | 7 | 1153730 | 0.606728 | 0.243936 | 1.250091 | >64 | CHUM | secondary care |
| MIS | 15 | 198132.7 | 7.570683 | 4.237254 | 12.48669 | 0-6 | CHUM | secondary care |
| MIS | 20 | 201740.1 | 9.913746 | 6.055574 | 15.31098 | 7-11 | CHUM | secondary care |
| MIS | 9 | 321339.6 | 2.800775 | 1.280693 | 5.316744 | 12-18 | CHUM | secondary care |
| DM | 8 | 8833.632 | 90.56298 | 39.09867 | 178.4452 | 12-18 | IMASIS | secondary care |
| DM | 72 | 57560.75 | 125.0852 | 97.8715 | 157.5242 | 19-40 | IMASIS | secondary care |
| DM | 101 | 95206.83 | 106.0848 | 86.40784 | 128.9027 | 41-64 | IMASIS | secondary care |
| DM | 35 | 79701.38 | 43.91392 | 30.58766 | 61.07363 | >64 | IMASIS | secondary care |
| DM | 12 | 37205.26 | 32.2535 | 16.66585 | 56.34037 | 0-6 | AUSOM | secondary care |
| DM | 27 | 42585.95 | 63.40118 | 41.78178 | 92.2454 | 7-11 | AUSOM | secondary care |
| DM | 43 | 30147 | 142.6344 | 103.2252 | 192.1276 | 12-18 | AUSOM | secondary care |
| DM | 69 | 193785 | 35.60648 | 27.70396 | 45.06228 | 19-40 | AUSOM | secondary care |
| DM | 76 | 376811.8 | 20.16922 | 15.89105 | 25.24479 | 41-64 | AUSOM | secondary care |
| DM | 58 | 183776.5 | 31.56007 | 23.96488 | 40.79871 | >64 | AUSOM | secondary care |
| DM | 5 | 198124.5 | 2.523666 | 0.819427 | 5.889395 | 0-6 | CHUM | secondary care |
| DM | 11 | 201674.6 | 5.454331 | 2.722783 | 9.759305 | 7-11 | CHUM | secondary care |
| DM | 17 | 321162.7 | 5.293267 | 3.083523 | 8.475033 | 12-18 | CHUM | secondary care |
| DM | 63 | 1301201 | 4.841681 | 3.72048 | 6.194613 | 19-40 | CHUM | secondary care |
| DM | 106 | 1254468 | 8.449798 | 6.918002 | 10.21979 | 41-64 | CHUM | secondary care |
| DM | 164 | 1149738 | 14.26412 | 12.16455 | 16.62202 | >64 | CHUM | secondary care |

### Supplementary File 7. Numeric values of incidence rates with 95% confidence intervals corresponding to Figure 5

###### Supplementary Table 7. Numeric values of incidence rates with 95% confidence intervals corresponding to Figure 5

| **outcome** | **# events** | **Person years** | **Incidence**  **(per 100000 pys)** | **95% CI**  **lower bound** | **95% CI**  **upper bound** | **Sex** | **database** | **Care sector** |
| --- | --- | --- | --- | --- | --- | --- | --- | --- |
| POTS symptoms | 223442 | 3539527 | 6312.765 | 6286.616 | 6338.995 | Male | P+ | both |
| POTS symptoms | 308621 | 3405897 | 9061.371 | 9029.43 | 9093.398 | Female | P+ | both |
| POTS symptoms | 5199 | 451223.2 | 1152.201 | 1121.092 | 1183.955 | Male | CORIVA | both |
| POTS symptoms | 10691 | 462398.4 | 2312.075 | 2268.454 | 2356.325 | Female | CORIVA | both |
| POTS symptoms | 94445 | 3273997 | 2884.7 | 2866.332 | 2903.157 | Male | NLHR@UIO | both |
| POTS symptoms | 155334 | 2850306 | 5449.731 | 5422.663 | 5476.901 | Female | NLHR@UIO | both |
| POTS diagnosis | 56906 | 4306181 | 1321.496 | 1310.66 | 1332.399 | Male | P+ | both |
| POTS diagnosis | 75112 | 4630848 | 1621.992 | 1610.413 | 1633.634 | Female | P+ | both |
| POTS diagnosis | 953 | 469806.7 | 202.8494 | 190.1733 | 216.1482 | Male | CORIVA | both |
| POTS diagnosis | 1926 | 506415.7 | 380.3199 | 363.5226 | 397.6932 | Female | CORIVA | both |
| POTS diagnosis | 2574 | 3689240 | 69.77048 | 67.10089 | 72.51904 | Male | NLHR@UIO | both |
| POTS diagnosis | 3891 | 3605707 | 107.9123 | 104.5479 | 111.3573 | Female | NLHR@UIO | both |
| ME/CFS symptoms | 185549 | 3774773 | 4915.501 | 4893.16 | 4937.918 | Male | P+ | both |
| ME/CFS symptoms | 264516 | 3698052 | 7152.847 | 7125.615 | 7180.159 | Female | P+ | both |
| ME/CFS symptoms | 3694 | 459176.1 | 804.4845 | 778.7485 | 830.8542 | Male | CORIVA | both |
| ME/CFS symptoms | 6885 | 484382.3 | 1421.398 | 1388.019 | 1455.377 | Female | CORIVA | both |
| ME/CFS symptoms | 57733 | 3446724 | 1675.011 | 1661.375 | 1688.731 | Male | NLHR@UIO | both |
| ME/CFS symptoms | 103809 | 3134219 | 3312.117 | 3291.999 | 3332.328 | Female | NLHR@UIO | both |
| ME/CFS diagnosis | 15405 | 4436997 | 347.1943 | 341.733 | 352.721 | Male | P+ | both |
| ME/CFS diagnosis | 28274 | 4784916 | 590.8985 | 584.0308 | 597.8269 | Female | P+ | both |
| ME/CFS diagnosis | 217 | 473955.3 | 45.78491 | 39.89541 | 52.29896 | Male | CORIVA | both |
| ME/CFS diagnosis | 445 | 514801.8 | 86.44103 | 78.59523 | 94.85802 | Female | CORIVA | both |
| ME/CFS diagnosis | 433 | 3703134 | 11.6928 | 10.61725 | 12.84777 | Male | NLHR@UIO | both |
| ME/CFS diagnosis | 1062 | 3625996 | 29.2885 | 27.55327 | 31.10439 | Female | NLHR@UIO | both |
| MIS | 192 | 4493758 | 4.272593 | 3.689587 | 4.921563 | Male | P+ | both |
| MIS | 225 | 4892933 | 4.598469 | 4.017199 | 5.240214 | Female | P+ | both |
| MIS | 61 | 474499.5 | 12.85565 | 9.833554 | 16.51362 | Male | CORIVA | both |
| MIS | 100 | 515815.2 | 19.38679 | 15.77386 | 23.57953 | Female | CORIVA | both |
| MIS | 35 | 3703681 | 0.945006 | 0.658231 | 1.314274 | Male | NLHR@UIO | both |
| MIS | 16 | 3627265 | 0.441104 | 0.252129 | 0.716325 | Female | NLHR@UIO | both |
| DM | 3524 | 4461023 | 78.99534 | 76.40849 | 81.64743 | Male | P+ | both |
| DM | 3171 | 4863569 | 65.19903 | 62.94927 | 67.50865 | Female | P+ | both |
| DM | 211 | 471652 | 44.73638 | 38.90337 | 51.19719 | Male | CORIVA | both |
| DM | 182 | 513265.1 | 35.45926 | 30.4946 | 41.00181 | Female | CORIVA | both |
| DM | 2456 | 3669532 | 66.92951 | 64.30843 | 69.63 | Male | NLHR@UIO | both |
| DM | 1623 | 3601907 | 45.05947 | 42.89371 | 47.30625 | Female | NLHR@UIO | both |
| POTS symptoms | 18263 | 2245232 | 813.4127 | 801.6579 | 825.2967 | Male | CPRD GOLD | primary care |
| POTS symptoms | 26589 | 2013678 | 1320.42 | 1304.596 | 1336.388 | Female | CPRD GOLD | primary care |
| POTS symptoms | 37022 | 1065748 | 3473.804 | 3438.507 | 3509.372 | Male | IPCI | primary care |
| POTS symptoms | 53804 | 926439.5 | 5807.611 | 5758.641 | 5856.894 | Female | IPCI | primary care |
| POTS symptoms | 100024 | 7825944 | 1278.108 | 1270.199 | 1286.053 | Male | CPRD Aurum | primary care |
| POTS symptoms | 154729 | 6750301 | 2292.179 | 2280.772 | 2303.629 | Female | CPRD Aurum | primary care |
| POTS diagnosis | 1326 | 2633728 | 50.34689 | 47.67316 | 53.13154 | Male | CPRD GOLD | primary care |
| POTS diagnosis | 2051 | 2657671 | 77.17285 | 73.86876 | 80.58665 | Female | CPRD GOLD | primary care |
| POTS diagnosis | 905 | 1348510 | 67.11111 | 62.80939 | 71.62988 | Male | IPCI | primary care |
| POTS diagnosis | 1295 | 1391211 | 93.08436 | 88.08299 | 98.29572 | Female | IPCI | primary care |
| POTS diagnosis | 9380 | 9235762 | 101.5617 | 99.5167 | 103.6382 | Male | CPRD Aurum | primary care |
| POTS diagnosis | 15040 | 9063834 | 165.9342 | 163.2928 | 168.6077 | Female | CPRD Aurum | primary care |
| ME/CFS symptoms | 10690 | 2393360 | 446.6524 | 438.2251 | 455.2011 | Male | CPRD GOLD | primary care |
| ME/CFS symptoms | 18299 | 2159669 | 847.3058 | 835.0732 | 859.6728 | Female | CPRD GOLD | primary care |
| ME/CFS symptoms | 21705 | 1185702 | 1830.561 | 1806.288 | 1855.078 | Male | IPCI | primary care |
| ME/CFS symptoms | 36162 | 1088278 | 3322.863 | 3288.702 | 3357.29 | Female | IPCI | primary care |
| ME/CFS symptoms | 61890 | 8309948 | 744.77 | 738.9138 | 750.6611 | Male | CPRD Aurum | primary care |
| ME/CFS symptoms | 113520 | 7225417 | 1571.12 | 1561.994 | 1580.287 | Female | CPRD Aurum | primary care |
| ME/CFS diagnosis | 324 | 2644039 | 12.25398 | 10.95586 | 13.66361 | Male | CPRD GOLD | primary care |
| ME/CFS diagnosis | 669 | 2671097 | 25.04589 | 23.1837 | 27.01784 | Female | CPRD GOLD | primary care |
| ME/CFS diagnosis | 102 | 1354240 | 7.5319 | 6.141364 | 9.143213 | Male | IPCI | primary care |
| ME/CFS diagnosis | 331 | 1399161 | 23.65703 | 21.17683 | 26.34791 | Female | IPCI | primary care |
| ME/CFS diagnosis | 1253 | 9288153 | 13.4903 | 12.7536 | 14.25847 | Male | CPRD Aurum | primary care |
| ME/CFS diagnosis | 3419 | 9133208 | 37.43482 | 36.19042 | 38.71109 | Female | CPRD Aurum | primary care |
| MIS | 47 | 9307160 | 0.504988 | 0.371046 | 0.671526 | Male | CPRD Aurum | primary care |
| MIS | 26 | 9179976 | 0.283225 | 0.185012 | 0.41499 | Female | CPRD Aurum | primary care |
| DM | 691 | 2637013 | 26.20389 | 24.28628 | 28.23267 | Male | CPRD GOLD | primary care |
| DM | 547 | 2672336 | 20.46898 | 18.78936 | 22.25848 | Female | CPRD GOLD | primary care |
| DM | 241 | 1352161 | 17.82332 | 15.64392 | 20.22138 | Male | IPCI | primary care |
| DM | 185 | 1399793 | 13.21624 | 11.38033 | 15.26401 | Female | IPCI | primary care |
| DM | 2408 | 9263286 | 25.9951 | 24.96709 | 27.05457 | Male | CPRD Aurum | primary care |
| DM | 1874 | 9146624 | 20.48843 | 19.57121 | 21.43755 | Female | CPRD Aurum | primary care |
| POTS symptoms | 978 | 115705.9 | 845.2466 | 793.096 | 899.9257 | Male | IMASIS | secondary care |
| POTS symptoms | 1170 | 128195.2 | 912.6706 | 861.1172 | 966.5039 | Female | IMASIS | secondary care |
| POTS symptoms | 2575 | 361603.7 | 712.1055 | 684.8638 | 740.1528 | Male | AUSOM | secondary care |
| POTS symptoms | 3415 | 367738.4 | 928.6494 | 897.7616 | 960.3288 | Female | AUSOM | secondary care |
| POTS symptoms | 926 | 2130144 | 43.47124 | 40.71605 | 46.3638 | Male | CHUM | secondary care |
| POTS symptoms | 880 | 2286033 | 38.49463 | 35.99296 | 41.12435 | Female | CHUM | secondary care |
| POTS diagnosis | 334 | 120052.3 | 278.2121 | 249.172 | 309.7074 | Male | IMASIS | secondary care |
| POTS diagnosis | 346 | 134048 | 258.1165 | 231.6325 | 286.7987 | Female | IMASIS | secondary care |
| POTS diagnosis | 229 | 381005.5 | 60.10411 | 52.57104 | 68.41366 | Male | AUSOM | secondary care |
| POTS diagnosis | 229 | 396366 | 57.77488 | 50.53374 | 65.76241 | Female | AUSOM | secondary care |
| POTS diagnosis | 472 | 2134221 | 22.1158 | 20.16538 | 24.20394 | Male | CHUM | secondary care |
| POTS diagnosis | 319 | 2291226 | 13.92268 | 12.43659 | 15.53746 | Female | CHUM | secondary care |
| ME/CFS symptoms | 1036 | 117421 | 882.2952 | 829.3806 | 937.7005 | Male | IMASIS | secondary care |
| ME/CFS symptoms | 1509 | 126752.3 | 1190.511 | 1131.195 | 1252.131 | Female | IMASIS | secondary care |
| ME/CFS symptoms | 676 | 374680.2 | 180.4205 | 167.0744 | 194.5492 | Male | AUSOM | secondary care |
| ME/CFS symptoms | 1028 | 387405.9 | 265.3548 | 249.3796 | 282.0849 | Female | AUSOM | secondary care |
| ME/CFS symptoms | 411 | 2133513 | 19.264 | 17.44638 | 21.21953 | Male | CHUM | secondary care |
| ME/CFS symptoms | 446 | 2288574 | 19.48812 | 17.72122 | 21.3835 | Female | CHUM | secondary care |
| ME/CFS diagnosis | 24 | 121197.3 | 19.80242 | 12.68778 | 29.46443 | Male | IMASIS | secondary care |
| ME/CFS diagnosis | 83 | 135122.5 | 61.42574 | 48.92526 | 76.14644 | Female | IMASIS | secondary care |
| ME/CFS diagnosis | 46 | 382192.3 | 12.03583 | 8.811738 | 16.05411 | Male | AUSOM | secondary care |
| ME/CFS diagnosis | 70 | 397623.5 | 17.60459 | 13.72364 | 22.24234 | Female | AUSOM | secondary care |
| ME/CFS diagnosis | 63 | 2138087 | 2.946559 | 2.264217 | 3.769929 | Male | CHUM | secondary care |
| ME/CFS diagnosis | 54 | 2294084 | 2.35388 | 1.768307 | 3.071303 | Female | CHUM | secondary care |
| MIS | 40 | 2138812 | 1.870197 | 1.336096 | 2.546678 | Male | CHUM | secondary care |
| MIS | 18 | 2294819 | 0.784376 | 0.464871 | 1.239652 | Female | CHUM | secondary care |
| DM | 140 | 120621.1 | 116.0659 | 97.63676 | 136.9623 | Male | IMASIS | secondary care |
| DM | 80 | 134818.6 | 59.33898 | 47.05212 | 73.85249 | Female | IMASIS | secondary care |
| DM | 83 | 381060 | 21.78135 | 17.34872 | 27.00126 | Male | AUSOM | secondary care |
| DM | 58 | 396707 | 14.62036 | 11.10185 | 18.90021 | Female | AUSOM | secondary care |
| DM | 218 | 2134637 | 10.21251 | 8.901744 | 11.66194 | Male | CHUM | secondary care |
| DM | 148 | 2291731 | 6.458001 | 5.45949 | 7.586276 | Female | CHUM | secondary care |
